# Improved detection of multiple sclerosis gadolinium-enhancing lesions: 3D T1 Turbo-Spin-Echo Outperforms 3D T1 Turbo-Field Echo MRI

**DOI:** 10.1101/2025.04.09.25325510

**Authors:** Pablo Naval-Baudin, William F Bermúdez Bravo, Vanessa I Pineda-Borja, Pablo Arroyo-Pereiro, Ignacio Martínez-Zalacaín, Lucía Romero, Paloma Mora, Nahum Calvo, Antonio Martínez-Yélamos, Sergio Martínez-Yélamos, Mònica Cos, Albert Pons-Escoda, Carlos Majós

**Affiliations:** Radiology Department, Hospital Universitari de Bellvitge, L’Hospitalet de Llobregat, Carrer de Feixa Llarga SN, 08907, Barcelona, Spain; Institut de Diagnòstic Per La Imatge (IDI), Centre Bellvitge, L’Hospitalet de Llobregat, 08907, Barcelona, Spain; Translational Imaging Biomarkers Group, Bellvitge Biomedical Research Institute (IDIBELL), L’Hospitalet de Llobregat, 08907, Barcelona, Spain; Departament of Clinical Sciences, School of Medicine, Universitat de Barcelona (UB), L’Hospitalet de Llobregat 08907, Barcelona, Spain; Radiology Department, Hospital Regional Dr Valentin Gomez Farias, El Capullo, 45100 Zapopan, Jal., México; Radiology Department, Instituto Nacional De Ciencias Neurológicas, Lima 15003, Perú; Radiology Department, Hospital Nacional Edgardo Rebagliati Martins, Jesús María 15072, Perú; Multiple Sclerosis Unit, Department of Neurology, Hospital Universitari de Bellvitge. Neurology and Neurogenetics Group. Neuroscience Program, Institut d’Investigació Biomèdica de Bellvitge (IDIBELL). Department of Clinical Sciences, School of Medicine, Universitat de Barcelona (UB), L’Hospitalet de Llobregat, Spain; Head and neck diseases research group, Bellvitge Biomedical Research Institute (IDIBELL), L’Hospitalet de Llobregat, 08907, Barcelona, Spain; Neuro-oncology Unit, Bellvitge Biomedical Research Institute (IDIBELL), L’Hospitalet de Llobregat, 08907, Barcelona, Spain

**Author notes:** Correspondence: Pablo Naval-Baudin, Radiology Department, Hospital Universitari de Bellvitge, L’Hospitalet de Llobregat, Carrer de Feixa Llarga SN, 08907, Barcelona, Spain. X/twitter: @pnavalbaudin.

**Keywords:** Multiple sclerosis, Magnetic resonance imaging, Contrast media, Gadolinium

## Abstract

**Objectives:** To compare the performance of 3D T1 turbo spin echo (3DT1TSE) and 3D T1 turbo field echo (3DT1TFE) MRI in detecting gadolinium-enhancing lesions in multiple sclerosis (MS).

**Methods:** We retrospectively analyzed 255 3T MRIs from MS patients, each including post-contrast 3DT1TSE and 3DT1TFE sequences. Two blinded readers independently assessed enhancing lesions per sequence. A consensus review, incorporating longitudinal imaging and additional sequences, served as the reference standard.

**Results:** The consensus identified 70 enhancing lesions in 31 patients. All 70 were visible on 3DT1TSE, while 64 (91%) were detectable on 3DT1TFE. Reader sensitivity was higher for 3DT1TSE (84% and 90%) than 3DT1TFE (45% and 40%) (p < 0.01). Inter-reader agreement was excellent for 3DT1TSE (ICC = 0.90) and moderate for 3DT1TFE (ICC = 0.69). Although false positives were more common with 3DT1TSE, they were readily excluded during consensus reading. In six patients, enhancing lesions were detected only on 3DT1TSE, with treatment escalation in two.

**Conclusion:** 3DT1TSE outperformed 3DT1TFE in sensitivity and reader agreement for enhancing lesion detection in MS. Incorporating 3DT1TSE into standard MRI protocols may improve disease activity assessment and clinical decision-making.

## Introduction

Multiple sclerosis (MS) is a chronic demyelinating disease of the central nervous system. MRI is essential for MS diagnosis, disease monitoring, and treatment guidance. Gadolinium-based contrast agents (GBCAs) enable detection of enhancing lesions, which reflect active inflammation and influence clinical decision-making ^1–3^.

Three-dimensional T1-weighted spoiled gradient-echo sequences, such as 3D T1 turbo field echo (3DT1TFE), are widely used in post-contrast MS imaging protocols at 3T. More recently, 3D T1 turbo spin echo (3DT1TSE) sequences—commercially known as VISTA, SPACE, or CUBE—have been introduced, offering different image contrast and technical properties ^4,5^.

Earlier work showed 3DT1GRE outperformed 2D T1 spin echo for lesion detection in MS at 3T ^6^, while 3DT1TSE also outperformed 2D T1 spin echo for detecting enhancing MS lesions ^7^. In other neurological conditions, such as brain metastases, 3DT1TSE has been shown to be superior to 3DT1GRE ^8,9^. A recent prospective MS study found that 3DT1TSE identified more radiologically active patients and more enhancing lesions per patient than 3DT1GRE ^10^.

3DT1TSE may enhance lesion detection through several mechanisms: a black-blood effect that reduces vascular artifacts ^4^, higher signal-to-noise and contrast-to-noise ratio ^4,5^, and different T1-weighted contrast properties. While longer post-contrast delays improve lesion visibility ^11–13^. concerns remain about false positives in 3DT1TSE, especially near small veins ^14^.

We hypothesize that 3DT1TSE improves detection of gadolinium-enhancing MS lesions compared to 3DT1TFE in real-world clinical settings, and that false positives can be readily excluded using complementary imaging. This study compares the diagnostic performance and clinical relevance of these sequences and evaluates inter-rater agreement.

## Methods

### Study design

This was a retrospective, cross-sectional, diagnostic accuracy study. The setting was a Spanish public tertiary university hospital, a reference center for MS. Per protocol, all MS neuroimaging procedures were performed using the same Philips Ingenia 3T scanner. Both 3DT1TSE and 3DT1TFE post-contrast sequences, as part of the MS MRI neuroimaging protocol, were performed in our center between February and April 2022 and were components of our continuous update and quality testing scheme. The manuscript structure follows STARD 2015 guidelines ^15^.

### Participants

Initial study candidates were retrieved from our hospital’s MS cohort. This cohort was prospectively followed, and clinical data were systematically structured using the European Database for Multiple Sclerosis (EDMUS) ^16^. Inclusion was voluntary, and all patients signed informed consent to be included in the database.

The inclusion criteria were as follows: 1) Persons diagnosed with MS with systematic follow-up in our MS unit; 2) MRI performed using the same Philips Ingenia 3T scanner in our center between February 1 and April 30, 2022; 3) scan performed with a 32-channel head coil; 4) minimum MRI sequences required: FLAIR, 3DT1TSE, and 3DT1TFE, both post-GBCA. The only exclusion criterion was low-quality imaging after a visual quality filter in either of the two T1-weighted sequences.

A power analysis was conducted to determine the required sample size, indicating a minimum of 158 patients to detect a 20% difference in sensitivity between 3DT1TSE and 3DT1TFE (α = 0.05, power = 90%). We included all eligible patients during the specified 3-month study period to ensure sufficient statistical power and account for potential data loss.

### Imaging data

As specified in the inclusion criteria, all studies were acquired on the same Philips Ingenia 3T scanner, using a 32-channel head coil. When available, alongside post-contrast 3DT1TFE, 3DT1TSE, and FLAIR, we retrieved same-study susceptibility-weighted imaging with phase enhancement (SWIp), along with prior and follow-up post-contrast weighted imaging.

Post-contrast sequences were acquired following DWI and T2-weighted imaging, with 3DT1TSE acquired earlier and 3DT1TFE later. The acquisition order intentionally favored 3DT1TFE, given known improvements in lesion conspicuity with delayed imaging ^11–13^ reported increased sensitivity at 3T using later acquisition times. Detailed timing and protocol parameters are available in the Supplementary Material.

### Clinical data

Demographic and clinical data, including disease duration, EDSS, relapse history, and treatment status, were retrieved from our prospectively maintained MS database (EDMUS) ^16^. DMTs were categorized as none, moderate-, or high-efficacy (see Supplementary Table 1 for details).

### Data preparation

Images were stripped of identifying data and assigned unique study subject identifiers. FLAIR, post-contrast 3DT1TFE, and post-contrast 3DT1TSE sequences were used for blinded reading, with additional sequences retrieved for unblinded consensus reading if necessary.

For blinded reading, post-contrast T1-weighted images were divided into two batches (A and B). Each batch contained all patients, with half having 3DT1TSE and the other half, 3DT1TFE, alternating between batches. FLAIR images were included in both batches. DICOM images were re-anonymized for each batch, ensuring batch-specific identifiers did not correspond to batches or global study identifiers. This re-anonymization process was performed using the DICOM sorting toolkit ^17^.

### Single blinded readings

Blinded readings were conducted by two external radiologists, each reviewing FLAIR plus either 3DT1TSE or 3DT1TFE in two batches with a one-month washout. Batch assignments and re-anonymization procedures are detailed in the Supplementary Material.

### Consensus readings

A consensus reading was conducted by two radiologists together (P.N-B. and A.P-E.), with 6 and 10 years of subspecialized neuroradiology experience, respectively, in an MS reference center. They had access to the results from both blinded readers and the following sequences: pre-contrast 3DT1TFE, post-contrast 3DT1TFE, post-contrast 3DT1TSE, FLAIR, and SWIp. Where available, they also had access to pre-baseline and/or post-baseline post-contrast T1-weighted imaging. An interval of at least six months was sought between the oldest and newest post-contrast T1-weighted image analyzed to guarantee the resolution of acute lesion enhancement.

The consensus reading process involved the following:

- Matching lesions between the two blinded readers. The screen capture of each individual lesion from each reader was matched by visual examination to establish a global lesion database.
- Establishing a comprehensive “gold-standard” reference after simultaneously evaluating all available sequences (post-contrast 3DT1TSE, post-contrast 3DT1TFE, FLAIR, pre-contrast 3DT1TFE, SWIp, and, when available, prior and follow-up post-contrast T1WI); this reference was used to determine whether a suspected lesion was associated with an enhancing MS lesion, or was not a true enhancing MS lesion (e.g., vascular enhancements).
- Independently analyzing each post-contrast sequence (3DT1TSE and 3DT1TFE) to identify whether the lesions were retrospectively enhanced on each sequence.

### Data analysis

MRI sequence acquisition parameters were extracted from the original DICOM files, using a custom script based on the pydicom package in Python version 3.9.13.

Statistical analyses and plots were performed in R version 4.4.1.

For lesion-level analyses, the number of enhancing lesions detected for each sequence (3DT1TSE and 3DT1TFE) and by each reader were calculated. The lesion-level performance metrics calculated included sensitivity, positive predictive value, and F1 score. Because true negatives were unavailable, dependent metrics such as specificity and the negative predictive value were not calculated.

For patient-level analysis, we counted the number of patients with at least one enhancing lesion. Patient-level performance metrics (any patient with an enhancing lesion) included sensitivity, specificity, positive predictive value, negative predictive value, accuracy, and F1 score. The Wilcoxon signed-rank test was used to compare the number of lesions detected per patient on the 3DT1TFE and 3DT1TSE sequences by each reader. The McNemar test was performed to assess the difference in the presence/absence of lesions between the 3DT1TFE and 3DT1TSE sequences, detected by each reader at the patient level. Lesion count per patient was compared between readers for each sequence, with the intra-class correlation coefficient (ICC) for single-fixed raters, using the ‘ICC’ function from the ‘irr’ package in R. Interpretation of ICC values followed commonly accepted guidelines ^18^.

Post-hoc SNR and CNR analyses were performed in a representative subsample to compare lesion conspicuity between sequences. Calculation methods and ROI placement criteria are detailed in the Supplementary Material.

### Clinical impact analysis

Following the lesion-wise and patient-wise analyses, a retrospective review of the EDMUS database was conducted to assess the clinical implications of the imaging findings. This review focused on two key aspects: firstly, subsequent treatment changes in patients in which both readers had the same discrepancy in lesion detection between 3DT1TSE and 3DT1TFE sequences (both readers saw the lesion on one sequence and not on the other); and secondly for cases identified as false positives on 3DT1TSE, the presence of reported enhancing lesions in the original radiological assessment. This additional analysis aimed to contextualize the imaging findings within real-world clinical practice and evaluate the potential impact of utilizing 3DT1TSE for detecting gadolinium-enhancing lesions in MS patients.

## Results

A total of 255 patients met the final selection criteria. Fig. 1 depicts the participant selection process and Table 1 presents the main demographic and clinical characteristics of the study sample.

**Fig. 1.**
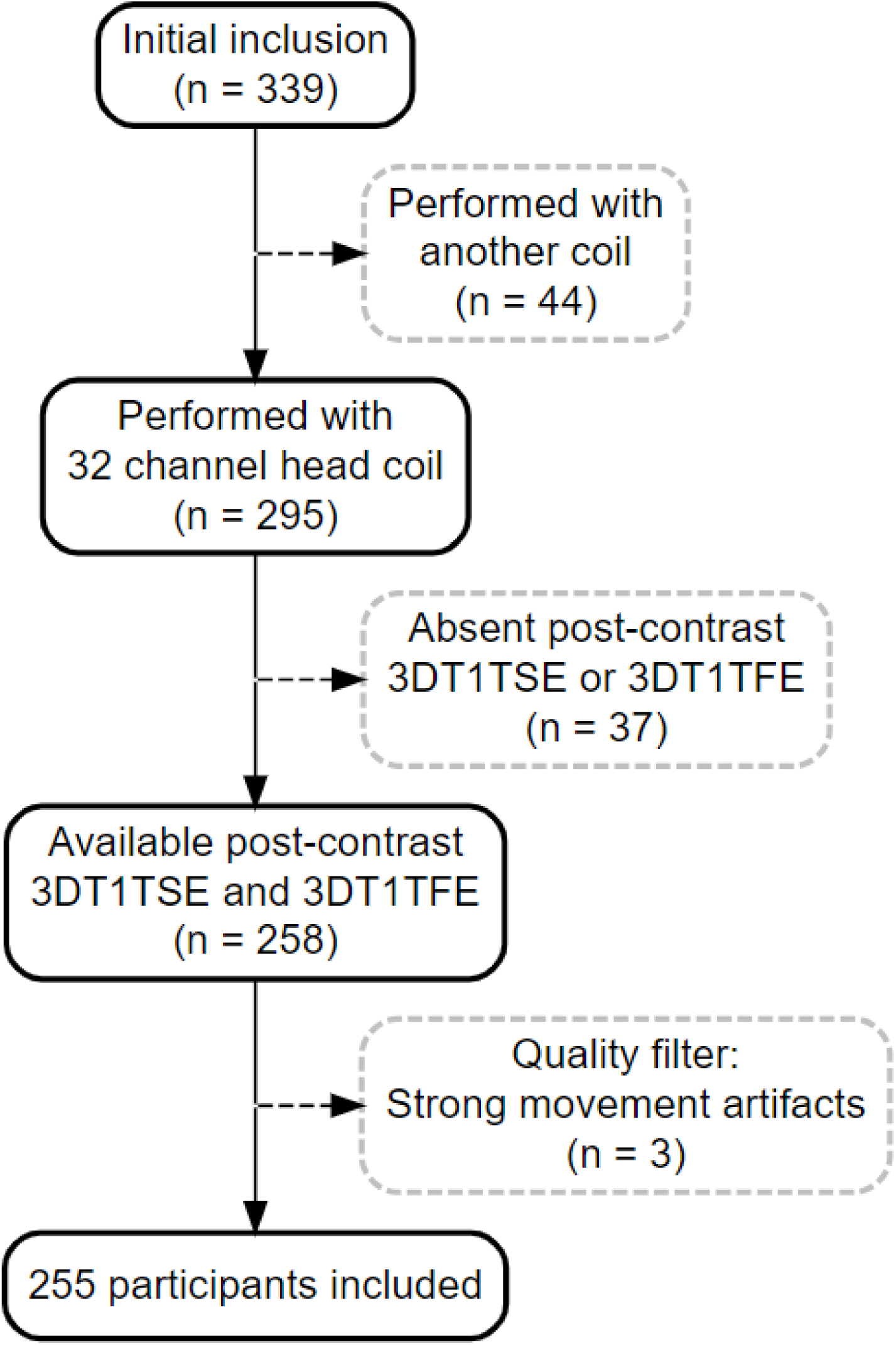
Patient inclusion flowchart. Abbreviations: 3DT1TSE, 3D T1 turbo spin echo; 3DT1TFE, 3D T1 turbo field echo.

**Table 1.**
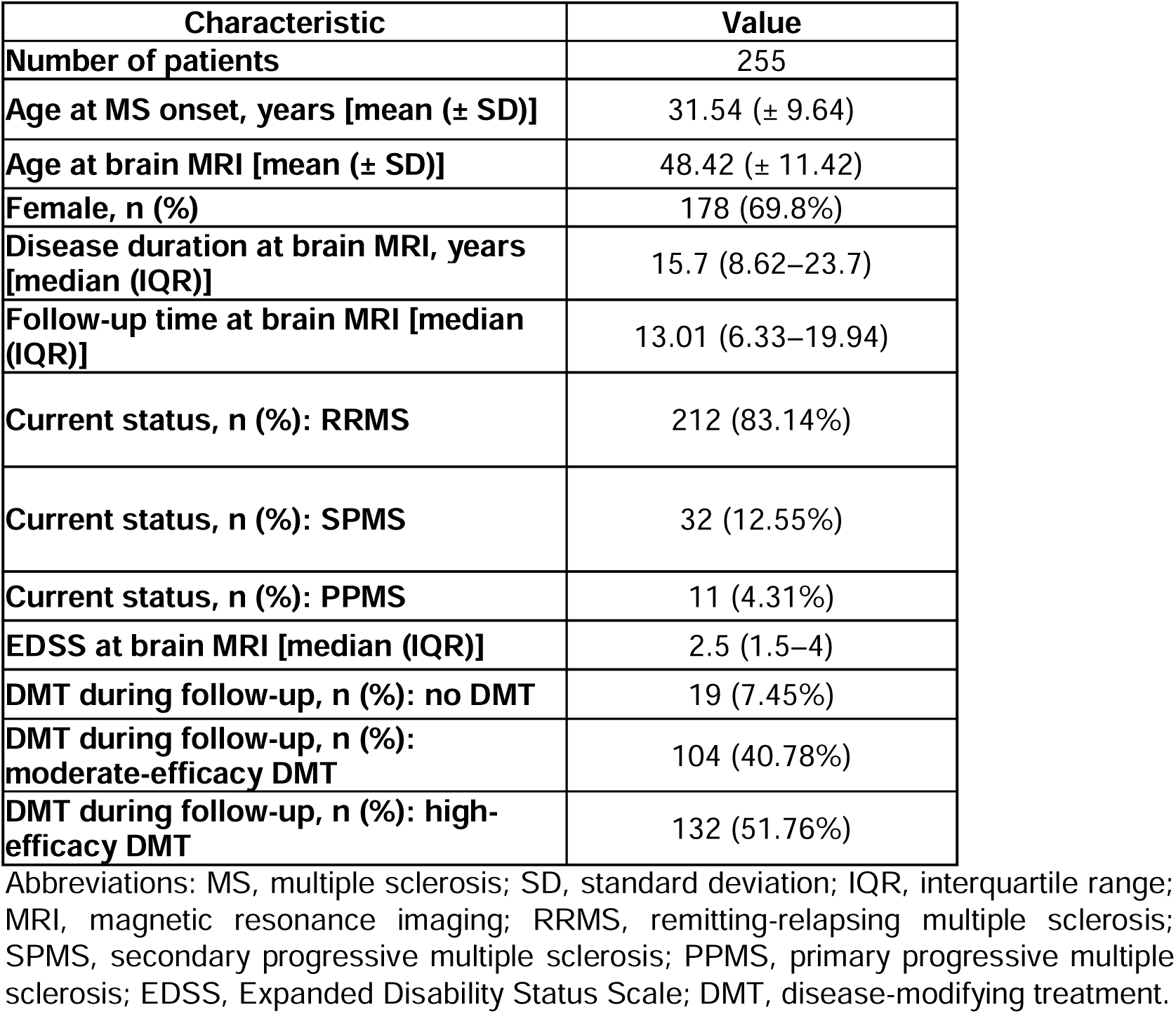
Clinical characteristics of MS cohort at brain MRI.

### Lesion-level analysis

The consensus reading established a reference of 70 enhancing lesions present in 31 of the 255 patients. All 70 of these lesions were visible on 3DT1TSE, whereas 64 lesions (91.4%) were visible on 3DT1TFE.

Of the 70 lesions visible on 3DT1TSE, Reader 1 identified 59 (84%) and Reader 2 identified 63 (90%). Of the 64 lesions visible on 3DT1TFE, Reader 1 identified 29 (45%) and Reader 2 identified 26 (40%). Reader 1 identified five false-positive lesions on 3DT1TSE, whereas Reader 2 identified nine false-positive lesions. On the other hand, each reader identified one false-positive lesion on 3DT1TFE.

Fig. 2a presents a detailed lesion-level analysis of the enhancing MS lesions detected on the 3DT1TFE and 3DT1TSE sequences. This figure shows the number of lesions detected, missed, and falsely identified by each reader for both sequences. Table 2 presents an analysis of lesion-level performance metrics for each sequence and reader.

**Fig. 2.**
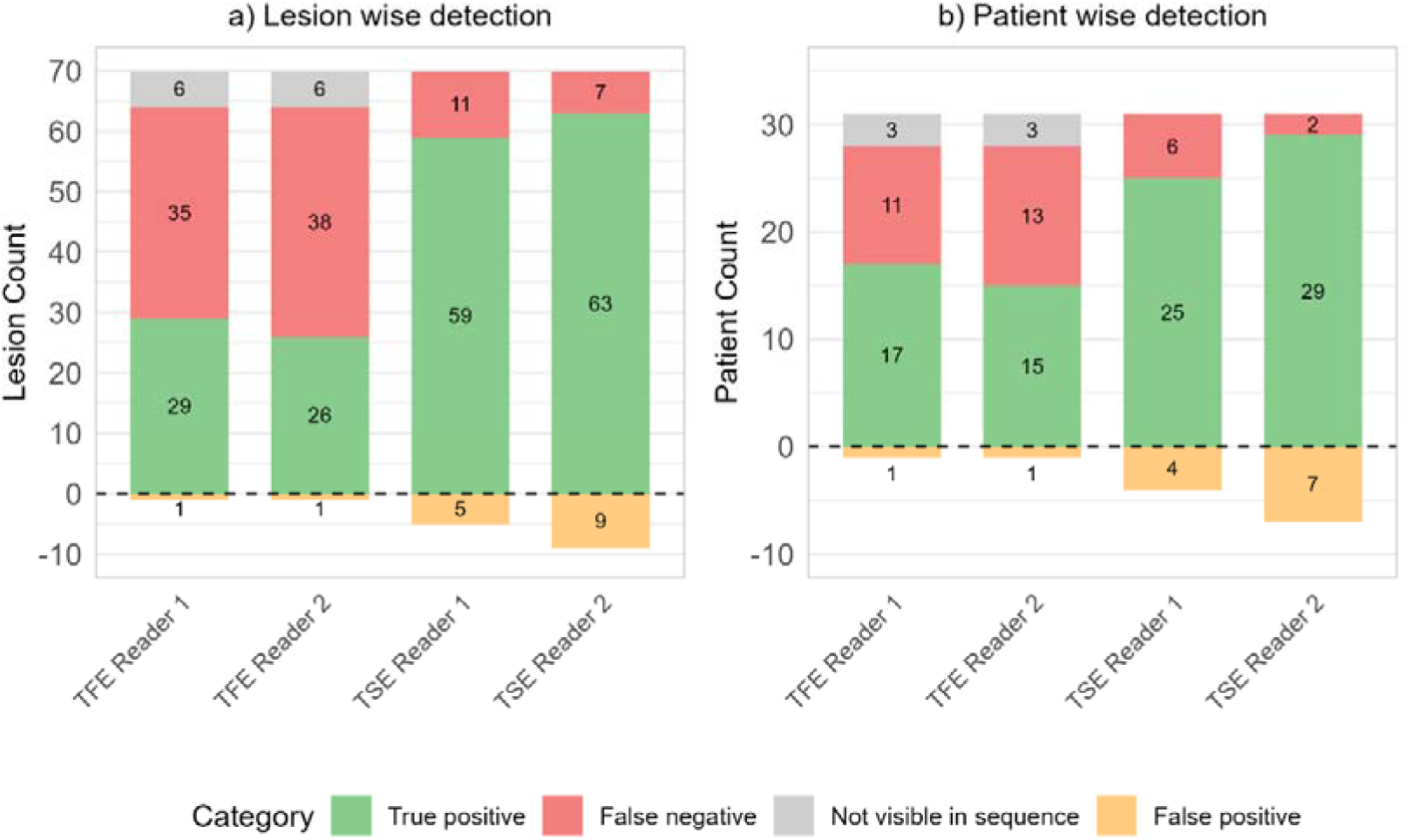
Detection success by sequence and reader. **(a)** Lesion-wise detection. **(b)** Patient-wise detection. The stacked bar charts show true positives, missed cases, cases not visible in sequence, and false positives for 3D T1 turbo-field-echo (TFE) and 3D T1 turbo-spin-echo (TSE) sequences.

**Table 2.** Lesion-wise performance metrics for each sequence and reader. In the lesion-wise analysis, true negative data are unavailable; thus, metrics such as specificity and negative predictive value (NPV) cannot be calculated.

| **Metric** | **Reader 1 TFE** | **Reader 2 TFE** | **Reader 1 TSE** | **Reader 2 TSE** |
| --- | --- | --- | --- | --- |
| **Sensitivity** | 0.45 | 0.41 | 0.84 | 0.90 |
| **PPV** | 0.97 | 0.96 | 0.92 | 0.88 |
| **F1 score** | 0.62 | 0.57 | 0.88 | 0.89 |

Abbreviations: 3D T1-weighted turbo field echo (TFE), 3D T1-weighted turbo spin echo (TSE), negative predictive value (NPV).

Figs. 3 to 6 display examples of the lesions identified on post-contrast 3DT1TSE but missed or absent on 3DT1TFE. Screen captures of all lesions included in the study are available as Supplementary Figs. 1 to 83 to ensure full transparency.

**Fig. 3.**
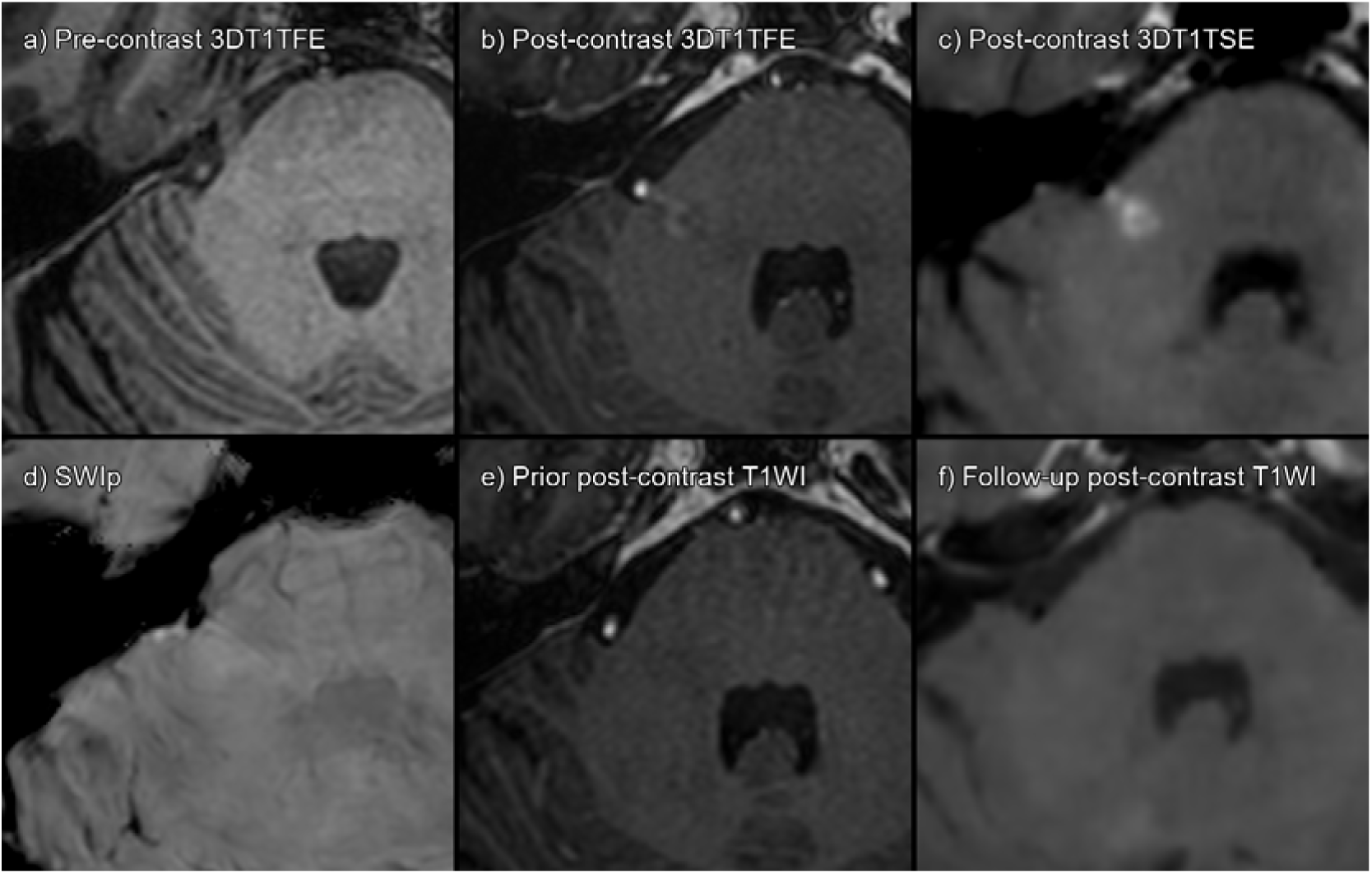
True enhancing lesion. **(a)** 3DT1TFE without contrast. **(b)** 3DT1TFE post-contrast. **(c)** 3DT1TSE post-contrast. **(d)** SWIp. **(e)** Prior post-contrast 3DT1TFE, 4 years pre-baseline. **(f)** Follow-up post-contrast 3DT1TSE, 6 months post-baseline. Enhancing right cerebellar peduncle lesion; this lesion was missed by one of the readers on 3DT1TFE but detected by both readers on 3DT1TSE.

**Fig. 4.**
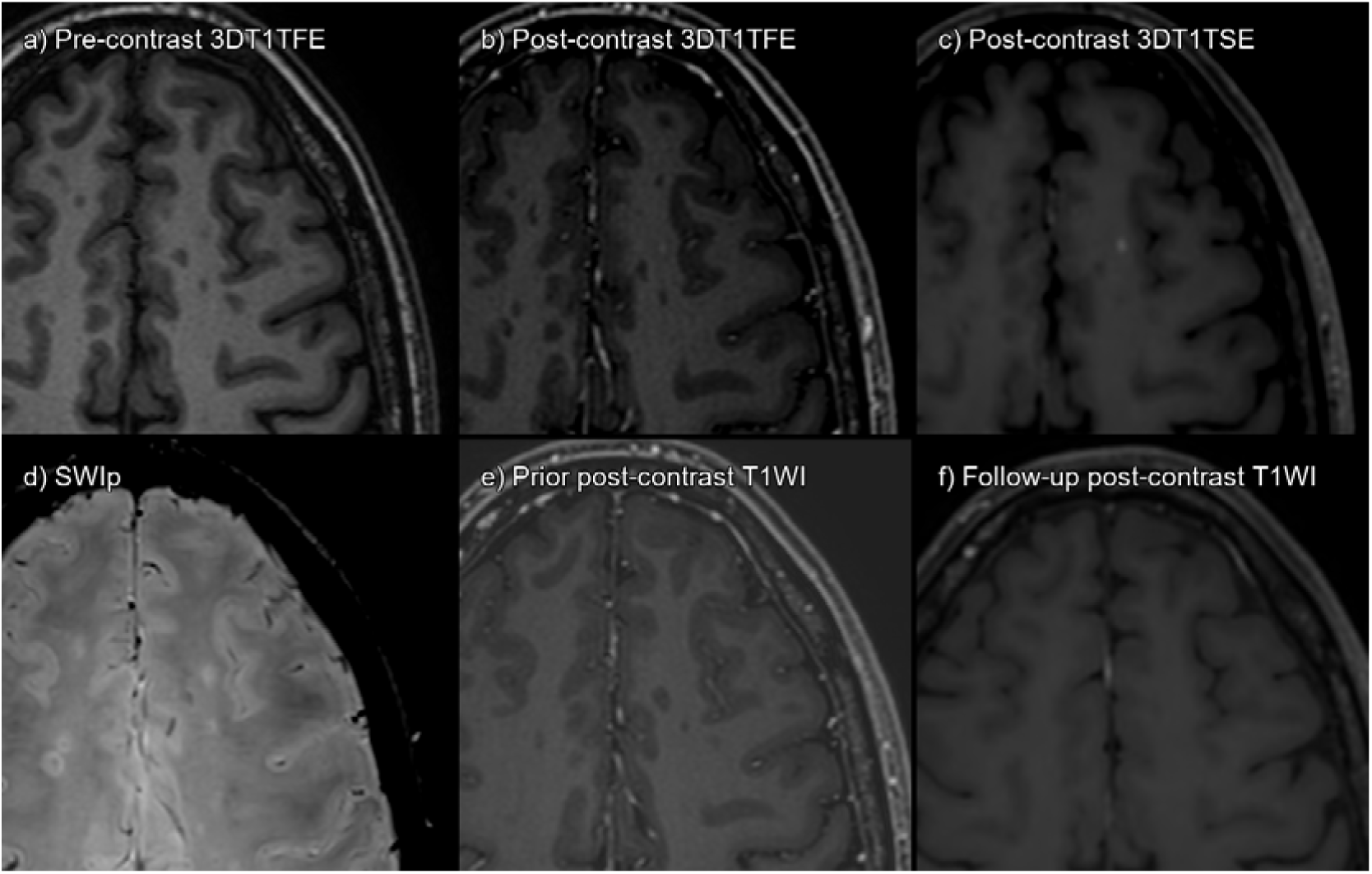
True enhancing lesion, missed by both readers on 3DT1TFE, but subtle enhancement detected on consensus reading. **(a)** 3DT1TFE without contrast. **(b)** 3DT1TFE post-contrast. **(c)** 3DT1TSE post-contrast. **(d)** SWIp. **(e)** Prior post-contrast 3DT1TFE, 15 months pre-baseline. **(f)** Follow-up post-contrast 3DT1TSE, 5 months post-baseline. Enhancing left subcortical superior frontal lesion; this lesion was missed by both readers on 3DT1TFE but detected by both on 3DT1TSE.

**Fig. 5.**
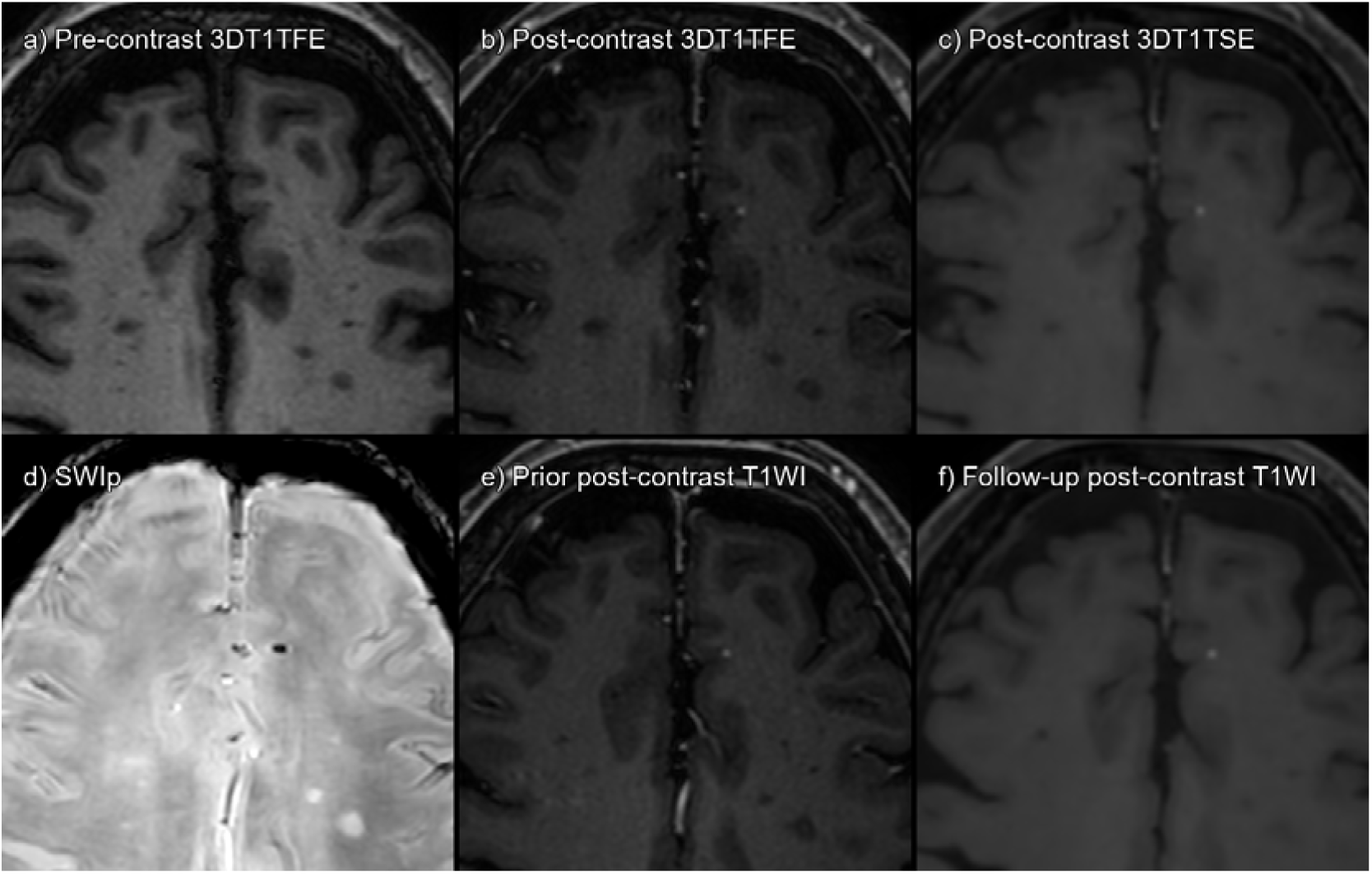
False positives on both sequences. **(a)** 3DT1TFE without contrast. **(b)** 3DT1TFE post-contrast. **(c)** 3DT1TSE post-contrast. **(d)** SWIp. **(e)** Prior post-contrast 3DT1TFE, 4 months pre-baseline. **(f)** Follow-up post-contrast 3DT1TSE, 7 months post-baseline. Dotlike subcortical enhancement is present both on TSE and TFE. However, it is already present in prior and follow-up imaging, and on SWIp, it can be seen as a dotlike paramagnetic lesion. Probable small, cavernous angioma. Detected as a false positive by both readers on TSE but by neither on TFE.

**Fig. 6.**
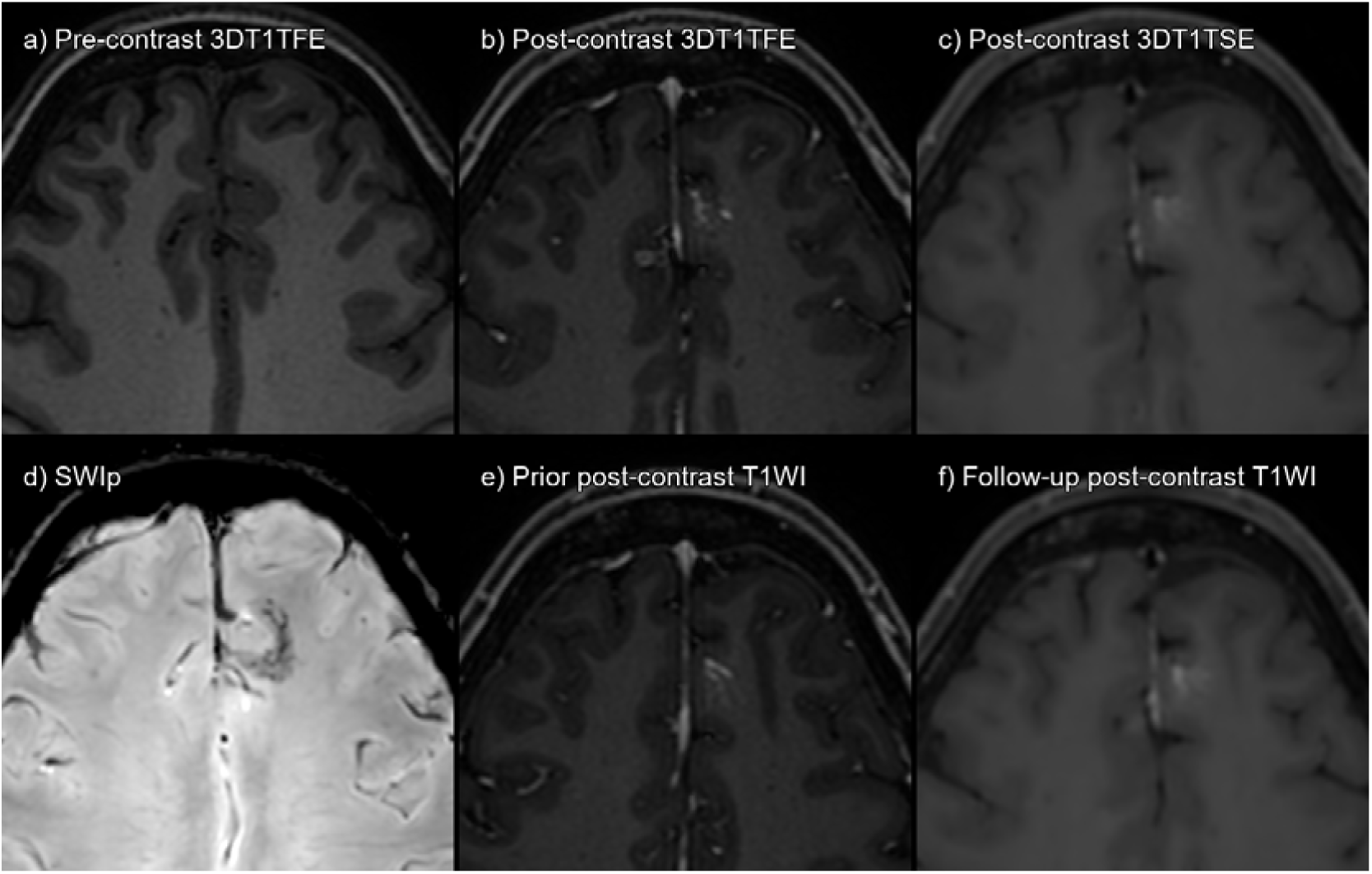
False positives on 3DT1TSE. **(a)** 3DT1TFE without contrast. **(b)** 3DT1TFE post-contrast. **(c)** 3DT1TSE post-contrast. **(d)** SWIp. **(e)** Prior post-contrast 3DT1TFE, 15 months pre-baseline. **(f)** Follow-up post-contrast 3DT1TSE, 13 months post-baseline. Left prefrontal parasagittal developmental venous anomaly (DVA). One of the readers mistakenly labeled this an enhancing lesion on 3DT1TSE. The *caput medusae* sign typical of DVAs is evident on 3DT1TFE. However, it is also easily detectable as a false positive if SWIp, prior, or follow-up imaging is available.

### Patient-level analysis

The consensus reading established that 31 patients had at least one enhancing lesion on 3DT1TSE, whereas 28 patients had at least one enhancing lesion on 3DT1TFE. Reader 1 identified 25 (80%), and Reader 2 identified 29 (94%) of the 31 patients with at least one enhancing lesion on 3DT1TSE. Moreover, Reader 1 detected four false-positive patients and Reader 2 identified seven false-positive patients. In addition, of the 28 patients with at least one enhancing lesion on 3DT1TFE, Reader 1 identified 17 (61%) and Reader 2 identified 15 (54%), with each reader identifying one false-positive patient.

Fig. 2b presents a patient-level analysis of enhancing MS lesions detected by 3DT1TFE and 3DT1TSE sequences. This figure shows the number of patients correctly identified as having at least one enhancing lesion, the number of patients missed, and the number of false-positive patients for each reader and sequence. For a more detailed evaluation of patient-level performance, including metrics such as sensitivity, specificity, and accuracy, refer to Table 3.

**Table 3.**
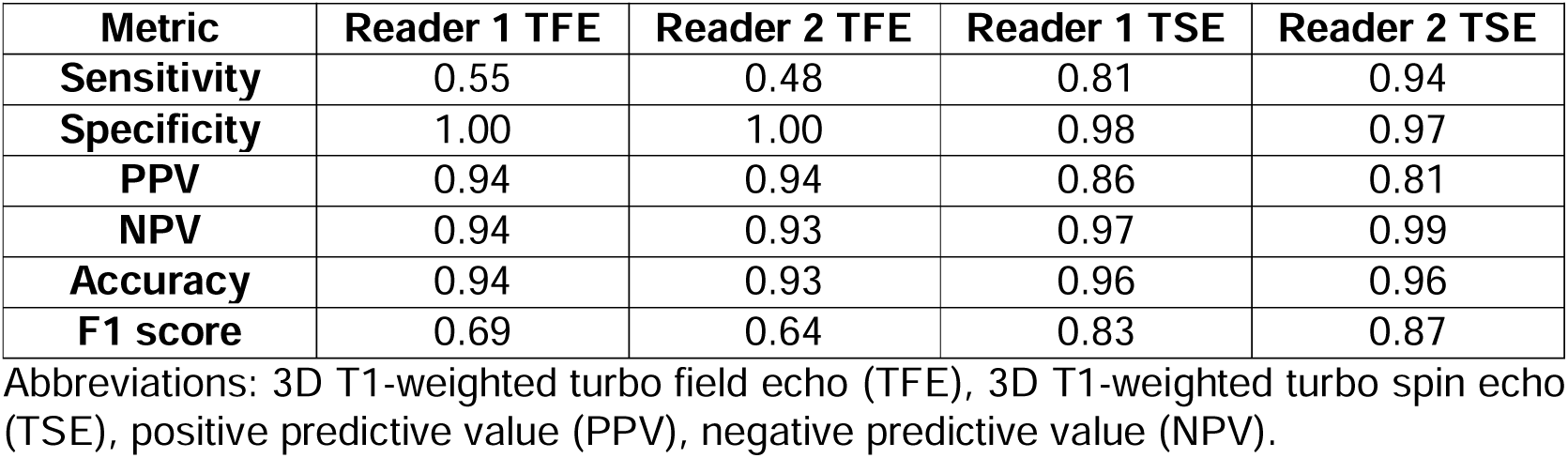
Patient-wise performance metrics for each sequence and reader.

| **Metric** | **Reader 1 TFE** | **Reader 2 TFE** | **Reader 1 TSE** | **Reader 2 TSE** |
| --- | --- | --- | --- | --- |
| **Sensitivity** | 0.55 | 0.48 | 0.81 | 0.94 |
| **Specificity** | 1.00 | 1.00 | 0.98 | 0.97 |
| **PPV** | 0.94 | 0.94 | 0.86 | 0.81 |
| **NPV** | 0.94 | 0.93 | 0.97 | 0.99 |
| **Accuracy** | 0.94 | 0.93 | 0.96 | 0.96 |
| **F1 score** | 0.69 | 0.64 | 0.83 | 0.87 |

Abbreviations: 3D T1-weighted turbo field echo (TFE), 3D T1-weighted turbo spin echo (TSE), positive predictive value (PPV), negative predictive value (NPV).

The Wilcoxon signed-rank test, comparing the number of lesions detected per patient on the 3DT1TFE and 3DT1TSE sequences, revealed a significant difference between these sequences for Reader 1 (*p* < 0.001) and Reader 2 (*p* < 0.001).

McNemar’s test for the presence/absence of lesions showed a significant difference between 3DT1TFE and 3DT1TSE for Reader 1 (χ^2^ = 7.69, *p* = 0.0056) and Reader 2 (χ^2^ = 16.4, *p* < 0.001).

The correlation for the total lesion count per patient demonstrated excellent reliability for 3DT1TSE (ICC = 0.90) and moderate for 3DT1TFE (ICC = 0.69).

Overall, this analysis demonstrates that 3DT1TSE outperformed 3DT1TFE in correctly identifying patients with enhancing lesions, whereas 3DT1TFE resulted in fewer false positives.

### Signal and contrast to noise ratio analyses

In a subset of ten patients, we performed a post-hoc SNR and CNR analysis, to quantitatively assess lesion conspicuity on the two post-contrast sequences. For 3DT1TFE, the mean SNR was 363.80 ± 280.04, whereas for 3DT1TSE it was 495.49 ± 270.77. Similarly, the mean CNR was 95.86 ± 59.35 for 3DT1TFE and 159.58 ± 80.66 for 3DT1TSE. Wilcoxon signed-rank tests demonstrated that these differences were statistically significant, both for SNR (*p* = 0.0099) and CNR (*p* = 0.0029).

### Clinical impact

There were six patients for which both readers detect in agreeance at least one true enhancing lesion on 3DT1TSE but not on 3DT1TFE. Of these six patients, two had their treatment switched from moderate- to high-efficacy drugs because these enhancing lesions were detected on MRI.

Regarding false positives, one patient had at least one false-positive enhancing lesion detected by both readers on 3DT1TSE, and nine more patients had at least one false-positive lesion detected by one of the two readers. Upon review of the results of the radiological reports for all these patients, all were correctly classified as true negatives in the real-world radiological report.

## Discussion

This study demonstrates that 3DT1TSE sequences offer superior sensitivity in detecting gadolinium-enhancing lesions in MS compared to 3DT1TFE sequences. Readers consistently displayed better sensitivity with 3DT1TSE. Inter-rater reliability was higher for 3DT1TSE, indicating better consistency in lesion detection. Notably, blinded readers identified less than half of the enhancing lesions on 3DT1TFE, whereas false positives on 3DT1TSE were easily identifiable as true negatives on consensus readings with access to a full MRI exam.

Our study also demonstrates the real-world impact of using a more sensitive technique; six patients had true enhancing lesions detected by both readers on 3DT1TSE and not 3DT1TFE. In two of these patients, detecting these lesions contributed to treatment changes, illustrating the potential clinical significance of using more sensitive MRI techniques.

Our findings align with recent studies, which demonstrated that 3DT1TSE detected significantly more radiologically active patients and contrast-enhanced lesions per patient compared to 3DGRE T1-WI ^10^. Moreover, our results challenge concerns about false positives on 3DT1TSE, as raised by some authors ^14^. Instead, we found a considerable number of false negatives on 3DT1TFE, suggesting a potential underestimation of disease activity when relying solely on this sequence. This finding has major implications for MS management, given the critical role of accurate disease activity assessment and the importance of precise prognostic biomarkers ^19^.

The detection of enhancing lesions via MRI plays a crucial role in treatment decision-making for patients with MS. International guidelines ^20–23^ recommend considering treatment changes based on MRI evidence of disease activity. Here, using 3DT1TSE sequences allowed the detection of enhancing lesions that were altogether not visible on 3DT1TFE sequences for six patients, while more than half of the lesions that actually did enhance on 3DT1TFE were missed by the readers on this sequence.

The higher sensitivity of 3DT1TSE for gadolinium-enhancing lesion detection can be attributed to several factors. The inherent “black blood” effect of 3DT1TSE, whereby the lumen of blood vessels appears hypointense, facilitates lesion detection by reducing eye fatigue from enhancing structures such as cortical veins ^4,24^. Furthermore, the different contrast mechanisms of 3DT1TSE may be more sensitive to gadolinium’s T1-shortening effects. Technical factors such as a higher signal- and contrast-to-noise ratio and reduced susceptibility artifacts in 3DT1TSE may also contribute to improved lesion detection, particularly in artifact-prone regions ^5^.

The superior sensitivity of 3DT1TSE may lead to earlier and more reliable identification of disease activity, which is particularly relevant given the growing emphasis on early and effective treatment in MS to prevent long-term disability ^25^. By more accurately identifying active inflammation, clinicians could make more informed decisions about treatment escalation or de-escalation, potentially improving patient outcomes and quality of life ^23^.

Our findings’ implications extend beyond clinical practice and could impact clinical trials for MS. The increased sensitivity in detecting active lesions could lead to a more accurate assessment of disease activity, potentially allowing for the earlier detection of treatment effects with reduced sample sizes ^26^. On the other hand, this improved lesion detection could influence how “no evidence of disease activity” is defined in trials, potentially making this endpoint more stringent ^27^.

Nevertheless, non-contrast 3DT1TFE still holds substantial value in MS imaging. Pre-contrast 3DT1TFE offers better anatomical definition, facilitates atrophy calculations, and allows for sub-region segmentation. It also provides crucial information on deeply T1-hypointense voxels, characteristic of paramagnetic rim lesions ^28^. Recent research has identified a novel “T1-dark rim” sign on 3DT1TFE, which may serve as an accessible surrogate marker for chronic active lesions ^29^. Hence, it may be advisable to perform 3DT1TFE for non-contrast imaging and 3DT1TSE for post-contrast imaging.

Our study also has several limitations. First, it was performed at a single center, using one scanner and a retrospective design, which may affect the generalizability of the results. Second, we relied on purely visual assessment without automated detection tools, introducing a potential for human error—though this reflects typical real-world practice. Third, the fixed order of sequence acquisition (3DT1TSE followed by 3DT1TFE) can be considered a limitation because scans performed later are known to benefit from increased lesion conspicuity due to greater gadolinium wash-in ^11–13^. Paradoxically, this timing bias should have favored 3DT1TFE, yet it still underperformed compared to 3DT1TSE. Because this was a retrospective study aligned with our routine clinical workflow, reversing or randomizing the order was not feasible; however, future prospective protocols could address this by acquiring sequences in random or alternate orders to isolate the effect of timing from sequence characteristics.

By contrast, our work also has notable strengths. We analyzed a large sample of 255 MS patients using standardized protocols on the same 3T scanner, minimizing hardware and acquisition variability. The scans were interpreted by external radiologists blinded to the study’s hypotheses, limiting reader bias. Finally, we performed a comprehensive consensus reading that leveraged multiple sequences (including prior imaging) to establish a robust reference standard. This design maximizes diagnostic accuracy while aligning with current clinical practice.

In conclusion, our findings provide compelling evidence for the superior performance of 3DT1TSE in detecting gadolinium-enhancing lesions in MS. This enhanced detection capability could impact both clinical practice and research, potentially leading to more accurate disease monitoring and treatment decisions. Thus, future multi-center studies with long-term follow-up are needed to validate these findings across different clinical settings and assess their impact on patient outcomes.

## Data Availability

The data that support the findings of this study are available from the corresponding author upon reasonable request.

## Abbreviations

MS: multiple sclerosis
3DT1TSE: 3D T1-weighted turbo spin echo
3DT1TFE: 3D T1-weighted turbo field echo
GBCA: gadolinium-based contrast agent
FLAIR: fluid-attenuated inversion recovery
EDSS: expanded disability status scale
DMT: disease-modifying treatment
ICC: intra-class correlation coefficient
PRL: paramagnetic rim lesion
SWIp: susceptibility-weighted imaging with phase enhancement
SNR: signal to noise ratio
CNR: contrast to noise ratio

## Statements and Declarations

### Ethical Considerations

This study was approved by the Research Ethics Committee of Bellvitge University Hospital (reference PR192/24). The requirement for informed consent was waived due to the retrospective observational nature of the study and compliance with national and European Union regulations. All data were anonymized prior to analysis.

### Consent to Participate

Not applicable.

### Consent for Publication

Not applicable.

## References

1. Filippi M, Bar-Or A, Piehl F, et al. Multiple sclerosis. Nat Rev Dis Prim 2018; 4: 43.

2. Thompson AJ, Banwell BL, Barkhof F, et al. Diagnosis of multiple sclerosis: 2017 revisions of the McDonald criteria. Lancet Neurol 2018; 17: 162–173.

3. Wattjes MP, Ciccarelli O, Reich DS, et al. 2021 MAGNIMS–CMSC–NAIMS consensus recommendations on the use of MRI in patients with multiple sclerosis. Lancet Neurol 2021; 20: 653–670.

4. Bapst B, Amegnizin J-L, Vignaud A, et al. Post-contrast 3D T1-weighted TSE MR sequences (SPACE, CUBE, VISTA/BRAINVIEW, isoFSE, 3D MVOX): Technical aspects and clinical applications. J Neuroradiol 2020; 47: 358–368.

5. Park J, Mugler JP, Horger W, et al. Optimized T1-weighted contrast for single-slab 3D turbo spin-echo imaging with long echo trains: application to whole-brain imaging. Magn Reson Med 2007; 58: 982–92.

6. Crombé A, Saranathan M, Ruet A, et al. MS lesions are better detected with 3D T1 gradient-echo than with 2D T1 spin-echo gadolinium-enhanced imaging at 3T. AJNR Am J Neuroradiol 2015; 36: 501–7.

7. Hodel J, Outteryck O, Ryo E, et al. Accuracy of postcontrast 3D turbo spin-echo MR sequence for the detection of enhanced inflammatory lesions in patients with multiple sclerosis. AJNR Am J Neuroradiol 2014; 35: 519–23.

8. Reichert M, Morelli JN, Runge VM, et al. Contrast-enhanced 3-dimensional SPACE versus MP-RAGE for the detection of brain metastases: considerations with a 32-channel head coil. Invest Radiol 2013; 48: 55–60.

9. Komada T, Naganawa S, Ogawa H, et al. Contrast-enhanced MR imaging of metastatic brain tumor at 3 tesla: utility of T(1)-weighted SPACE compared with 2D spin echo and 3D gradient echo sequence. Magn Reson Med Sci 2008; 7: 13–21.

10. De Panafieu A, Lecler A, Goujon A, et al. Contrast-Enhanced 3D Spin Echo T1-Weighted Sequence Outperforms 3D Gradient Echo T1-Weighted Sequence for the Detection of Multiple Sclerosis Lesions on 3.0 T Brain MRI. Invest Radiol 2023; 58: 314–319.

11. Filippi M, Yousry T, Rocca MA, et al. Sensitivity of delayed gadolinium-enhanced MRI in multiple sclerosis. Acta Neurol Scand 1997; 95: 331–4.

12. Silver NC, Good CD, Barker GJ, et al. Sensitivity of contrast enhanced MRI in multiple sclerosis. Effects of gadolinium dose, magnetization transfer contrast and delayed imaging. Brain 1997; 120 (Pt 7: 1149–61.

13. Rovira A, Auger C, Huerga E, et al. Cumulative Dose of Macrocyclic Gadolinium-Based Contrast Agent Improves Detection of Enhancing Lesions in Patients with Multiple Sclerosis. AJNR Am J Neuroradiol 2017; 38: 1486–1493.

14. Danieli L, Roccatagliata L, Distefano D, et al. Nonlesional Sources of Contrast Enhancement on Postgadolinium “Black-Blood” 3D T1-SPACE Images in Patients with Multiple Sclerosis. Am J Neuroradiol 2022; 43: 872–880.

15. Bossuyt PM, Reitsma JB, Bruns DE, et al. STARD 2015: An Updated List of Essential Items for Reporting Diagnostic Accuracy Studies. Radiology 2015; 277: 826–32.

16. Confavreux C, Compston DA, Hommes OR, et al. EDMUS, a European database for multiple sclerosis. J Neurol Neurosurg Psychiatry 1992; 55: 671–6.

17. Naval-Baudin P. DICOM Sorting Toolkit. Zenodo. Epub ahead of print 2024. DOI: 10.5281/zenodo.13094029.

18. Koo TK, Li MY. A Guideline of Selecting and Reporting Intraclass Correlation Coefficients for Reliability Research. J Chiropr Med 2016; 15: 155–63.

19. Naval-Baudin P, Arroyo-Pereiro P, Majós C. The pressing need for imaging biomarkers of disability progression in multiple sclerosis. Eur Radiol 2024; 34: 3823–3825.

20. NICE. Multiple sclerosis in adults: management. NICE Clin Guidel CG186 2014; 20–22.

21. Rae-Grant A, Day GS, Marrie RA, et al. Practice guideline recommendations summary: Disease-modifying therapies for adults with multiple sclerosis. Neurology 2018; 90: 777–788.

22. Meca-Lallana JE, Martínez Yélamos S, Eichau S, et al. Consensus statement of the Spanish Society of Neurology on the treatment of multiple sclerosis and holistic patient management in 2023. Neurologia 2024; 39: 196–208.

23. Montalban X, Gold R, Thompson AJ, et al. ECTRIMS/EAN Guideline on the pharmacological treatment of people with multiple sclerosis. Mult Scler J 2018; 24: 96–120.

24. Zhu C, Graves MJ, Yuan J, et al. Optimization of improved motion-sensitized driven-equilibrium (iMSDE) blood suppression for carotid artery wall imaging. J Cardiovasc Magn Reson 2014; 16: 61.

25. Giovannoni G, Butzkueven H, Dhib-Jalbut S, et al. Brain health: time matters in multiple sclerosis. Mult Scler Relat Disord 2016; 9: S5–S48.

26. Sormani MP, Bruzzi P. MRI lesions as a surrogate for relapses in multiple sclerosis: a meta-analysis of randomised trials. Lancet Neurol 2013; 12: 669–76.

27. Havrdova E, Galetta S, Hutchinson M, et al. Effect of natalizumab on clinical and radiological disease activity in multiple sclerosis: a retrospective analysis of the Natalizumab Safety and Efficacy in Relapsing-Remitting Multiple Sclerosis (AFFIRM) study. Lancet Neurol 2009; 8: 254–60.

28. Naval-Baudin P, Pons-Escoda A, Camins À, et al. Deeply 3D-T1-TFE hypointense voxels are characteristic of phase-rim lesions in multiple sclerosis. Eur Radiol 2024; 34: 1337–1345.

29. Naval-Baudin P, Pons-Escoda A, Castillo-Pinar A, et al. The T1-dark-rim: A novel imaging sign for detecting smoldering inflammation in multiple sclerosis. Eur J Radiol 2024; 173: 111358.

