## Supplementary for "Improved detection of multiple sclerosis gadolinium-enhancing lesions: 3D T1 Turbo-Spin-Echo Outperforms 3D T1 Turbo-Field Echo MRI"

#### SEQUENCE ACQUISITION TIMING

A DWI sequence lasting 1:41 minutes and a T2WI sequence lasting 2:55 minutes were acquired immediately after GBCA contrast administration and before any of the two post-contrast T1WI sequences. After DWI and T2WI, a 3DT1TSE was acquired, lasting 4:47 minutes on average. After the DWI, T2WI and the 3DT1TSE sequences, the 3DT1TFE was acquired, lasting 5:47 minutes on average. Therefore, the minimum mean delay before 3DT1TSE initiation after contrast administration was 4:36 min and for 3DT1TFE it was 9:23 min. The more delayed acquisition of 3DT1TFE was intended to give an advantage to 3DT1TFE.

#### SEQUENCE ACQUISITION PARAMETERS

3DT1TFE sequence parameters were as follows: Sagittal acquisition plane, in plane reconstruction voxel size of 0.46 x 0.46 mm with 0.5 mm slice thickness. Repetition time shortest (mean 9.76 ms, standard deviation 0.14 ms, range 8.12 – 10.11 ms), echo time shortest (mean 4.63 ms, standard deviation 0.09 ms, range 3.73 – 4.86), and flip angle 8°, turbo-field echo inversion pulse delay shortest (mean 1035.48 ms, standard deviation 26.08 ms, range 699.51 – 1155.95 ms), SENSE acceleration with factor 1.5, and duration (mean 05:47 min, standard deviation 00:37, range 04:20 - 06:15 range).

3DT1TSE sequence parameters: Sagittal acquisition plane, in-plane reconstruction pixel spacing 0.333 x 0.333 mm, spacing between slices 0.5 mm, slice thickness 1 mm, repetition time 600 ms, echo time shortest (mean 28.28 ms standard deviation 0.175 ms, range 26.84 – 28.33), flip angle 90°, CS-SENSE acceleration with factor 2, and duration (mean 04:47 min, standard deviation 00:19 min, range 03:32 - 05:23 min).

FLAIR sequence parameters: Sagittal acquisition plane, in-plane reconstruction pixel spacing 0.746 x 0.746 mm, spacing between slices 0.55 mm, slice thickness 1.1 mm, repetition time 5500 ms, echo time shortest (mean 296.15 ms, standard deviation 7.45 ms, range 283.67 – 319.98 ms), inversion time 1700 ms, flip angle 90°, SENSE acceleration with factor 3, and duration (mean 05:17 min, standard deviation 00:17 min, range 04:19 - 05:58 min).

SWIp sequence parameters: Axial acquisition plane, in-plane reconstruction pixel spacing 0.53 x 0.53 mm, spacing between slices 1 mm, slice thickness 2 mm, repetition time 50 ms, number of echoes 6, echo times = 7.2, 13.4, 19.6, 25.8, 32, and 38.2 ms, flip angle 12°, CS-SENSE acceleration with factor 1.7, and duration (4:23 min).

The gadolinium-based contrast agent (GBCA) was Gadobutrol (Gadovist® Bayer Hispania S.L.), administered intravenously manually, at a dosage relative to patient weight of 0.1 mmol/kg and concentration of 1 mmol/ml. The imaging sequence order of acquisition immediately after GBCA administration, was: First T2 turbo-spin-echo weighted imaging (with a mean duration of 2:55 min), second 3DT1TSE (with a mean duration of 4:47 min), and last 3DT1TFE (with a mean duration of 5:47 min). FLAIR imaging was acquired before intravenous GBCA administration.

#### DISEASE MODIFYING TREATMENT CLASSIFICATION

Moderate efficacy disease modifying treatment was considered if any of the following was being used at any moment of the follow-up: interferon beta 1a, interferon beta 1b, peginterferon beta 1a, glatiramer acetate, teriflunomide, dimethyl fumarate, cladribine, fingolimod, siponimod, ozanimod, and ponesimod.

High efficacy disease modifying treatment was considered if any of the following was being used at any moment of the follow-up: natalizumab, alemtuzumab, ocrelizumab, ofatumumab, rituximab, mitoxantrone and cyclophosphamide.

#### SINGLE BLINDED READINGS

The blinded readings were performed by two readers (WF.B.V. and VI.P.B.) external to our center and unaware of the study's hypotheses. Both readers were more accustomed to using 3DT1TFE or 3DT1GRE-like imaging than 3DT1TSE in their respective centers. They received brief training on typical non-MS vascular enhancements visible on post-contrast 3DT1TSE brain examinations, using examples external to the study sample.

The reading of each batch was separated by one month. Reader 1 started with Batch A, and Reader 2 started with Batch B. For each patient, in each batch the reader had access to only two sequences: a FLAIR sequence and one of the two post-contrast weighted images. For the second reading, they swapped batches, reviewing the FLAIR sequence along with the other post-contrast image.

Readings were performed using the RadiAnt DICOM Viewer software. The readers annotated a spreadsheet with the batch-level patient identifier and the number of enhancing lesions per patient, thought to correspond to enhancing MS lesions. They also saved screen captures of each enhancing lesion to facilitate cross-identification between batches and readers.

#### SIGNAL AND CONTRAST TO NOISE RATIO CALCULTION

We performed post-hoc signal to noise ratio (SNR) and contrast to noise ratio (CNR) analysis on patients with enhancing lesions identified in both sequences. We placed elliptical ROIs in (1) an enhancing lesion, (2) non-enhancing white matter and (3) air (background) on both 3DT1TSE and 3DT1TFE images, and annotated mean enhancing lesion intensity ( $S_{lesion}$ ), mean adjacent non-enhancing white-matter intensity ($S_{white-matter}$) and background noise as the standard deviation of signal in the background air ($\sigma_{air}$). The ROIs of the enhancing lesions were drawn including only voxels with clear enhancement, avoiding lesion-margin or partial volume effects. The ROIs of the adjacent white matter were drawn on non-enhancing white matter adjacent to the enhancing lesions.

We then calculated the SNR as the mean signal of the lesion ROI divided by the standard deviation of the air ROI $SNR = S_{lesion}/\sigma_{air}$, and the CNR as the absolute difference between lesion and non-enhancing white matter mean signals divided by that same standard deviation of the air ROI $CNR = (|S_{lesion} - S_{white-matter}|)/\sigma_{air}$.

#### LESION 005_01

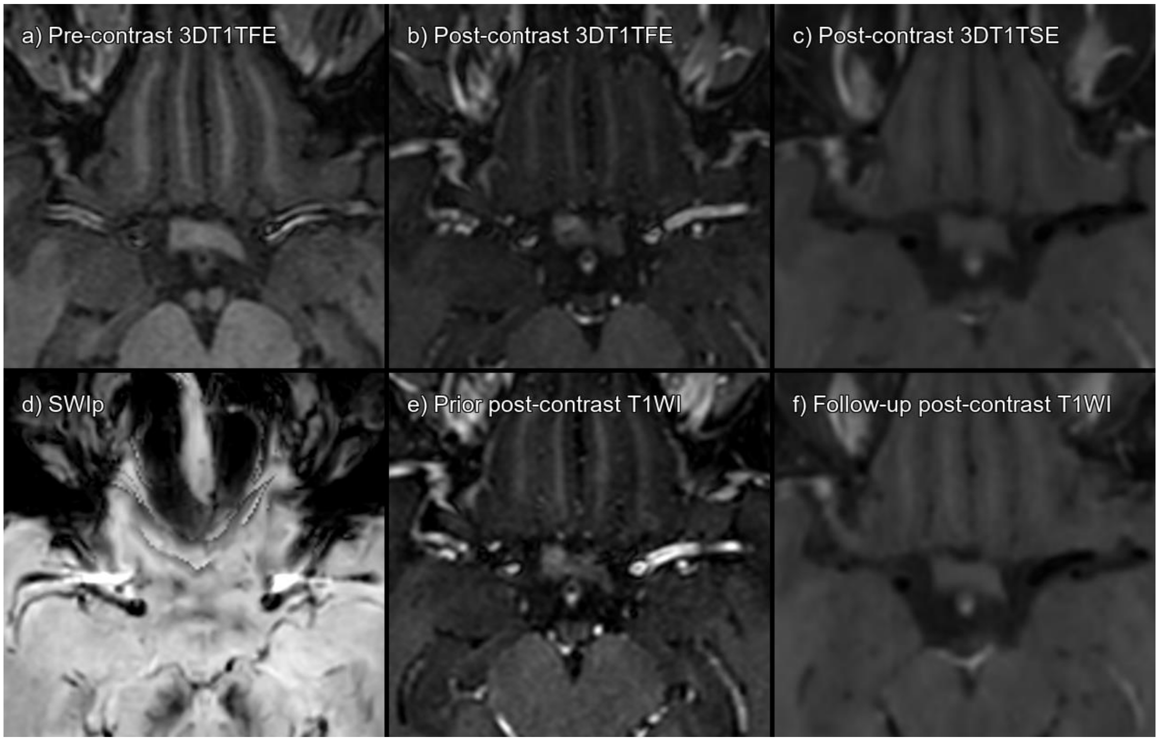


Supplementary figure 1. Lesion 005_01 a) 3DT1TFE without contrast, b) 3DT1TFE post-contrast, c) 3DT1TSE post-contrast, d) SWIp, e) Prior post-contrast T1-weighted imaging 5 months pre-baseline, f) Follow-up post-contrast T1-weighted imaging 4 months post-baseline.

Enhancing prechiasmatic right optic nerve, visible on 3DT1TFE, absent on 3DT1TSE post-contrast. Note focal hypointensity of lesion on SWIp. Note that the lesion was already present on prior 3DT1TFE postcontrast and absent on follow-up 3DT1TSE post-contrast. Consensus established false positive enhancement, possible capillary telangiectasia.

| **CONSENSUS** | **Not acute MS lesion enhancement** | | | |
|---|---|---|---|---|
| | **TFE sequence** | | **TSE sequence** | |
| Visible in sequence | Yes | | No | |
| | **Reader 1** | **Reader 2** | **Reader 1** | **Reader 2** |
| **Detected** | No | Yes | No | No |

Abbreviations: Multiple sclerosis (MS), Turbo Field Echo (TFE), Turbo Spin Echo (TSE), Susceptibility Weighted Imaging with phase enhancement (SWIp).

#### LESION 008_01

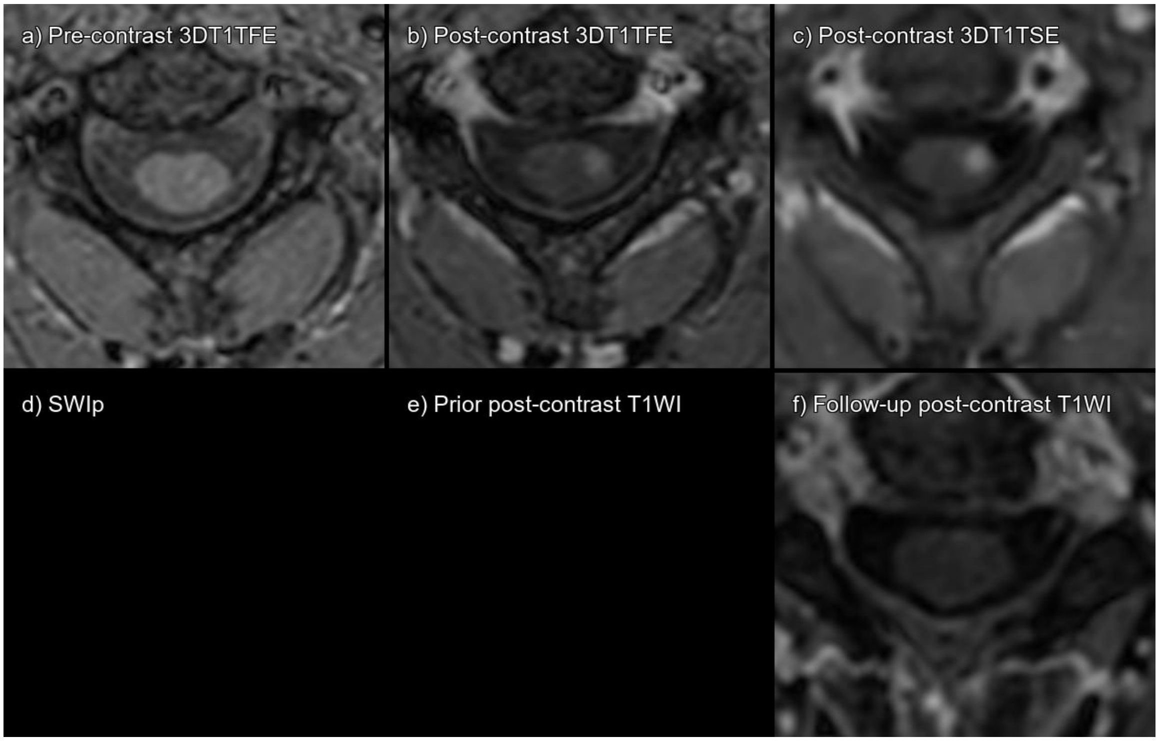


Supplementary figure 2. Lesion 008_01 a) 3DT1TFE without contrast, b) 3DT1TFE post-contrast, c) 3DT1TSE post-contrast, d) SWIp stack does not include this level, e) Prior post-contrast T1-weighted imaging not available, f) Follow-up post-contrast T1-weighted imaging 4 months post-baseline.

True enhancing spinal cord lesion that has disappeared on follow-up MRI.

| **CONSENSUS** | **True Enhancing MS Lesion** | | | |
|---|---|---|---|---|
| | **TFE sequence** | | **TSE sequence** | |
| Visible in sequence | Yes | | Yes | |
| | **Reader 1** | **Reader 2** | **Reader 1** | **Reader 2** |
| **Detected** | Yes | No | Yes | No |

Abbreviations: Multiple sclerosis (MS), Turbo Field Echo (TFE), Turbo Spin Echo (TSE), Susceptibility Weighted Imaging with phase enhancement (SWIp).

#### LESION 011_01

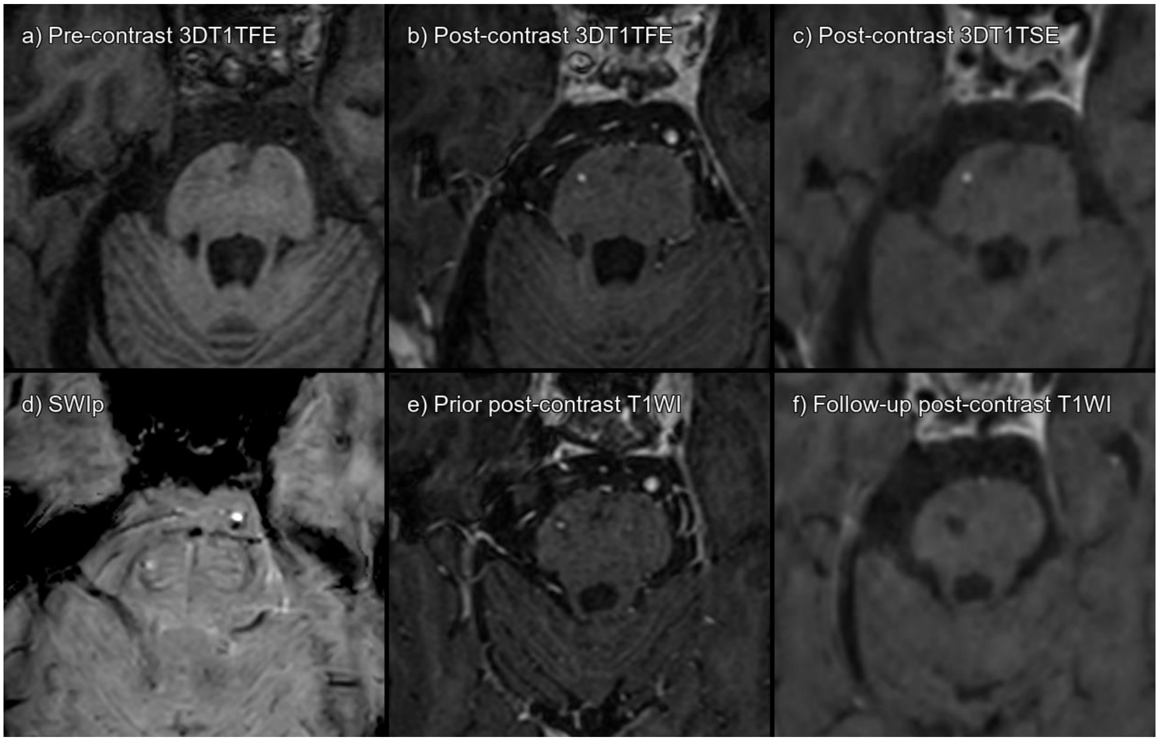


Supplementary figure 3. Lesion 011_01 a) 3DT1TFE without contrast, b) 3DT1TFE post-contrast, c) 3DT1TSE post-contrast, d) SWIp, e) Prior post-contrast T1-weighted imaging 14 months pre-baseline, f) Follow-up post-contrast T1-weighted imaging 13 months post-baseline.

Dot-like enhancing lesion on right aspect of the pons, both in post-contrast 3DT1TFE and 3DT1SE. On SWIp a spot-like hypointensity is found in same location. On previous 3DT1TFE, there was already enhancement. However, it is no longer visible on follow-up 3DT1TSE. Consensus established false positive enhancement on both 3DT1TFE and 3DT1TSE, possible cavernoma or vascular enhancement.

| **CONSENSUS** | **Not acute MS lesion enhancement** | | | |
|---|---|---|---|---|
| | **TFE sequence** | | **TSE sequence** | |
| Visible in sequence | Yes | | Yes | |
| | **Reader 1** | **Reader 2** | **Reader 1** | **Reader 2** |
| **Detected** | No | No | Yes | No |

Abbreviations: Multiple sclerosis (MS), Turbo Field Echo (TFE), Turbo Spin Echo (TSE), Susceptibility Weighted Imaging with phase enhancement (SWIp).

#### LESION 020_01

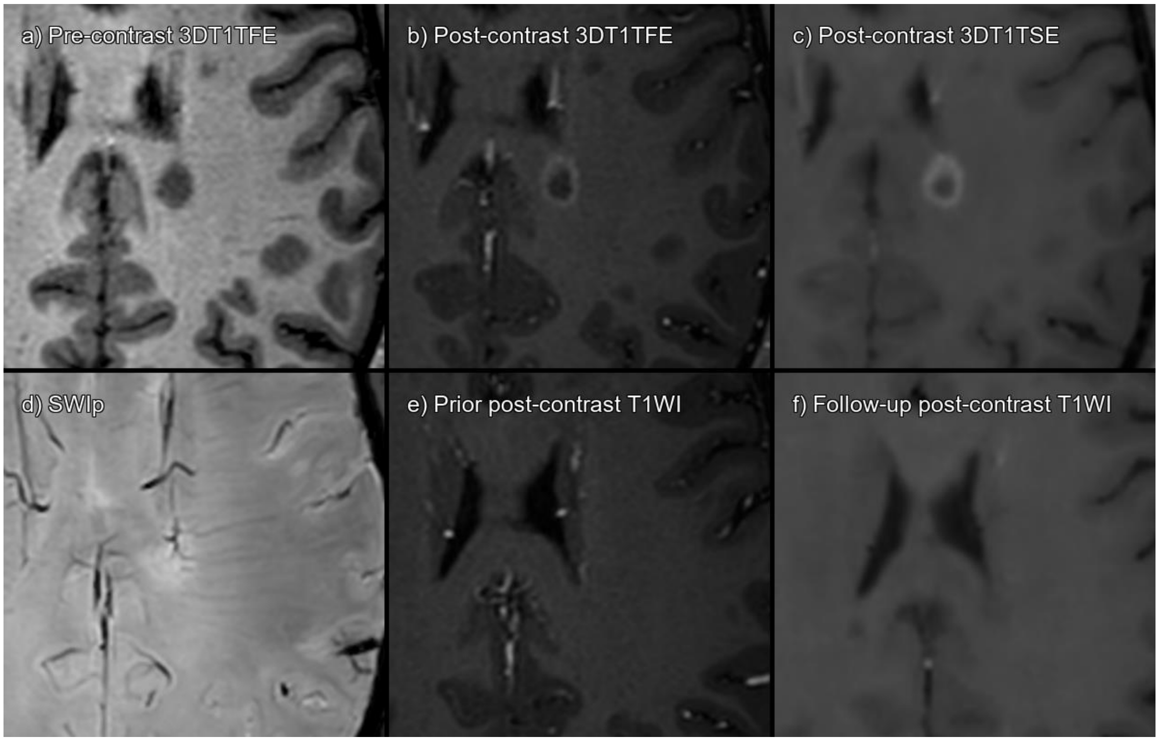


Supplementary figure 4. Lesion 020_01 a) 3DT1TFE without contrast, b) 3DT1TFE post-contrast, c) 3DT1TSE post-contrast, d) SWIp, e) Prior post-contrast T1-weighted imaging 13 months pre-baseline, f) Follow-up post-contrast T1-weighted imaging 5 months post-baseline.

True ring-enhancing periventricular lesion that has disappeared on follow-up MRI.

| **CONSENSUS** | **True Enhancing MS Lesion** | | | |
|---|---|---|---|---|
| | **TFE sequence** | | **TSE sequence** | |
| Visible in sequence | Yes | | Yes | |
| | **Reader 1** | **Reader 2** | **Reader 1** | **Reader 2** |
| **Detected** | Yes | Yes | Yes | Yes |

Abbreviations: Multiple sclerosis (MS), Turbo Field Echo (TFE), Turbo Spin Echo (TSE), Susceptibility Weighted Imaging with phase enhancement (SWIp).

#### LESION 027_01

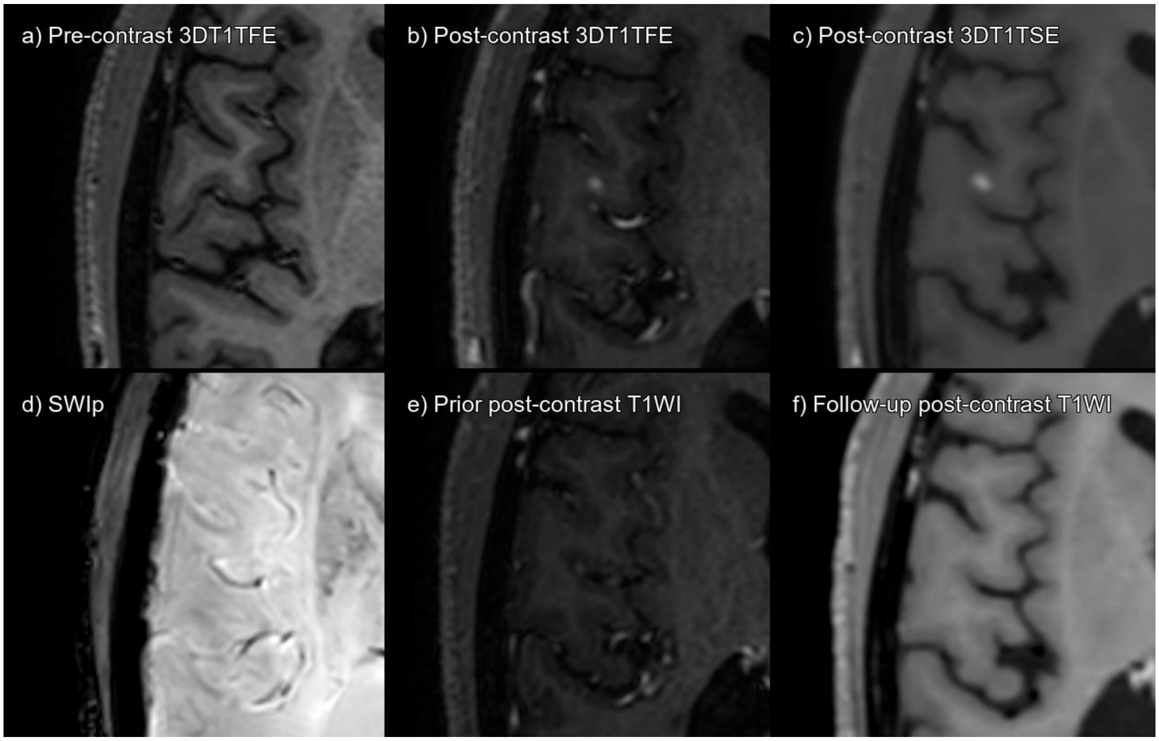


Supplementary figure 5. Lesion 027_01 a) 3DT1TFE without contrast, b) 3DT1TFE post-contrast, c) 3DT1TSE post-contrast, d) SWIp, e) Prior post-contrast T1-weighted imaging 12 months pre-baseline, f) Follow-up post-contrast T1-weighted imaging 6 months post-baseline.

True enhancing subcortical lesion that has disappeared on follow-up MRI.

| **CONSENSUS** | **True Enhancing MS Lesion** | | | |
|---|---|---|---|---|
| | **TFE sequence** | | **TSE sequence** | |
| Visible in sequence | Yes | | Yes | |
| | **Reader 1** | **Reader 2** | **Reader 1** | **Reader 2** |
| **Detected** | No | No | Yes | Yes |

Abbreviations: Multiple sclerosis (MS), Turbo Field Echo (TFE), Turbo Spin Echo (TSE), Susceptibility Weighted Imaging with phase enhancement (SWIp).

#### LESION 029_01

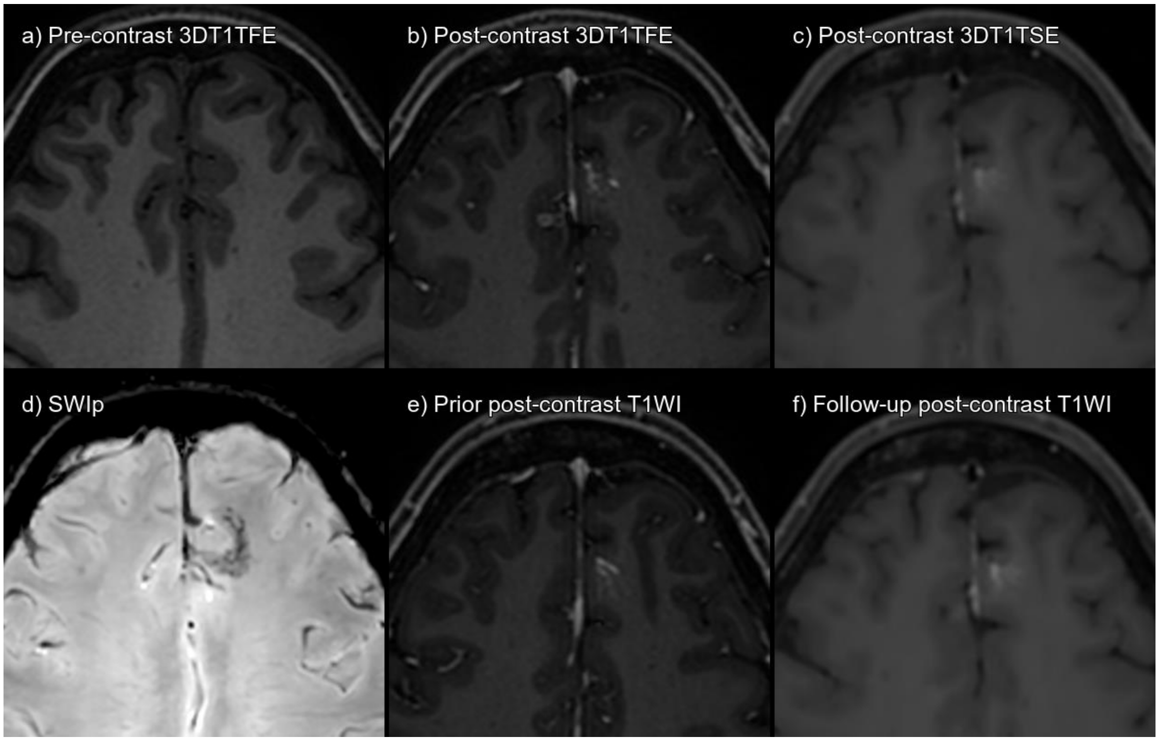


Supplementary figure 6. Lesion 029_01 a) 3DT1TFE without contrast, b) 3DT1TFE post-contrast, c) 3DT1TSE post-contrast, d) SWIp, e) Prior post-contrast T1-weighted imaging 15 months pre-baseline, f) Follow-up post-contrast T1-weighted imaging 13 months post-baseline.

Developmental venous anomaly, evident on SWIp and on 3DT1TFE post-contrast. On 3DT1TSE the vascular structures are not so easily defined. Consensus established the lesion as a false positive, corresponding to a developmental venous anomaly.

| **CONSENSUS** | **Not acute MS lesion enhancement** | | | |
|---|---|---|---|---|
| | **TFE sequence** | | **TSE sequence** | |
| Visible in sequence | Yes | | Yes | |
| | **Reader 1** | **Reader 2** | **Reader 1** | **Reader 2** |
| **Detected** | No | No | No | Yes |

Abbreviations: Multiple sclerosis (MS), Turbo Field Echo (TFE), Turbo Spin Echo (TSE), Susceptibility Weighted Imaging with phase enhancement (SWIp).

#### LESION 029_02

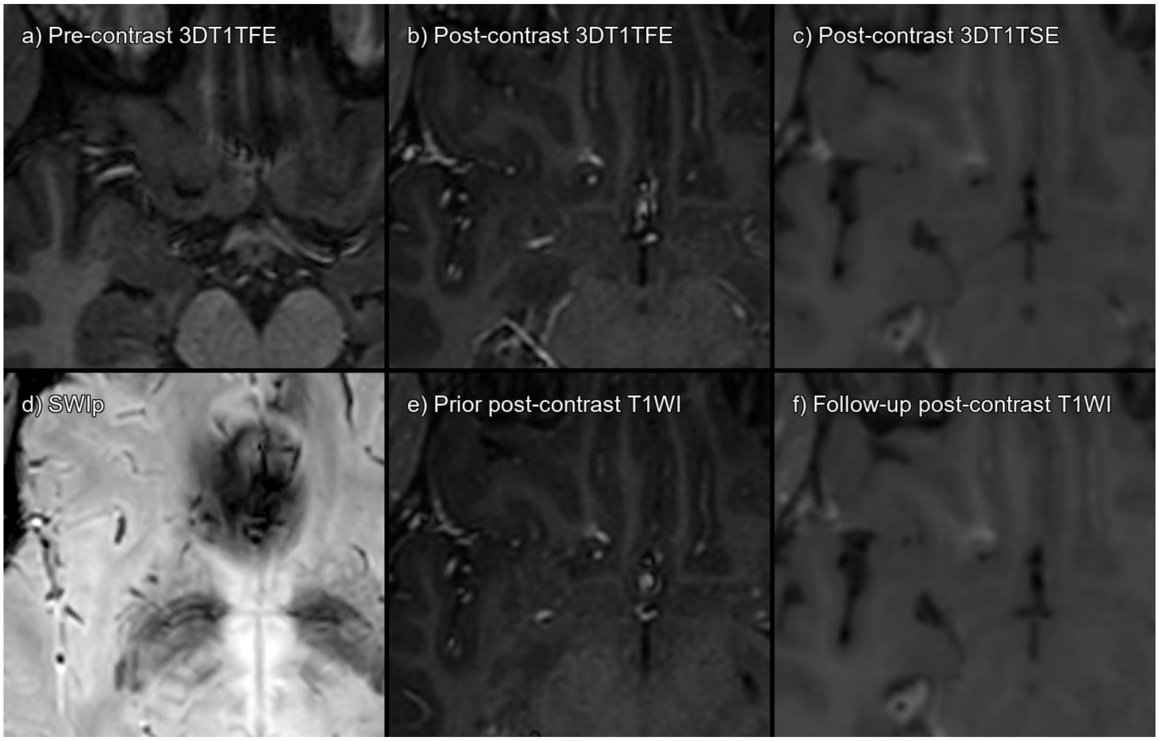


Supplementary figure 7. Lesion 029_02 a) 3DT1TFE without contrast, b) 3DT1TFE post-contrast, c) 3DT1TSE post-contrast, d) SWIp, e) Prior post-contrast T1-weighted imaging 15 months pre-baseline, f) Follow-up post-contrast T1-weighted imaging 13 months post-baseline.

Developmental venous anomaly, evident on SWIp and on 3DT1TFE post-contrast. On 3DT1TSE the vascular structures are not so easily defined. Consensus established the lesion as a false positive, corresponding to a developmental venous anomaly.

| **CONSENSUS** | **Not acute MS lesion enhancement** | | | |
|---|---|---|---|---|
| | **TFE sequence** | | **TSE sequence** | |
| Visible in sequence | Yes | | Yes | |
| | **Reader 1** | **Reader 2** | **Reader 1** | **Reader 2** |
| **Detected** | No | No | No | Yes |

Abbreviations: Multiple sclerosis (MS), Turbo Field Echo (TFE), Turbo Spin Echo (TSE), Susceptibility Weighted Imaging with phase enhancement (SWIp).

#### LESION 036_01

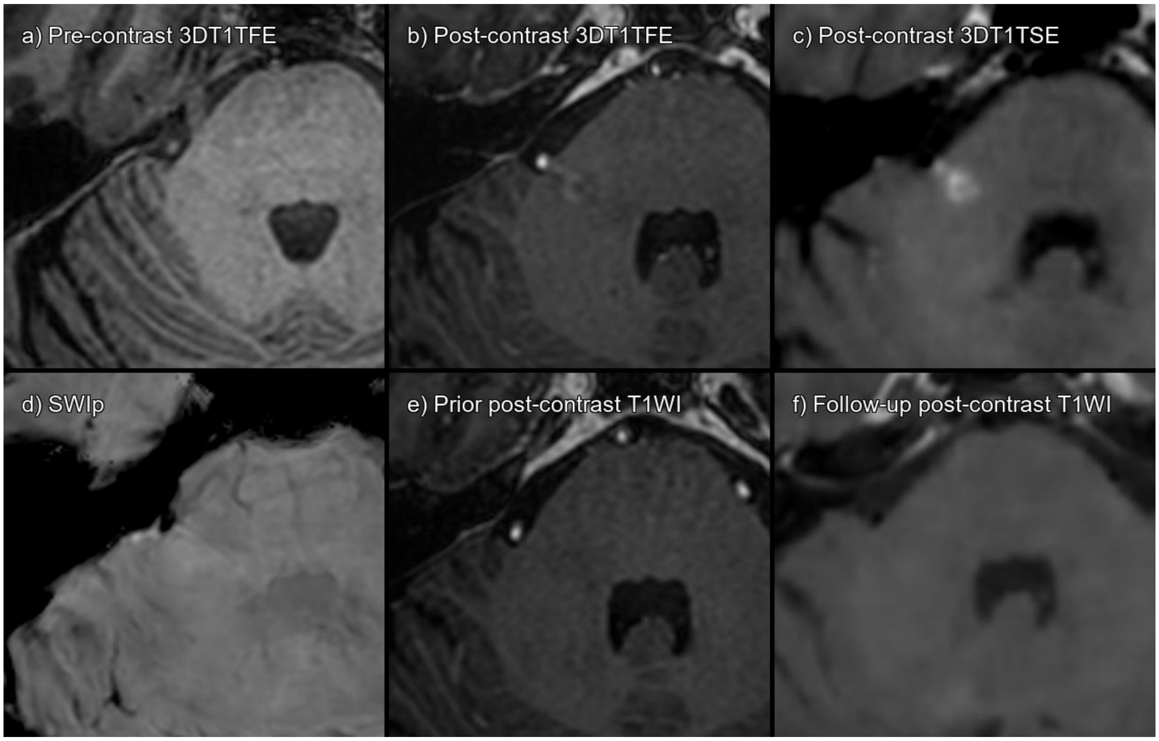


Supplementary figure 8. Lesion 036_01 a) 3DT1TFE without contrast, b) 3DT1TFE post-contrast, c) 3DT1TSE post-contrast, d) SWIp, e) Prior post-contrast T1-weighted imaging 4 years pre-baseline, f) Follow-up post-contrast T1-weighted imaging 6 months post-baseline.

True enhancing infratentorial lesion that has disappeared on follow-up MRI.

| **CONSENSUS** | **True Enhancing MS Lesion** | | | |
|---|---|---|---|---|
| | **TFE sequence** | | **TSE sequence** | |
| Visible in sequence | Yes | | Yes | |
| | **Reader 1** | **Reader 2** | **Reader 1** | **Reader 2** |
| **Detected** | Yes | No | Yes | Yes |

Abbreviations: Multiple sclerosis (MS), Turbo Field Echo (TFE), Turbo Spin Echo (TSE), Susceptibility Weighted Imaging with phase enhancement (SWIp).

#### LESION 056_01

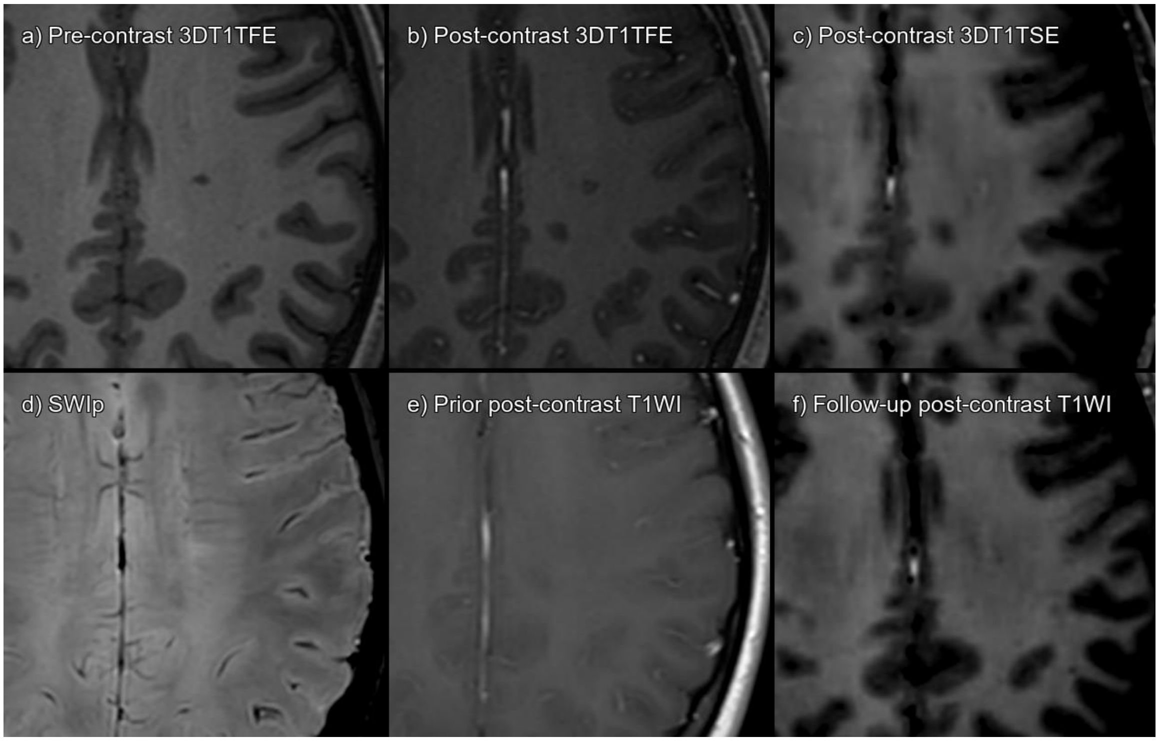


Supplementary figure 9. Lesion 056_01 a) 3DT1TFE without contrast, b) 3DT1TFE post-contrast, c) 3DT1TSE post-contrast, d) SWIp, e) Prior post-contrast T1-weighted imaging 6 months pre-baseline, f) Follow-up post-contrast T1-weighted imaging 6 months post-baseline.

True enhancing white-matter lesion that has disappeared on follow-up MRI. Subtle on 3DT1TSE, not visible on 3DT1TFE.

| **CONSENSUS** | **True Enhancing MS Lesion** | | | |
|---|---|---|---|---|
| | **TFE sequence** | | **TSE sequence** | |
| Visible in sequence | No | | Yes | |
| | **Reader 1** | **Reader 2** | **Reader 1** | **Reader 2** |
| **Detected** | No | No | No | Yes |

Abbreviations: Multiple sclerosis (MS), Turbo Field Echo (TFE), Turbo Spin Echo (TSE), Susceptibility Weighted Imaging with phase enhancement (SWIp).

#### LESION 069_01

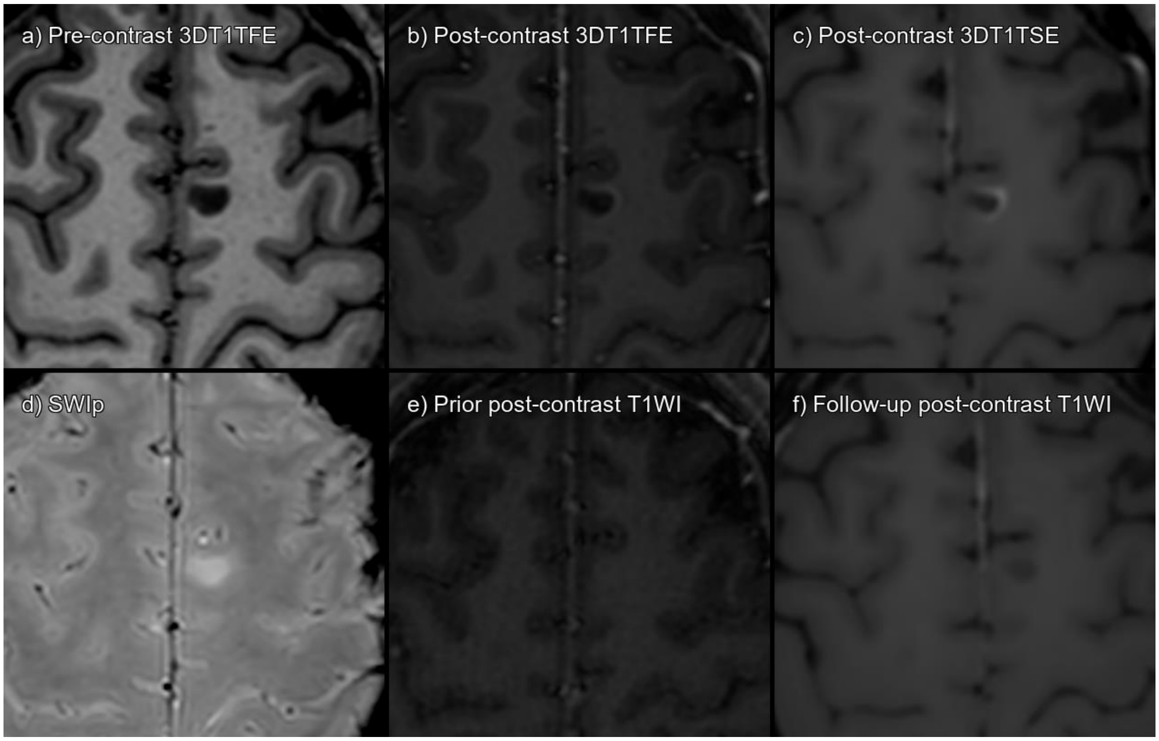


Supplementary figure 10. Lesion 069_01 a) 3DT1TFE without contrast, b) 3DT1TFE post-contrast, c) 3DT1TSE post-contrast, d) SWIp, e) Prior post-contrast T1-weighted imaging 7 months pre-baseline, f) Follow-up post-contrast T1-weighted imaging 8 months post-baseline.

True ring-enhancing white-matter lesion that has disappeared on follow-up MRI.

| **CONSENSUS** | **True Enhancing MS Lesion** | | | |
|---|---|---|---|---|
| | **TFE sequence** | | **TSE sequence** | |
| Visible in sequence | Yes | | Yes | |
| | **Reader 1** | **Reader 2** | **Reader 1** | **Reader 2** |
| **Detected** | Yes | Yes | Yes | Yes |

Abbreviations: Multiple sclerosis (MS), Turbo Field Echo (TFE), Turbo Spin Echo (TSE), Susceptibility Weighted Imaging with phase enhancement (SWIp).

#### LESION 080_01

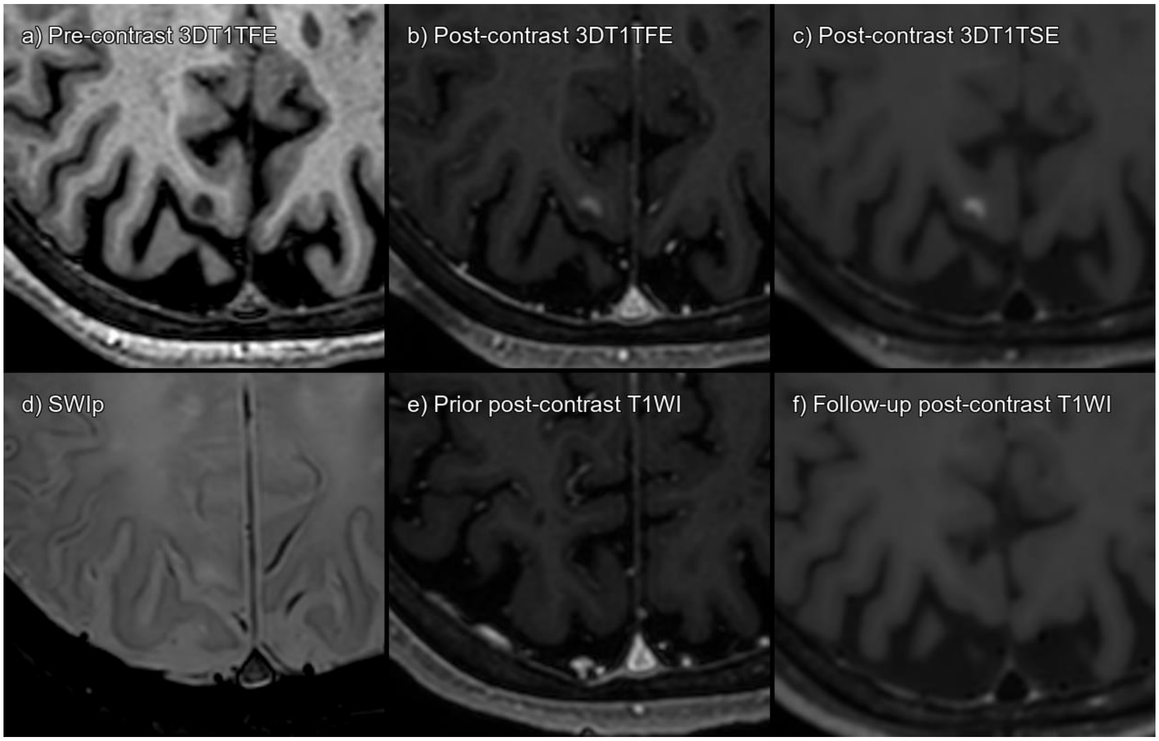


Supplementary figure 11. Lesion 080_01 a) 3DT1TFE without contrast, b) 3DT1TFE post-contrast, c) 3DT1TSE post-contrast, d) SWIp, e) Prior post-contrast T1-weighted imaging 12 months pre-baseline, f) Follow-up post-contrast T1-weighted imaging 6 months post-baseline.

True enhancing subcortical lesion that has disappeared on follow-up MRI.

| **CONSENSUS** | **True Enhancing MS Lesion** | | | |
|---|---|---|---|---|
| | **TFE sequence** | | **TSE sequence** | |
| Visible in sequence | Yes | | Yes | |
| | **Reader 1** | **Reader 2** | **Reader 1** | **Reader 2** |
| **Detected** | No | Yes | Yes | Yes |

Abbreviations: Multiple sclerosis (MS), Turbo Field Echo (TFE), Turbo Spin Echo (TSE), Susceptibility Weighted Imaging with phase enhancement (SWIp).

#### LESION 083_01

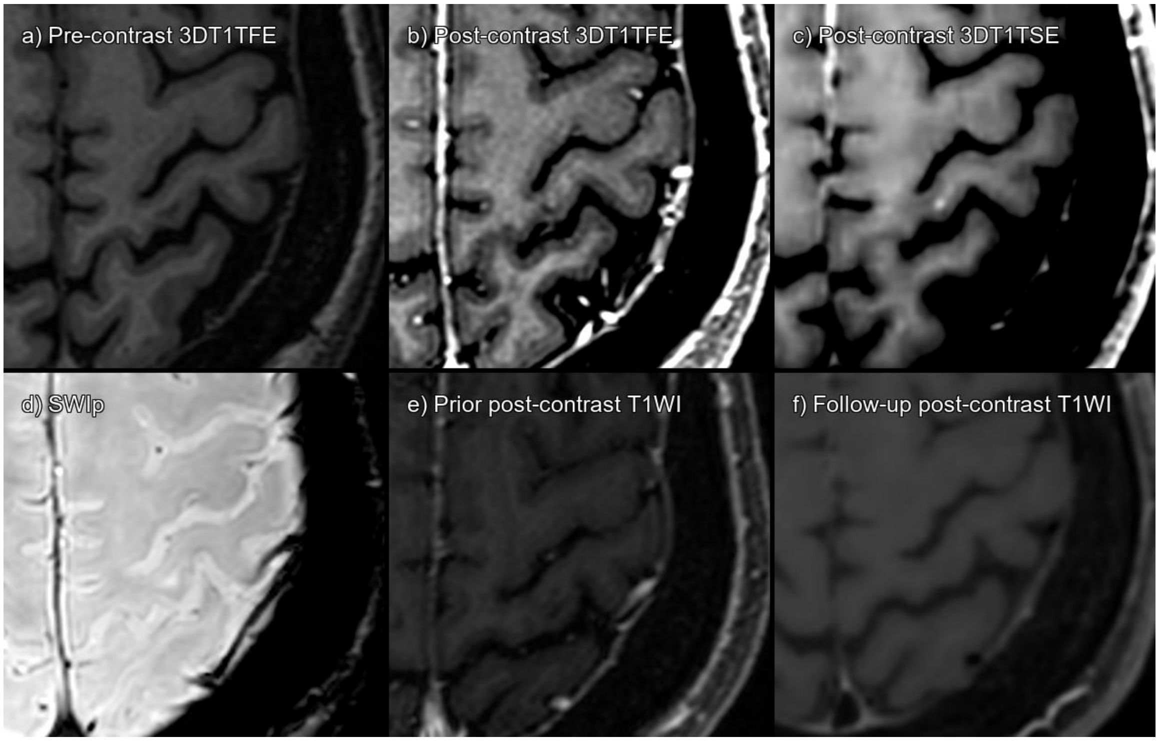


Supplementary figure 12. Lesion 083_01 a) 3DT1TFE without contrast, b) 3DT1TFE post-contrast, c) 3DT1TSE post-contrast, d) SWIp, e) Prior post-contrast T1-weighted imaging 4 months pre-baseline, f) Follow-up post-contrast T1-weighted imaging 3 months post-baseline.

True enhancing subcortical lesion, visible on 3DT1TSE and not on 3DT1TFE, that has disappeared on follow-up MRI.

| **CONSENSUS** | **True Enhancing MS Lesion** | | | |
|---|---|---|---|---|
| | **TFE sequence** | | **TSE sequence** | |
| Visible in sequence | No | | Yes | |
| | **Reader 1** | **Reader 2** | **Reader 1** | **Reader 2** |
| **Detected** | No | No | No | Yes |

Abbreviations: Multiple sclerosis (MS), Turbo Field Echo (TFE), Turbo Spin Echo (TSE), Susceptibility Weighted Imaging with phase enhancement (SWIp).

#### LESION 085_01

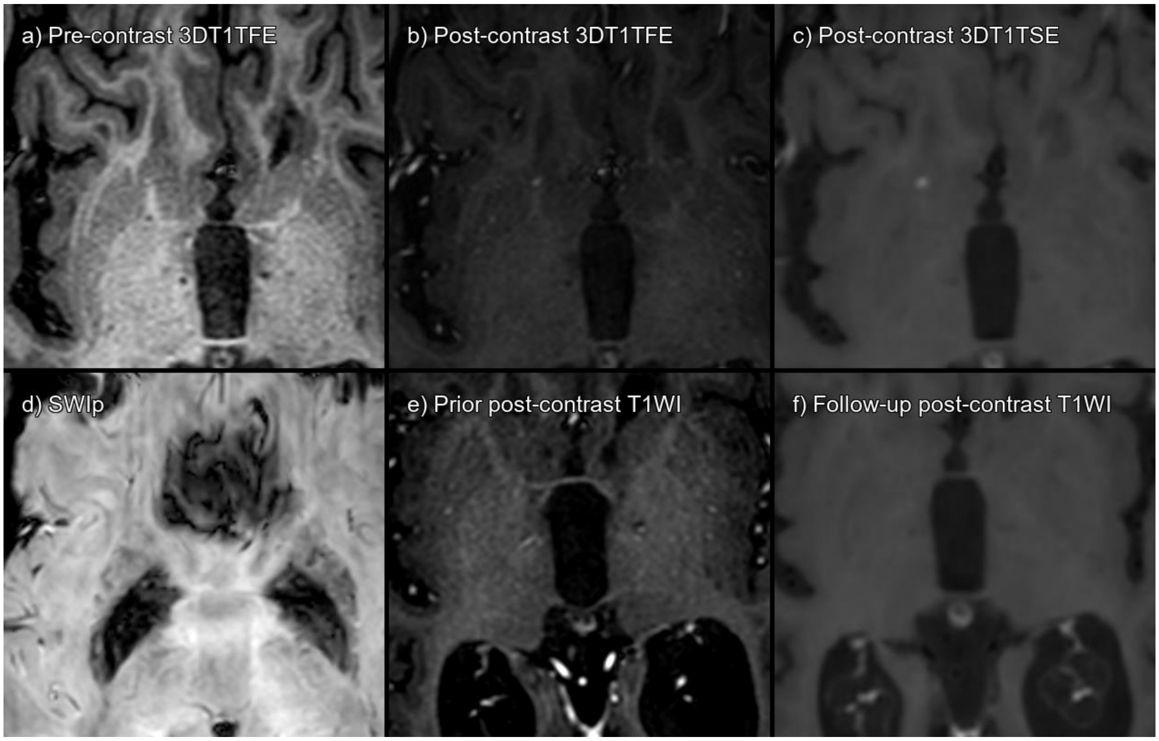


Supplementary figure 13. Lesion 085_01 a) 3DT1TFE without contrast, b) 3DT1TFE post-contrast, c) 3DT1TSE post-contrast, d) SWIp, e) Prior post-contrast T1-weighted imaging 9 months pre-baseline, f) Follow-up post-contrast T1-weighted imaging 10 months post-baseline.

True enhancing deep tissue lesion, that has disappeared on follow-up MRI.

| **CONSENSUS** | **True Enhancing MS Lesion** | | | |
|---|---|---|---|---|
| | **TFE sequence** | | **TSE sequence** | |
| Visible in sequence | Yes | | Yes | |
| | **Reader 1** | **Reader 2** | **Reader 1** | **Reader 2** |
| **Detected** | No | No | No | Yes |

Abbreviations: Multiple sclerosis (MS), Turbo Field Echo (TFE), Turbo Spin Echo (TSE), Susceptibility Weighted Imaging with phase enhancement (SWIp).

#### LESION 086_01

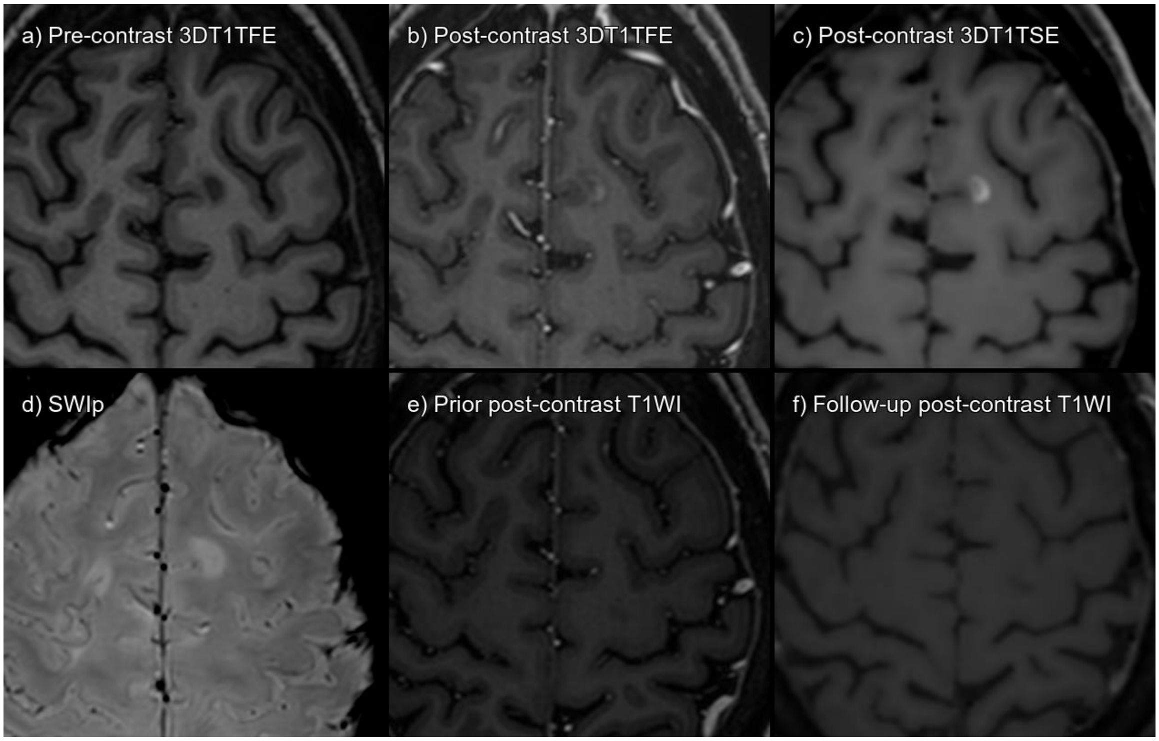


Supplementary figure 14. Lesion 086_01 a) 3DT1TFE without contrast, b) 3DT1TFE post-contrast, c) 3DT1TSE post-contrast, d) SWIp, e) Prior post-contrast T1-weighted imaging 3 years pre-baseline, f) Follow-up post-contrast T1-weighted imaging 7 months post-baseline.

True enhancing subcortical lesion, that has disappeared on follow-up MRI.

| **CONSENSUS** | **True Enhancing MS Lesion** | | | |
|---|---|---|---|---|
| | **TFE sequence** | | **TSE sequence** | |
| Visible in sequence | Yes | | Yes | |
| | **Reader 1** | **Reader 2** | **Reader 1** | **Reader 2** |
| **Detected** | No | Yes | Yes | Yes |

Abbreviations: Multiple sclerosis (MS), Turbo Field Echo (TFE), Turbo Spin Echo (TSE), Susceptibility Weighted Imaging with phase enhancement (SWIp).

#### LESION 088_01

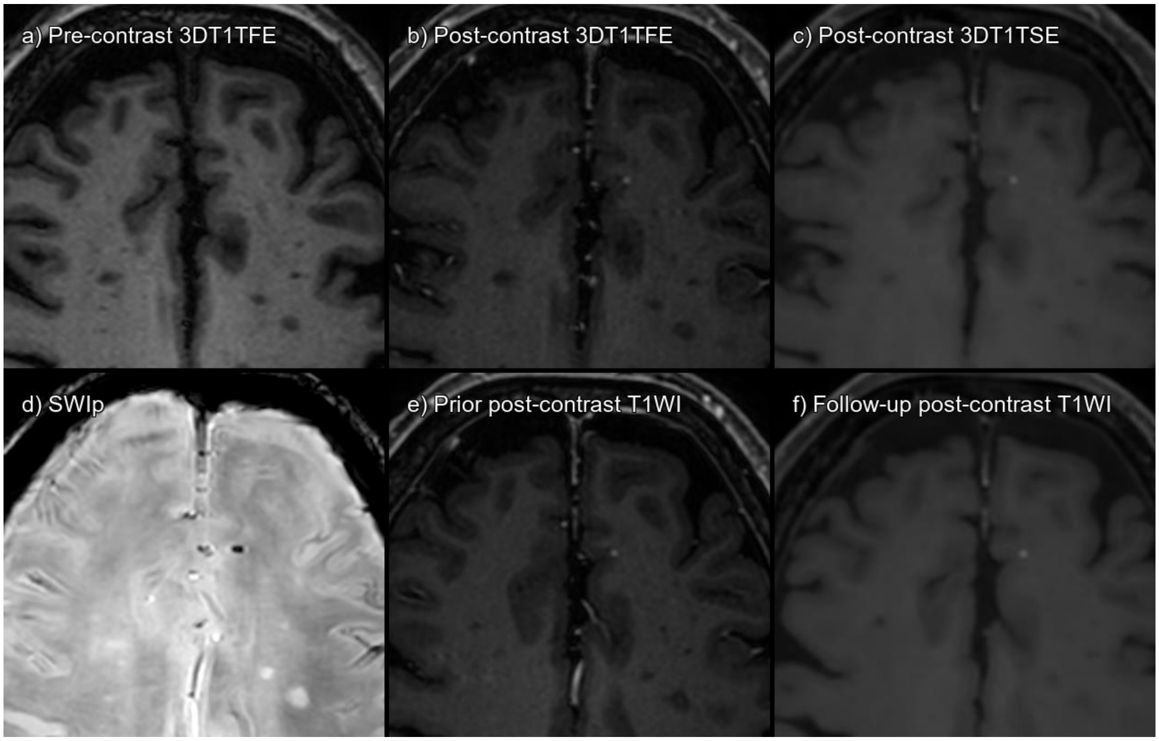


Supplementary figure 15. Lesion 088_01 a) 3DT1TFE without contrast, b) 3DT1TFE post-contrast, c) 3DT1TSE post-contrast, d) SWIp, e) Prior post-contrast T1-weighted imaging 4 months pre-baseline, f) Follow-up post-contrast T1-weighted imaging 7 months post-baseline.

Dot-like juxtacortical enhancement. Hypointense on SWI. Present on previous and on follow-up examination. Probable cavernoma. Consensus established the lesion as a false positive, probably corresponding to a cavernoma.

| **CONSENSUS** | **Not acute MS lesion enhancement** | | | |
|---|---|---|---|---|
| | **TFE sequence** | | **TSE sequence** | |
| Visible in sequence | Yes | | Yes | |
| | **Reader 1** | **Reader 2** | **Reader 1** | **Reader 2** |
| **Detected** | No | No | Yes | No |

Abbreviations: Multiple sclerosis (MS), Turbo Field Echo (TFE), Turbo Spin Echo (TSE), Susceptibility Weighted Imaging with phase enhancement (SWIp).

#### LESION 093_01

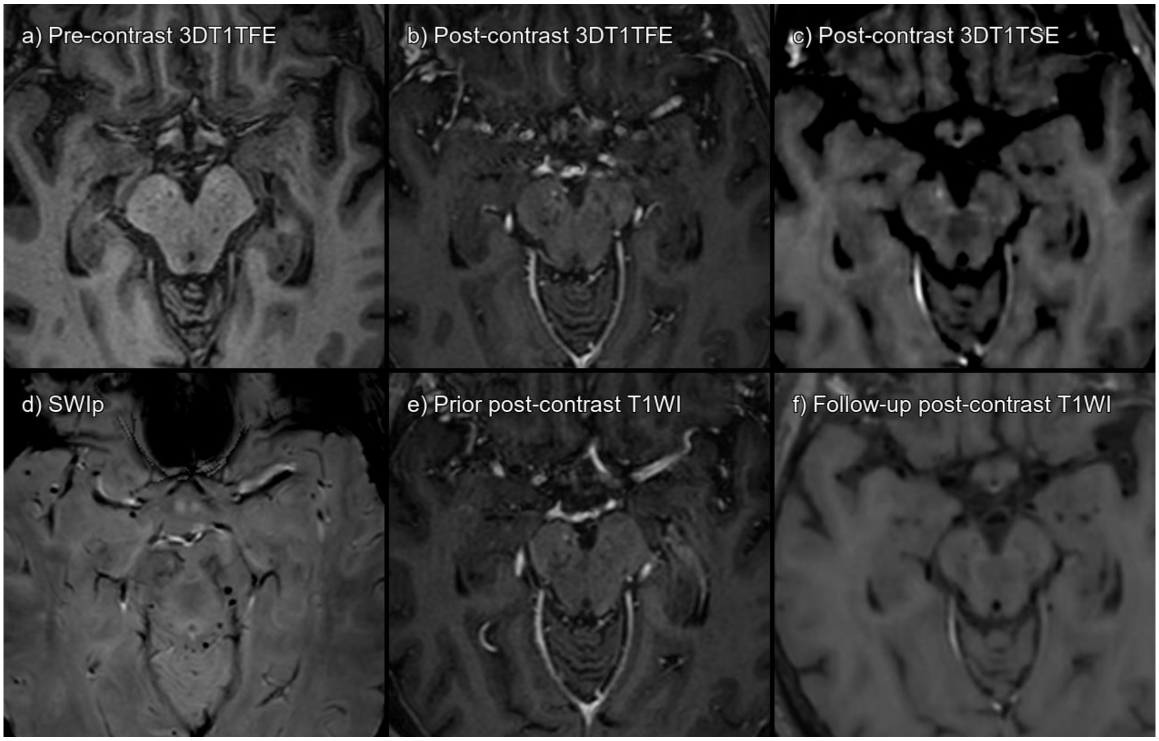


Supplementary figure 16. Lesion 093_01 a) 3DT1TFE without contrast, b) 3DT1TFE post-contrast, c) 3DT1TSE post-contrast, d) SWIp, e) Prior post-contrast T1-weighted imaging 4 months pre-baseline, f) Follow-up post-contrast T1-weighted imaging 7 months post-baseline.

Dot-like right mesencephalic enhancement. Present on previous examination. Absent on follow-up examination. Consensus established the lesion as a false positive, suggestive of a nonspecific probably vascular enhancement.

| **CONSENSUS** | **Not acute MS lesion enhancement** | | | |
|---|---|---|---|---|
| | **TFE sequence** | | **TSE sequence** | |
| Visible in sequence | Yes | | Yes | |
| | **Reader 1** | **Reader 2** | **Reader 1** | **Reader 2** |
| **Detected** | No | No | No | Yes |

Abbreviations: Multiple sclerosis (MS), Turbo Field Echo (TFE), Turbo Spin Echo (TSE), Susceptibility Weighted Imaging with phase enhancement (SWIp).

#### LESION 108_01

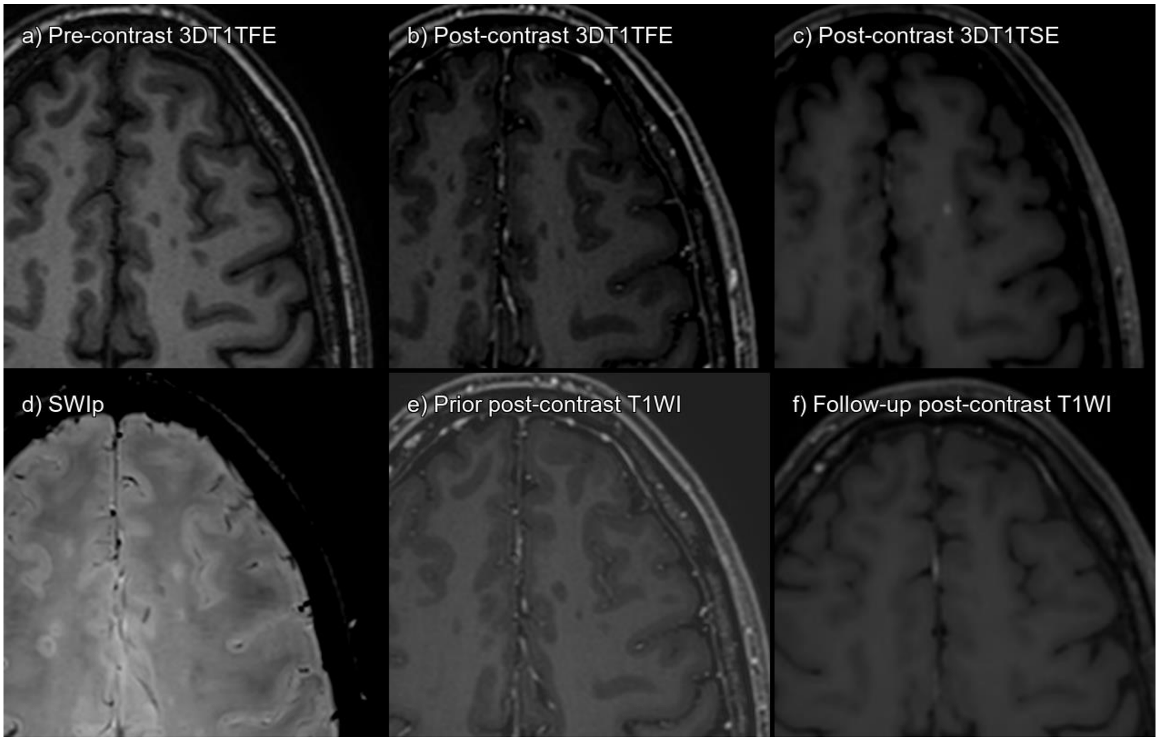


Supplementary figure 17. Lesion 108_01 a) 3DT1TFE without contrast, b) 3DT1TFE post-contrast, c) 3DT1TSE post-contrast, d) SWIp, e) Prior post-contrast T1-weighted imaging 15 months pre-baseline, f) Follow-up post-contrast T1-weighted imaging 5 months post-baseline.

True enhancing subcortical lesion, that has disappeared on follow-up MRI.

| **CONSENSUS** | **True Enhancing MS Lesion** | | | |
|---|---|---|---|---|
| | **TFE sequence** | | **TSE sequence** | |
| Visible in sequence | Yes | | Yes | |
| | **Reader 1** | **Reader 2** | **Reader 1** | **Reader 2** |
| **Detected** | No | No | Yes | Yes |

Abbreviations: Multiple sclerosis (MS), Turbo Field Echo (TFE), Turbo Spin Echo (TSE), Susceptibility Weighted Imaging with phase enhancement (SWIp).

#### LESION 108_02

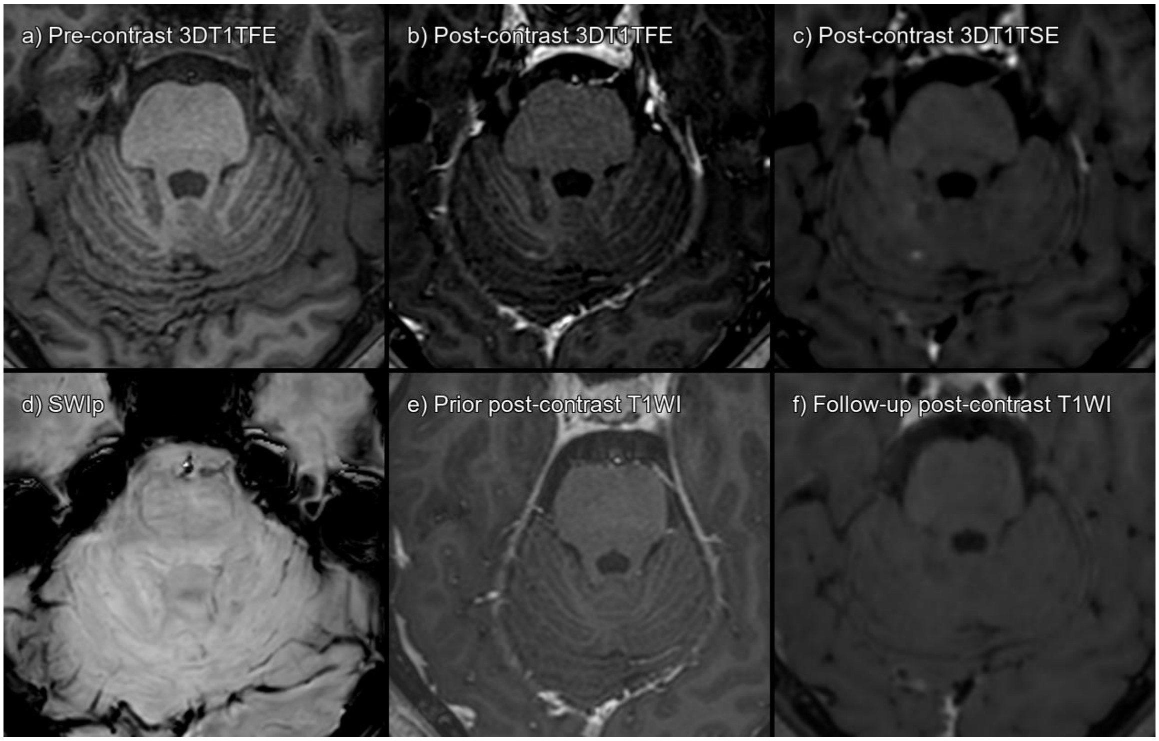


Supplementary figure 18. Lesion 108_02 a) 3DT1TFE without contrast, b) 3DT1TFE post-contrast, c) 3DT1TSE post-contrast, d) SWIp, e) Prior post-contrast T1-weighted imaging 15 months pre-baseline, f) Follow-up post-contrast T1-weighted imaging 5 months post-baseline.

True enhancing infratentorial lesion, that has disappeared on follow-up MRI.

| **CONSENSUS** | **True Enhancing MS Lesion** | | | |
|---|---|---|---|---|
| | **TFE sequence** | | **TSE sequence** | |
| Visible in sequence | Yes | | Yes | |
| | **Reader 1** | **Reader 2** | **Reader 1** | **Reader 2** |
| **Detected** | No | No | Yes | Yes |

Abbreviations: Multiple sclerosis (MS), Turbo Field Echo (TFE), Turbo Spin Echo (TSE), Susceptibility Weighted Imaging with phase enhancement (SWIp).

#### LESION 108_03

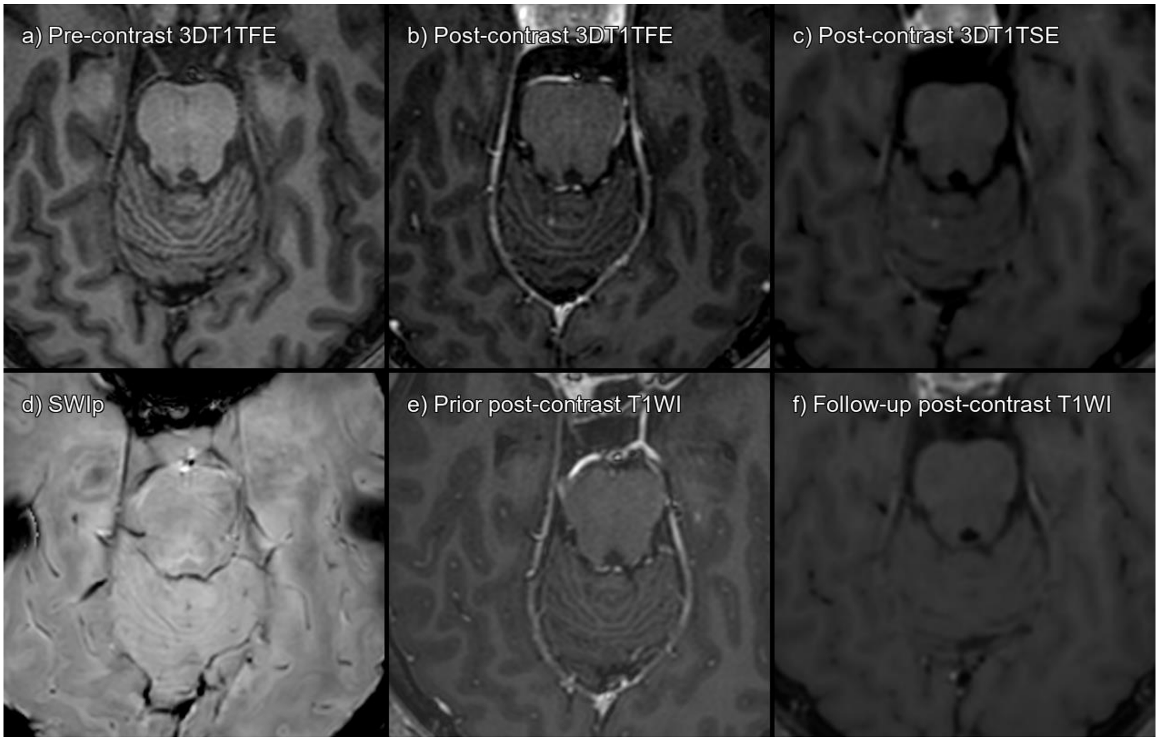


Supplementary figure 19. Lesion 108_03 a) 3DT1TFE without contrast, b) 3DT1TFE post-contrast, c) 3DT1TSE post-contrast, d) SWIp, e) Prior post-contrast T1-weighted imaging 15 months pre-baseline, f) Follow-up post-contrast T1-weighted imaging 5 months post-baseline.

True enhancing infratentorial lesion, that has disappeared on follow-up MRI.

| **CONSENSUS** | **True Enhancing MS Lesion** | | | |
|---|---|---|---|---|
| | **TFE sequence** | | **TSE sequence** | |
| Visible in sequence | Yes | | Yes | |
| | **Reader 1** | **Reader 2** | **Reader 1** | **Reader 2** |
| **Detected** | No | No | Yes | Yes |

Abbreviations: Multiple sclerosis (MS), Turbo Field Echo (TFE), Turbo Spin Echo (TSE), Susceptibility Weighted Imaging with phase enhancement (SWIp).

#### LESION 125_01

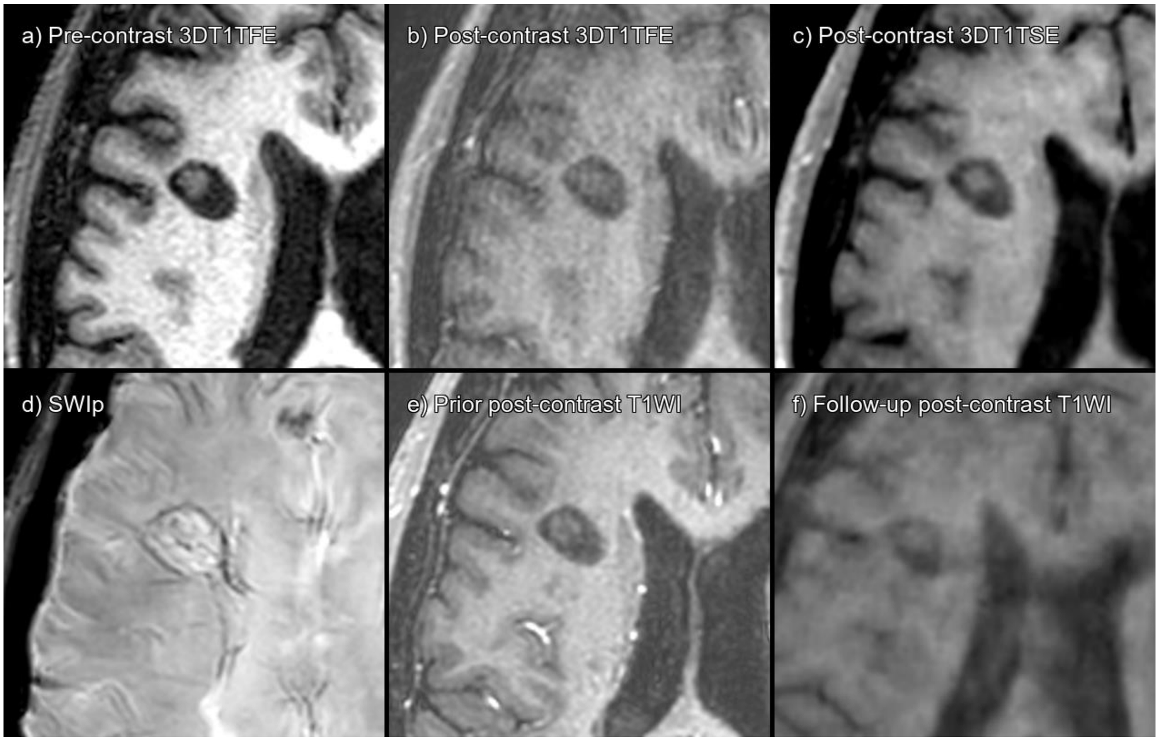


Supplementary figure 20. Lesion 125_01 a) 3DT1TFE without contrast, b) 3DT1TFE post-contrast, c) 3DT1TSE post-contrast, d) SWIp, e) Prior post-contrast T1-weighted imaging 20 months pre-baseline, f) Follow-up post-contrast T1-weighted imaging 6 months post-baseline.

This was not an enhancing lesion, but a lesion that had T1-dark rim and a T1-hyperintense center, as is evident in pre-contrast T1-weighted image. The lesion also has an iron rim on SWIp.

| **CONSENSUS** | **Not acute MS lesion enhancement** | | | |
|---|---|---|---|---|
| | **TFE sequence** | | **TSE sequence** | |
| Visible in sequence | No | | No | |
| | **Reader 1** | **Reader 2** | **Reader 1** | **Reader 2** |
| **Detected** | No | No | No | Yes |

Abbreviations: Multiple sclerosis (MS), Turbo Field Echo (TFE), Turbo Spin Echo (TSE), Susceptibility Weighted Imaging with phase enhancement (SWIp).

#### LESION 146_01

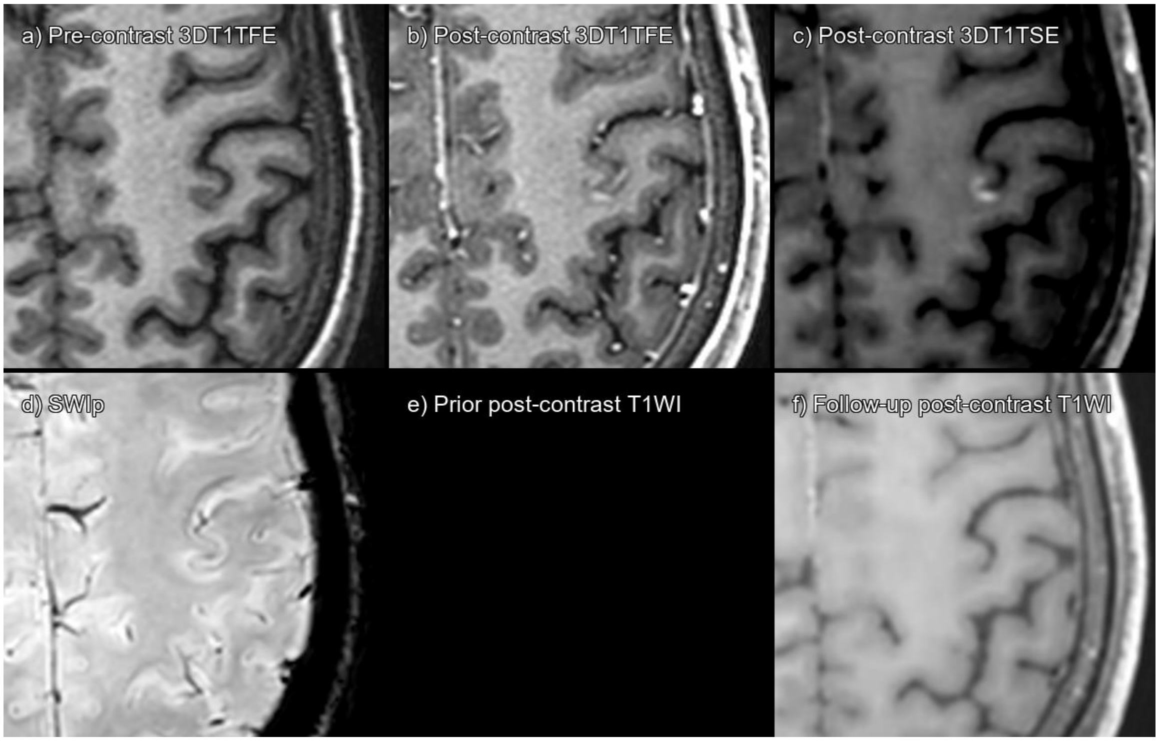


Supplementary figure 21. Lesion 146_01 a) 3DT1TFE without contrast, b) 3DT1TFE post-contrast, c) 3DT1TSE post-contrast, d) SWIp, e) Prior post-contrast T1-weighted imaging not available, f) Follow-up post-contrast T1-weighted imaging 6 months post-baseline.

True enhancing juxtacortical lesion, that has disappeared on follow-up MRI.

| **CONSENSUS** | **True Enhancing MS Lesion** | | | |
|---|---|---|---|---|
| | **TFE sequence** | | **TSE sequence** | |
| Visible in sequence | Yes | | Yes | |
| | **Reader 1** | **Reader 2** | **Reader 1** | **Reader 2** |
| **Detected** | Yes | No | Yes | Yes |

Abbreviations: Multiple sclerosis (MS), Turbo Field Echo (TFE), Turbo Spin Echo (TSE), Susceptibility Weighted Imaging with phase enhancement (SWIp).

#### LESION 146_02

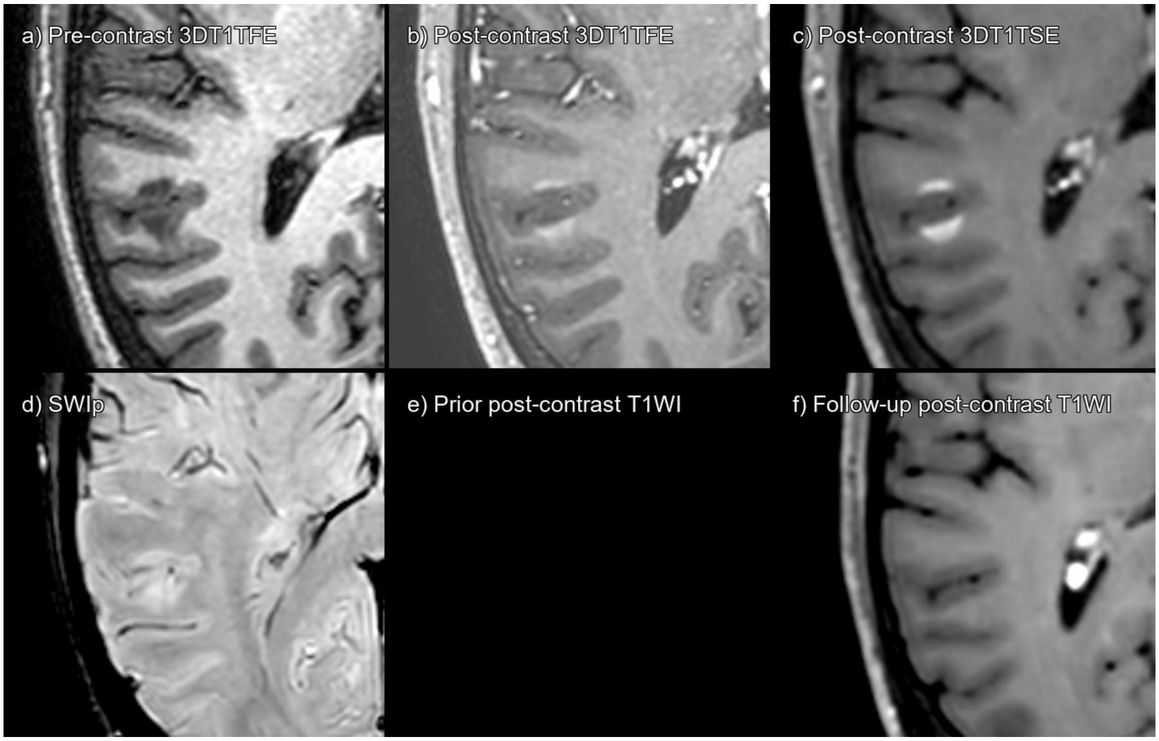


Supplementary figure 22. Lesion 146_02 a) 3DT1TFE without contrast, b) 3DT1TFE post-contrast, c) 3DT1TSE post-contrast, d) SWIp, e) Prior post-contrast T1-weighted imaging not available, f) Follow-up post-contrast T1-weighted imaging 6 months post-baseline.

True enhancing juxtacortical lesion, that has disappeared on follow-up MRI. Lesion 146_02 corresponds to the more anterior lesion visible.

| **CONSENSUS** | **True Enhancing MS Lesion** | | | |
|---|---|---|---|---|
| | **TFE sequence** | | **TSE sequence** | |
| Visible in sequence | Yes | | Yes | |
| | **Reader 1** | **Reader 2** | **Reader 1** | **Reader 2** |
| **Detected** | Yes | No | Yes | Yes |

Abbreviations: Multiple sclerosis (MS), Turbo Field Echo (TFE), Turbo Spin Echo (TSE), Susceptibility Weighted Imaging with phase enhancement (SWIp).

#### LESION 146_03

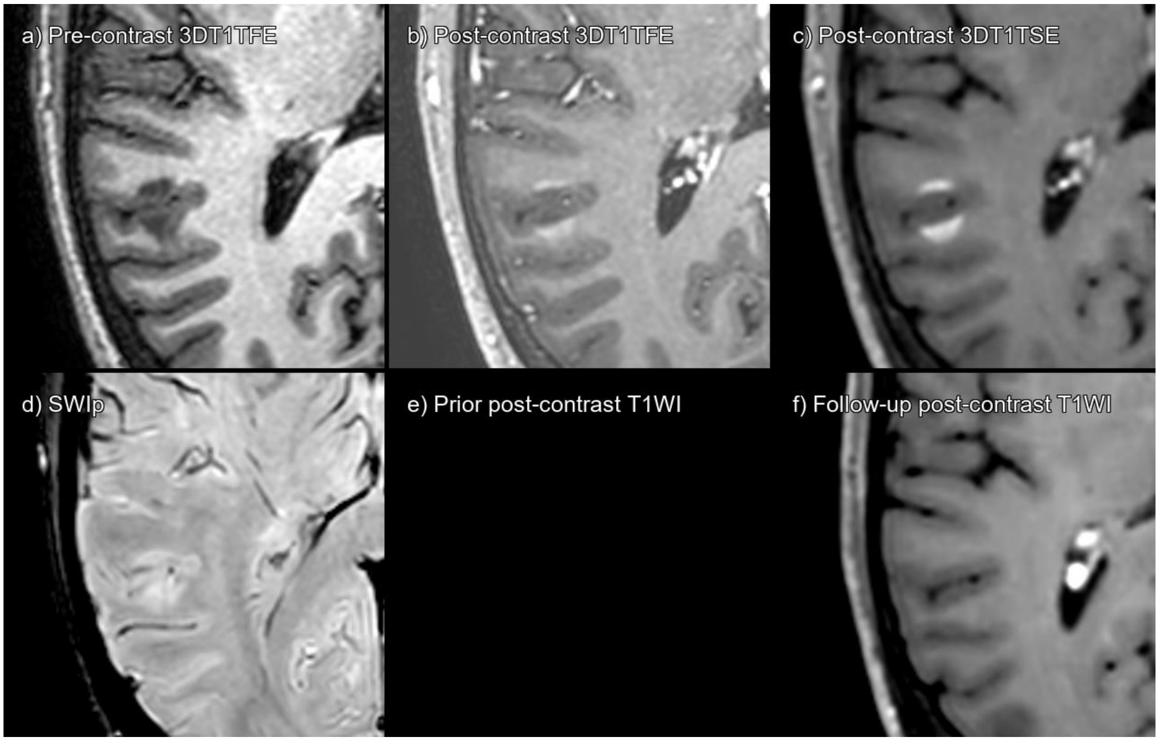


Supplementary figure 23. Lesion 146_03 a) 3DT1TFE without contrast, b) 3DT1TFE post-contrast, c) 3DT1TSE post-contrast, d) SWIp, e) Prior post-contrast T1-weighted imaging not available, f) Follow-up post-contrast T1-weighted imaging 6 months post-baseline.

True enhancing juxtacortical lesion, that has disappeared on follow-up MRI. Lesion 146_03 corresponds to the more posterior lesion visible.

| **CONSENSUS** | **True Enhancing MS Lesion** | | | |
|---|---|---|---|---|
| | **TFE sequence** | | **TSE sequence** | |
| Visible in sequence | Yes | | Yes | |
| | **Reader 1** | **Reader 2** | **Reader 1** | **Reader 2** |
| **Detected** | Yes | No | Yes | Yes |

Abbreviations: Multiple sclerosis (MS), Turbo Field Echo (TFE), Turbo Spin Echo (TSE), Susceptibility Weighted Imaging with phase enhancement (SWIp).

#### LESION 146_04

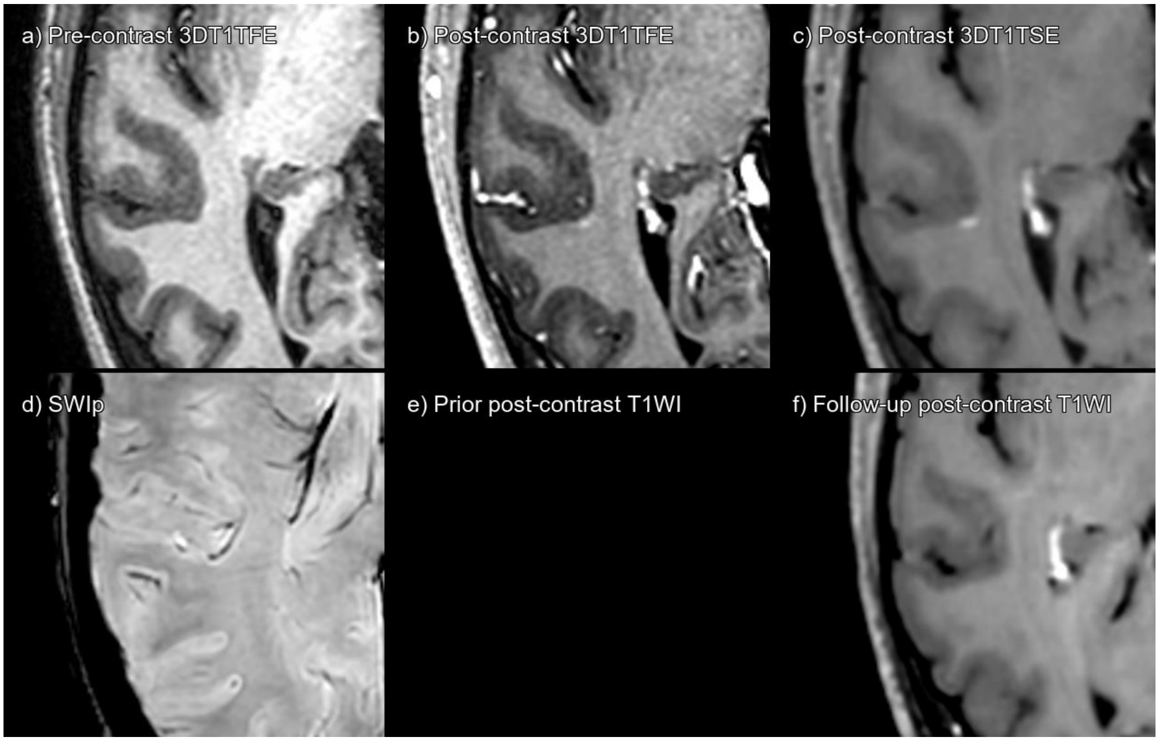


Supplementary figure 24. Lesion 146_04 a) 3DT1TFE without contrast, b) 3DT1TFE post-contrast, c) 3DT1TSE post-contrast, d) SWIp, e) Prior post-contrast T1-weighted imaging not available, f) Follow-up post-contrast T1-weighted imaging 6 months post-baseline.

True enhancing juxtacortical lesion, that has disappeared on follow-up MRI.

| **CONSENSUS** | **True Enhancing MS Lesion** | | | |
|---|---|---|---|---|
| | **TFE sequence** | | **TSE sequence** | |
| Visible in sequence | Yes | | Yes | |
| | **Reader 1** | **Reader 2** | **Reader 1** | **Reader 2** |
| **Detected** | Yes | No | Yes | Yes |

Abbreviations: Multiple sclerosis (MS), Turbo Field Echo (TFE), Turbo Spin Echo (TSE), Susceptibility Weighted Imaging with phase enhancement (SWIp).

#### LESION 146_05

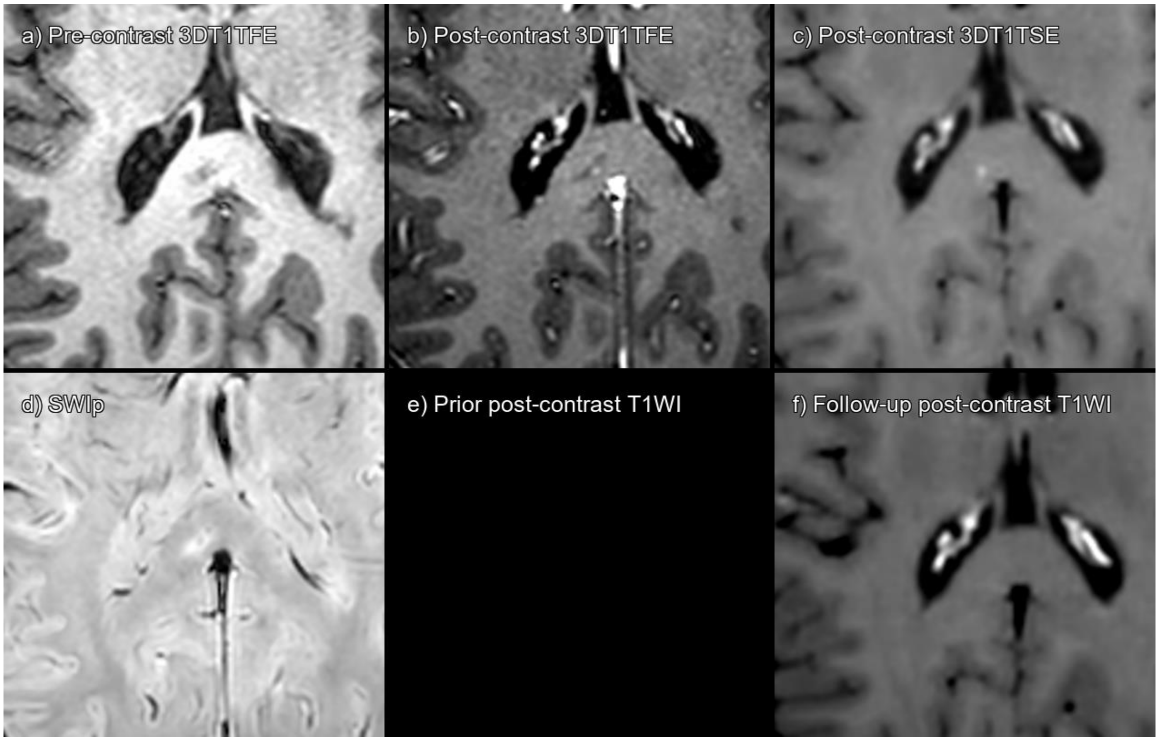


Supplementary figure 25. Lesion 146_05 a) 3DT1TFE without contrast, b) 3DT1TFE post-contrast, c) 3DT1TSE post-contrast, d) SWIp, e) Prior post-contrast T1-weighted imaging not available, f) Follow-up post-contrast T1-weighted imaging 2 years post-baseline.

True enhancing corpus callosum lesion, that has disappeared on follow-up MRI.

| **CONSENSUS** | **True Enhancing MS Lesion** | | | |
|---|---|---|---|---|
| | **TFE sequence** | | **TSE sequence** | |
| Visible in sequence | Yes | | Yes | |
| | **Reader 1** | **Reader 2** | **Reader 1** | **Reader 2** |
| **Detected** | Yes | No | Yes | No |

Abbreviations: Multiple sclerosis (MS), Turbo Field Echo (TFE), Turbo Spin Echo (TSE), Susceptibility Weighted Imaging with phase enhancement (SWIp).

#### LESION 146_06

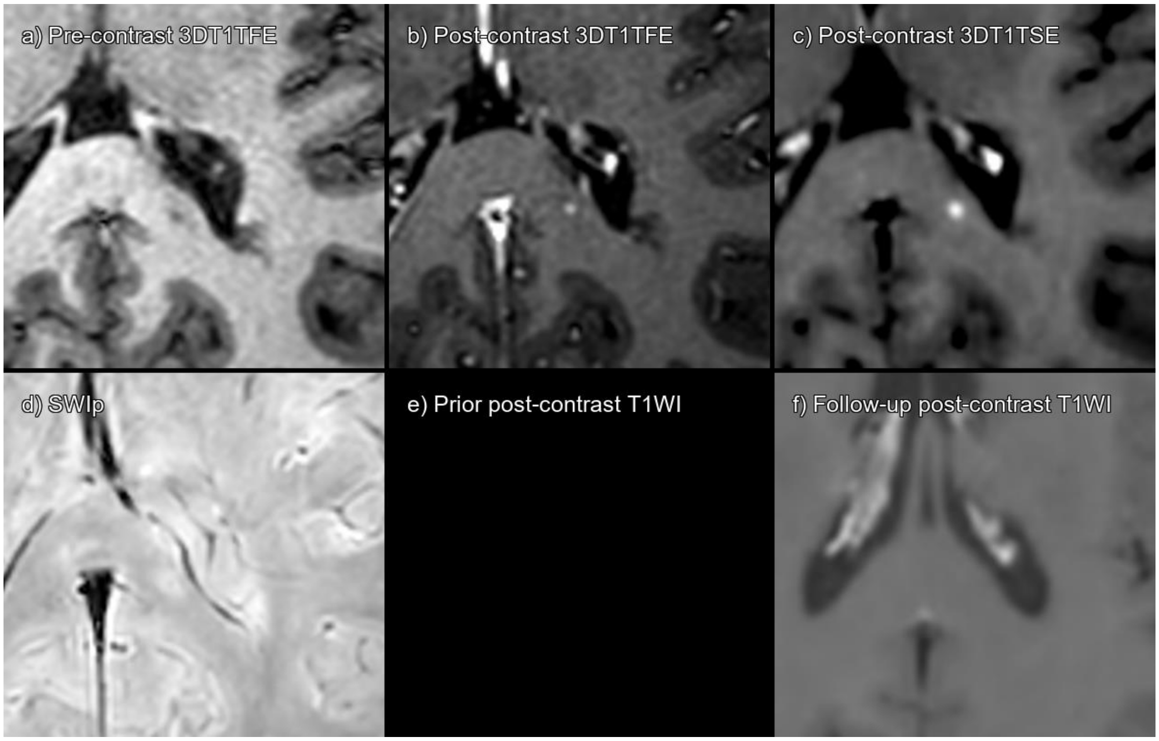


Supplementary figure 26. Lesion 146_06 a) 3DT1TFE without contrast, b) 3DT1TFE post-contrast, c) 3DT1TSE post-contrast, d) SWIp, e) Prior post-contrast T1-weighted imaging not available, f) Follow-up post-contrast T1-weighted imaging 2 years post-baseline.

True enhancing corpus callosum lesion, that has disappeared on follow-up MRI.

| **CONSENSUS** | **True Enhancing MS Lesion** | | | |
|---|---|---|---|---|
| | **TFE sequence** | | **TSE sequence** | |
| Visible in sequence | Yes | | Yes | |
| | **Reader 1** | **Reader 2** | **Reader 1** | **Reader 2** |
| **Detected** | No | No | Yes | No |

Abbreviations: Multiple sclerosis (MS), Turbo Field Echo (TFE), Turbo Spin Echo (TSE), Susceptibility Weighted Imaging with phase enhancement (SWIp).

#### LESION 160_01

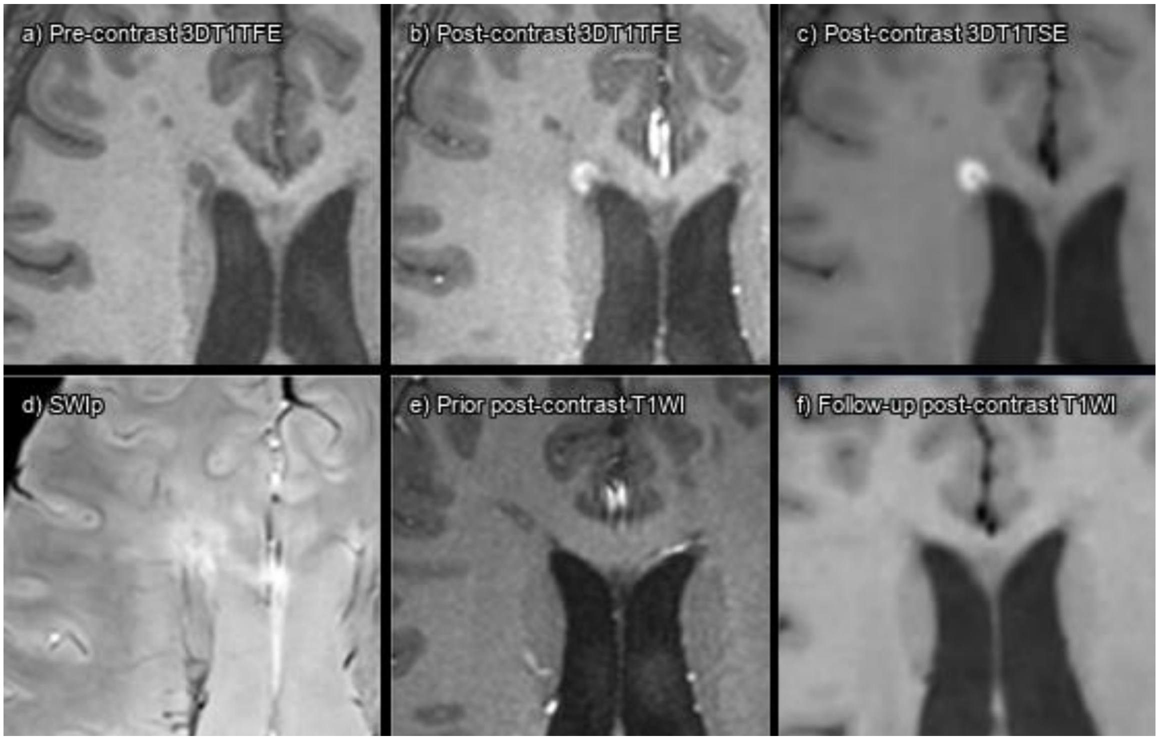


Supplementary figure 27. Lesion 160_01 a) 3DT1TFE without contrast, b) 3DT1TFE post-contrast, c) 3DT1TSE post-contrast, d) SWIp, e) Prior post-contrast T1-weighted imaging 17 months pre-baseline, f) Follow-up post-contrast T1-weighted imaging 12 months post-baseline.

True enhancing periventricular lesion, that has disappeared on follow-up MRI.

| **CONSENSUS** | **True Enhancing MS Lesion** | | | |
|---|---|---|---|---|
| | **TFE sequence** | | **TSE sequence** | |
| Visible in sequence | Yes | | Yes | |
| | **Reader 1** | **Reader 2** | **Reader 1** | **Reader 2** |
| **Detected** | Yes | Yes | Yes | Yes |

Abbreviations: Multiple sclerosis (MS), Turbo Field Echo (TFE), Turbo Spin Echo (TSE), Susceptibility Weighted Imaging with phase enhancement (SWIp).

#### LESION 162_01

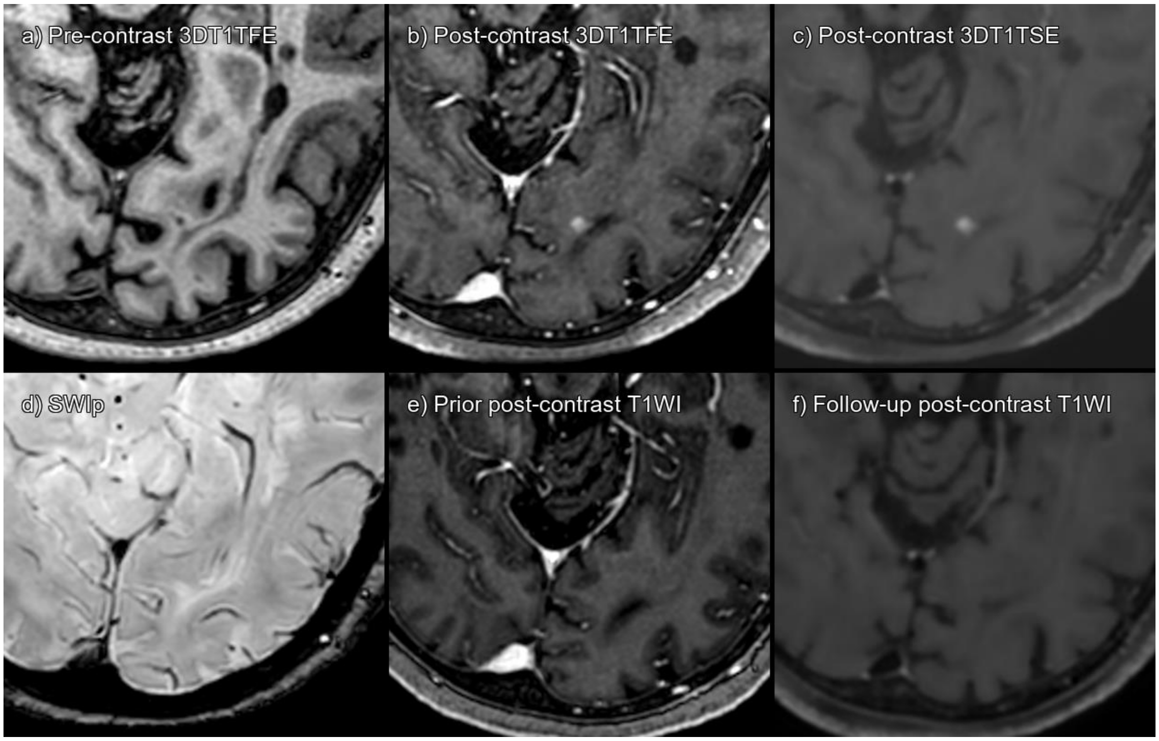


Supplementary figure 28. Lesion 162_01 a) 3DT1TFE without contrast, b) 3DT1TFE post-contrast, c) 3DT1TSE post-contrast, d) SWIp, e) Prior post-contrast T1-weighted imaging 6 years pre-baseline, f) Follow-up post-contrast T1-weighted imaging 8 months post-baseline.

True enhancing juxtacortical lesion, that has disappeared on follow-up MRI.

| **CONSENSUS** | **True Enhancing MS Lesion** | | | |
|---|---|---|---|---|
| | **TFE sequence** | | **TSE sequence** | |
| Visible in sequence | Yes | | Yes | |
| | **Reader 1** | **Reader 2** | **Reader 1** | **Reader 2** |
| **Detected** | No | Yes | Yes | Yes |

Abbreviations: Multiple sclerosis (MS), Turbo Field Echo (TFE), Turbo Spin Echo (TSE), Susceptibility Weighted Imaging with phase enhancement (SWIp).

#### LESION 162_02

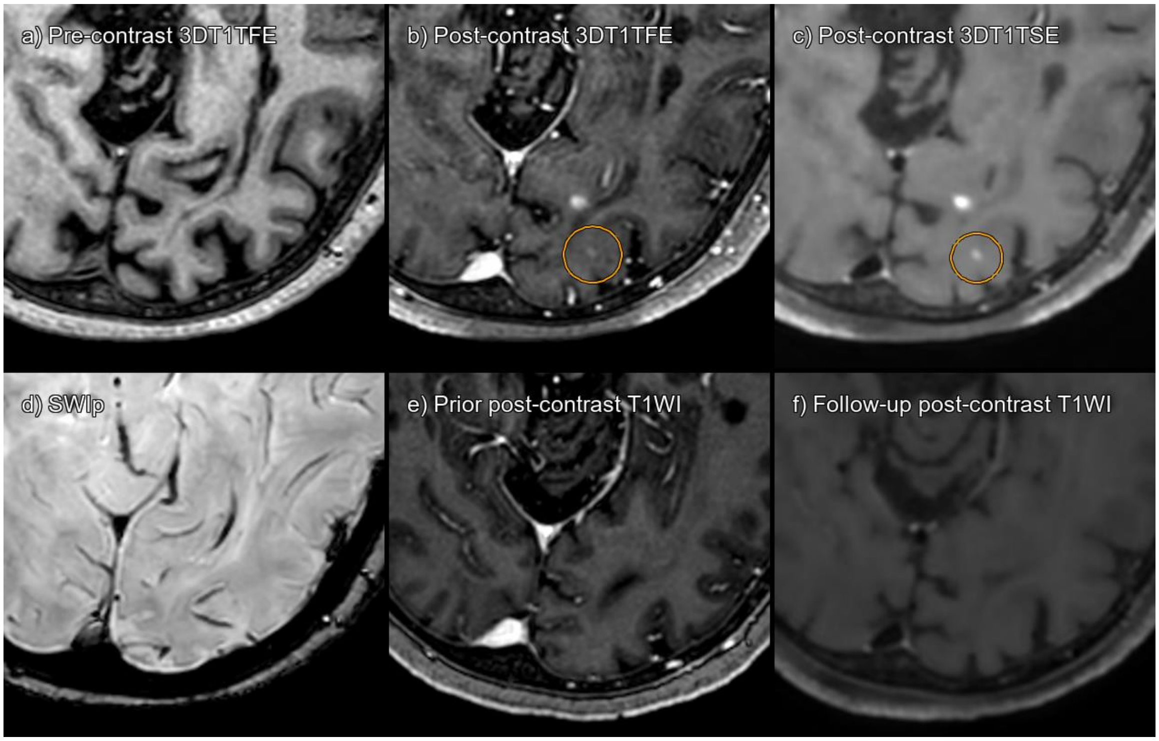


Supplementary figure 29. Lesion 162_02 a) 3DT1TFE without contrast, b) 3DT1TFE post-contrast, c) 3DT1TSE post-contrast, d) SWIp, e) Prior post-contrast T1-weighted imaging 6 years pre-baseline, f) Follow-up post-contrast T1-weighted imaging 8 months post-baseline.

True enhancing juxtacortical lesion, that has disappeared on follow-up MRI.

| **CONSENSUS** | **True Enhancing MS Lesion** | | | |
|---|---|---|---|---|
| | **TFE sequence** | | **TSE sequence** | |
| Visible in sequence | Yes | | Yes | |
| | **Reader 1** | **Reader 2** | **Reader 1** | **Reader 2** |
| **Detected** | No | No | Yes | Yes |

Abbreviations: Multiple sclerosis (MS), Turbo Field Echo (TFE), Turbo Spin Echo (TSE), Susceptibility Weighted Imaging with phase enhancement (SWIp).

#### LESION 162_03

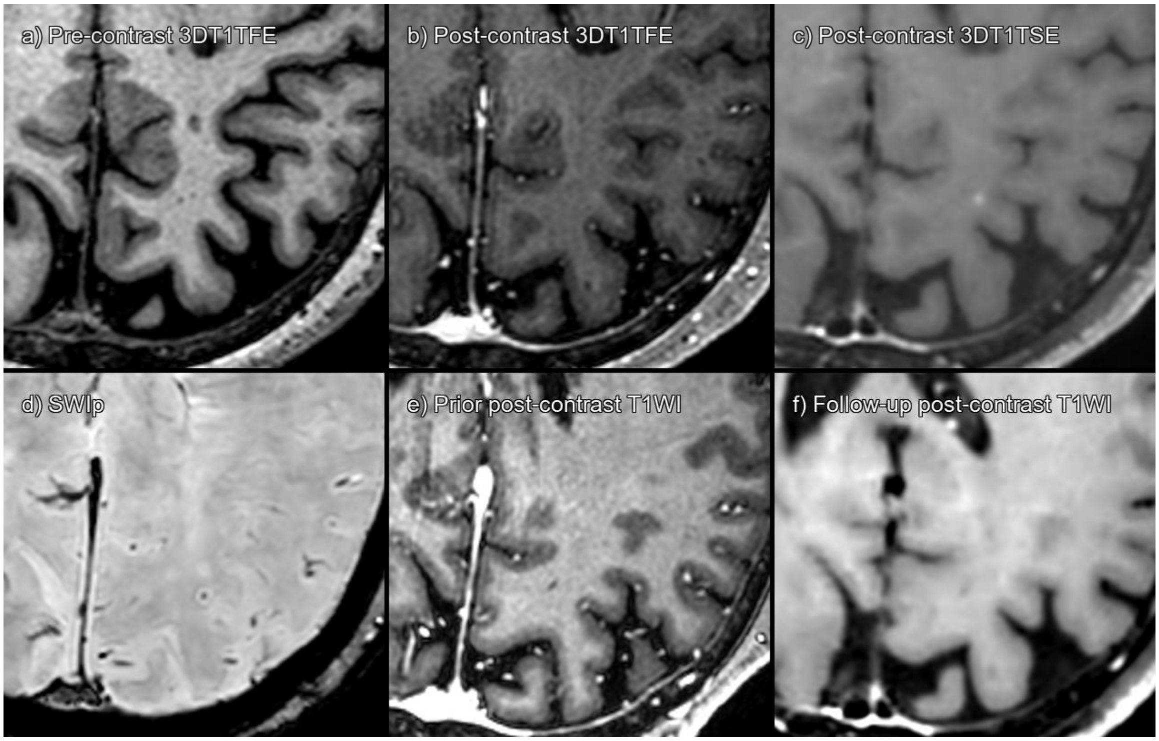


Supplementary figure 30. Lesion 162_03 a) 3DT1TFE without contrast, b) 3DT1TFE post-contrast, c) 3DT1TSE post-contrast, d) SWIp, e) Prior post-contrast T1-weighted imaging 6 years pre-baseline, f) Follow-up post-contrast T1-weighted imaging 8 months post-baseline.

True enhancing juxtacortical lesion, not visible on 3DT1TFE, but visible on 3DT1TSE, that has disappeared on follow-up MRI.

| **CONSENSUS** | **True Enhancing MS Lesion** | | | |
|---|---|---|---|---|
| | **TFE sequence** | | **TSE sequence** | |
| Visible in sequence | No | | Yes | |
| | **Reader 1** | **Reader 2** | **Reader 1** | **Reader 2** |
| **Detected** | No | No | Yes | Yes |

Abbreviations: Multiple sclerosis (MS), Turbo Field Echo (TFE), Turbo Spin Echo (TSE), Susceptibility Weighted Imaging with phase enhancement (SWIp).

#### LESION 162_04

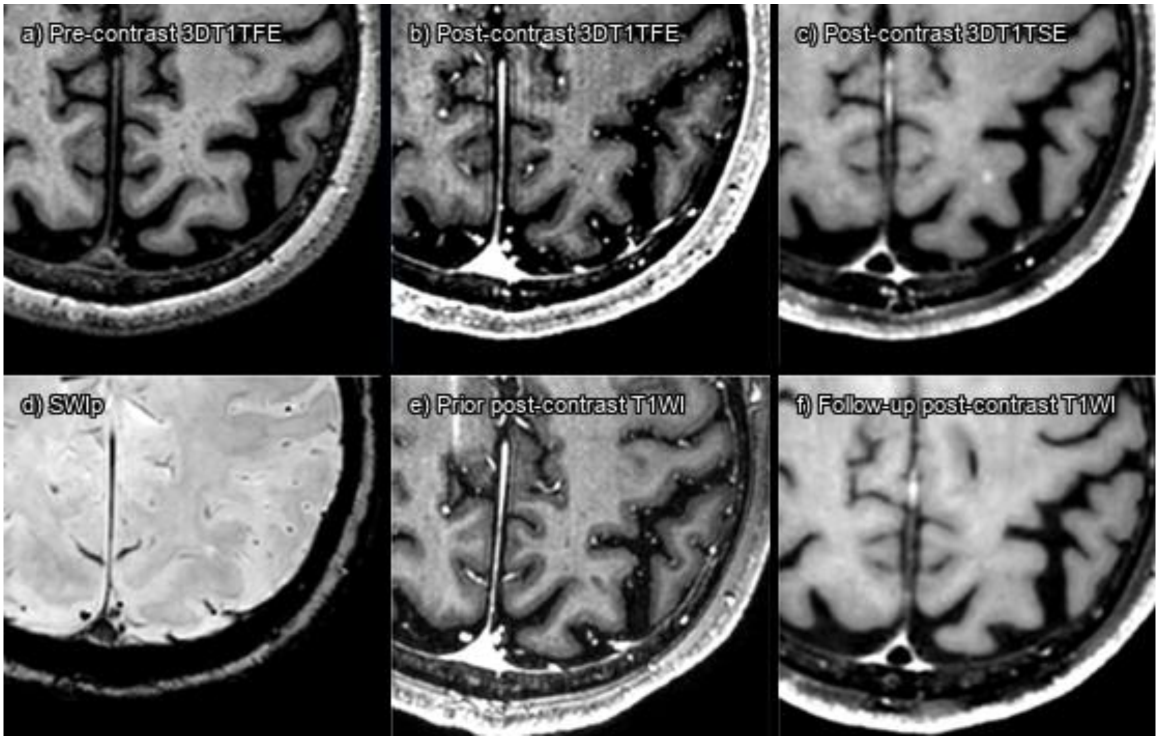


Supplementary figure 31. Lesion 162_04 a) 3DT1TFE without contrast, b) 3DT1TFE post-contrast, c) 3DT1TSE post-contrast, d) SWIp, e) Prior post-contrast T1-weighted imaging 6 months pre-baseline, f) Follow-up post-contrast T1-weighted imaging 8 months post-baseline.

True enhancing juxtacortical lesion, that has disappeared on follow-up MRI.

| **CONSENSUS** | **True Enhancing MS Lesion** | | | |
|---|---|---|---|---|
| | **TFE sequence** | | **TSE sequence** | |
| Visible in sequence | Yes | | Yes | |
| | **Reader 1** | **Reader 2** | **Reader 1** | **Reader 2** |
| **Detected** | No | No | Yes | No |

Abbreviations: Multiple sclerosis (MS), Turbo Field Echo (TFE), Turbo Spin Echo (TSE), Susceptibility Weighted Imaging with phase enhancement (SWIp).

#### LESION 165_01

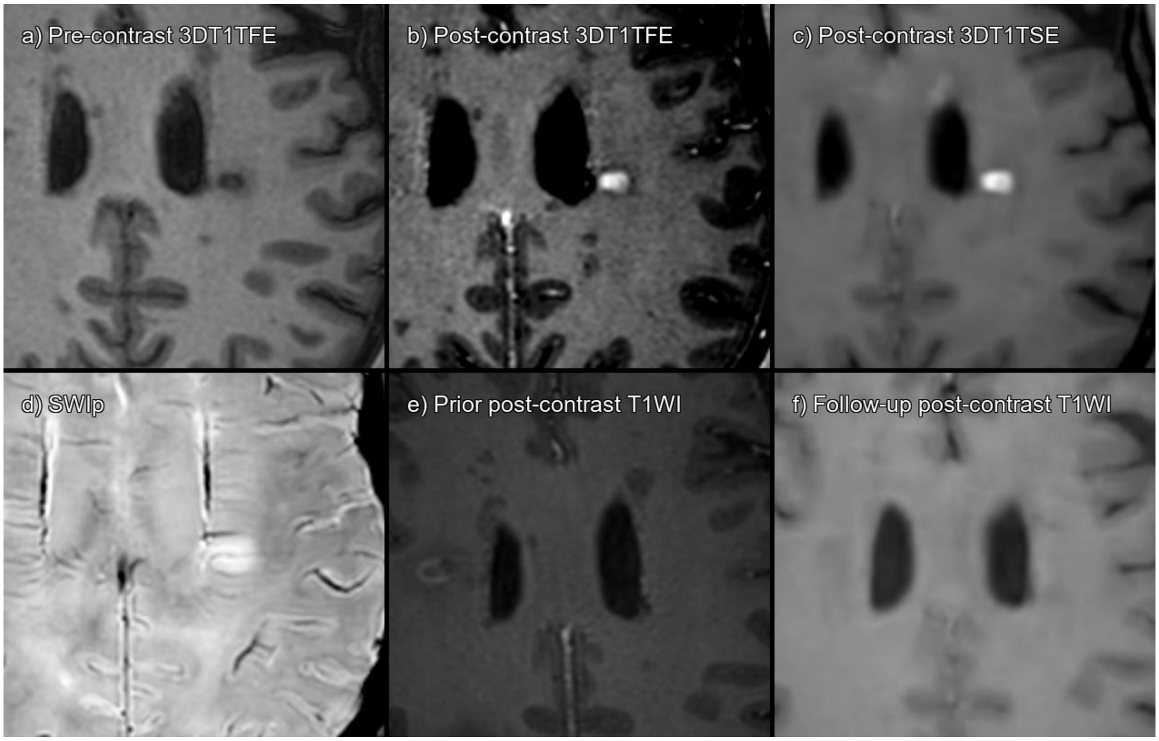


Supplementary figure 32. Lesion 165_01 a) 3DT1TFE without contrast, b) 3DT1TFE post-contrast, c) 3DT1TSE post-contrast, d) SWIp, e) Prior post-contrast T1-weighted imaging 6 months pre-baseline, f) Follow-up post-contrast T1-weighted imaging 8 months post-baseline.

True enhancing lesion, that has disappeared on follow-up MRI.

| **CONSENSUS** | **True Enhancing MS Lesion** | | | |
|---|---|---|---|---|
| | **TFE sequence** | | **TSE sequence** | |
| Visible in sequence | Yes | | Yes | |
| | **Reader 1** | **Reader 2** | **Reader 1** | **Reader 2** |
| **Detected** | Yes | Yes | Yes | Yes |

Abbreviations: Multiple sclerosis (MS), Turbo Field Echo (TFE), Turbo Spin Echo (TSE), Susceptibility Weighted Imaging with phase enhancement (SWIp).

#### LESION 165_02

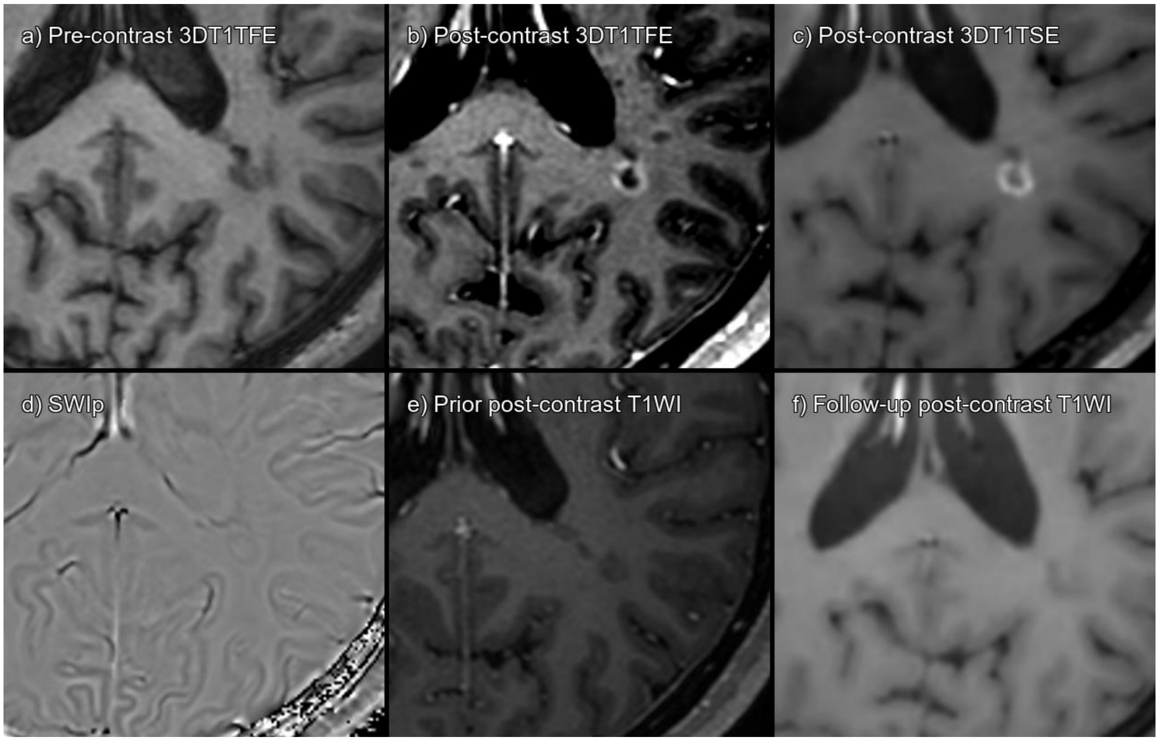


Supplementary figure 33. Lesion 165_02 a) 3DT1TFE without contrast, b) 3DT1TFE post-contrast, c) 3DT1TSE post-contrast, d) SWIp, e) Prior post-contrast T1-weighted imaging 6 months pre-baseline, f) Follow-up post-contrast T1-weighted imaging 8 months post-baseline.

True enhancing lesion, that has disappeared on follow-up MRI.

| **CONSENSUS** | **True Enhancing MS Lesion** | | | |
|---|---|---|---|---|
| | **TFE sequence** | | **TSE sequence** | |
| Visible in sequence | Yes | | Yes | |
| | **Reader 1** | **Reader 2** | **Reader 1** | **Reader 2** |
| **Detected** | Yes | Yes | Yes | Yes |

Abbreviations: Multiple sclerosis (MS), Turbo Field Echo (TFE), Turbo Spin Echo (TSE), Susceptibility Weighted Imaging with phase enhancement (SWIp).

#### LESION 165_03

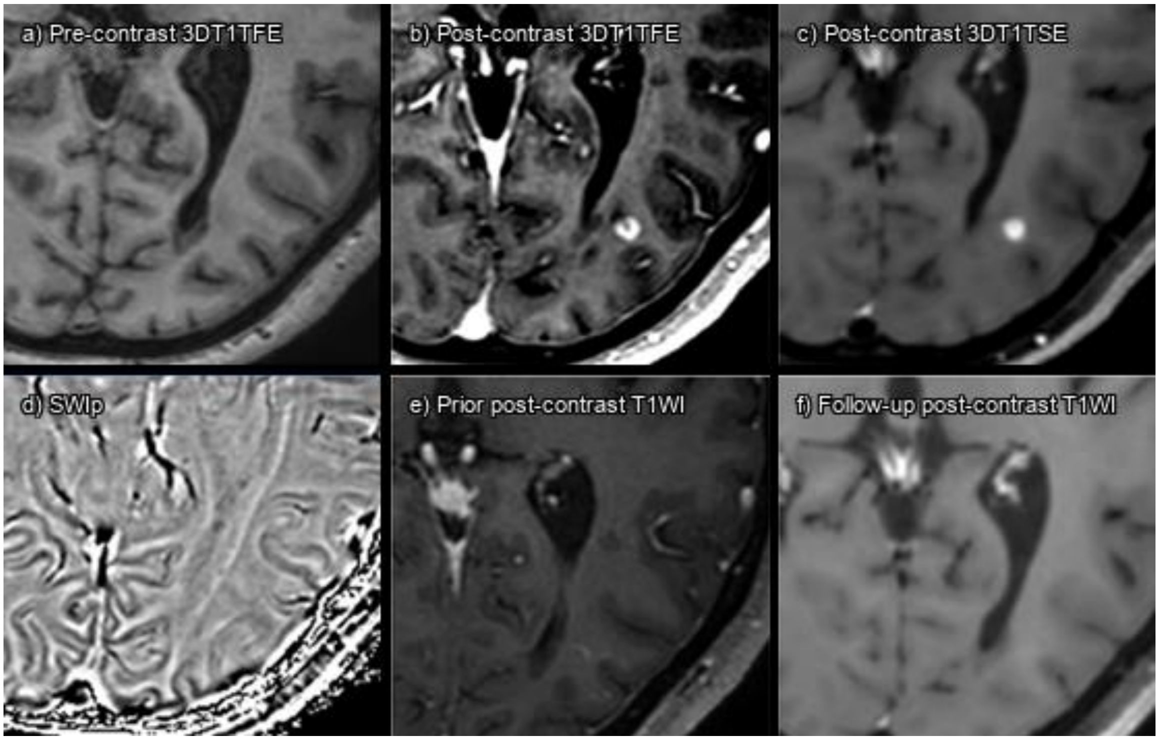


Supplementary figure 34. Lesion 165_03 a) 3DT1TFE without contrast, b) 3DT1TFE post-contrast, c) 3DT1TSE post-contrast, d) SWIp phase, e) Prior post-contrast T1-weighted imaging 6 months pre-baseline, f) Follow-up post-contrast T1-weighted imaging 8 months post-baseline.

True enhancing lesion, that has disappeared on follow-up MRI.

| **CONSENSUS** | **True Enhancing MS Lesion** | | | |
|---|---|---|---|---|
| | **TFE sequence** | | **TSE sequence** | |
| Visible in sequence | Yes | | Yes | |
| | **Reader 1** | **Reader 2** | **Reader 1** | **Reader 2** |
| **Detected** | Yes | Yes | Yes | Yes |

Abbreviations: Multiple sclerosis (MS), Turbo Field Echo (TFE), Turbo Spin Echo (TSE), Susceptibility Weighted Imaging with phase enhancement (SWIp).

### LESION 172_01

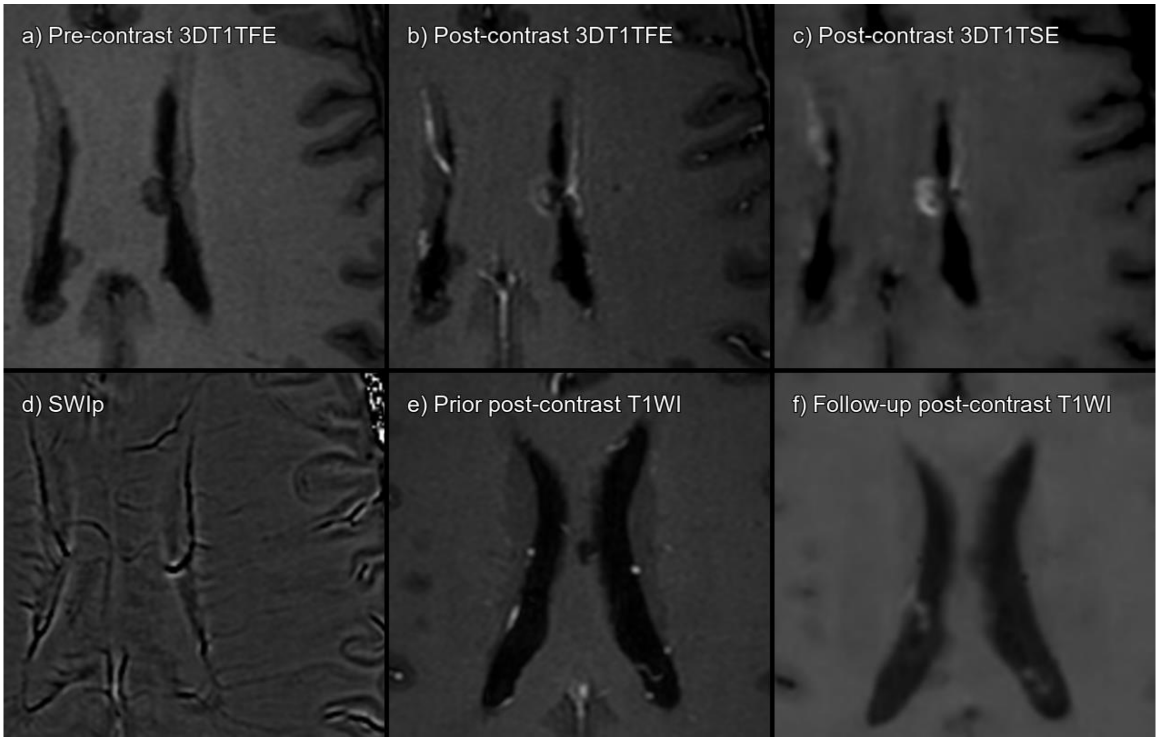


Supplementary figure 35. Lesion 172_01 a) 3DT1TFE without contrast, b) 3DT1TFE post-contrast, c) 3DT1TSE post-contrast, d) SWIp phase, e) Prior post-contrast T1-weighted imaging 6 months pre-baseline, f) Follow-up post-contrast T1-weighted imaging 6 months post-baseline.

True enhancing lesion, that has disappeared on follow-up MRI.

| **CONSENSUS** | **True Enhancing MS Lesion** | | | |
|---|---|---|---|---|
| | **TFE sequence** | | **TSE sequence** | |
| Visible in sequence | Yes | | Yes | |
| | **Reader 1** | **Reader 2** | **Reader 1** | **Reader 2** |
| **Detected** | Yes | No | Yes | Yes |

Abbreviations: Multiple sclerosis (MS), Turbo Field Echo (TFE), Turbo Spin Echo (TSE), Susceptibility Weighted Imaging with phase enhancement (SWIp).

#### LESION 173_01

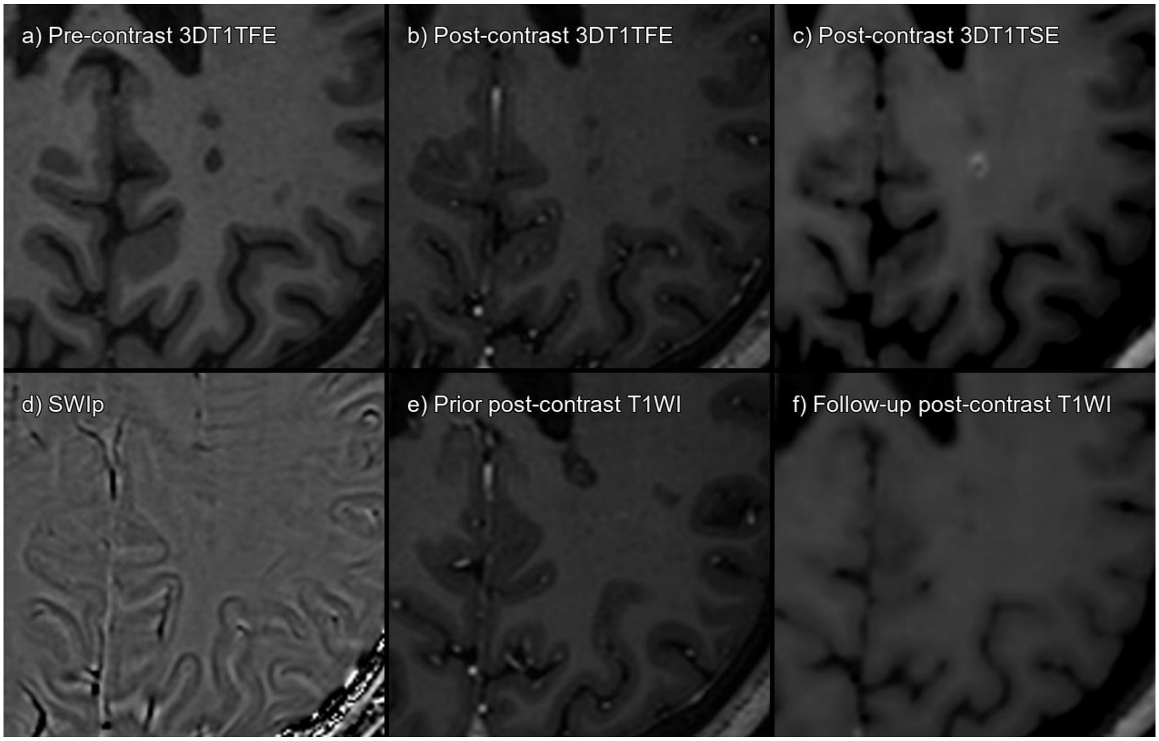


Supplementary figure 36. Lesion 173_01 a) 3DT1TFE without contrast, b) 3DT1TFE post-contrast, c) 3DT1TSE post-contrast, d) SWIp phase, e) Prior post-contrast T1-weighted imaging 6 months pre-baseline, f) Follow-up post-contrast T1-weighted imaging 6 months post-baseline.

True enhancing lesion, visible on 3DT1TSE but not on 3DT1TFE, that has disappeared on follow-up MRI.

| **CONSENSUS** | **True Enhancing MS Lesion** | | | |
|---|---|---|---|---|
| | **TFE sequence** | | **TSE sequence** | |
| Visible in sequence | No | | Yes | |
| | **Reader 1** | **Reader 2** | **Reader 1** | **Reader 2** |
| **Detected** | No | No | Yes | Yes |

Abbreviations: Multiple sclerosis (MS), Turbo Field Echo (TFE), Turbo Spin Echo (TSE), Susceptibility Weighted Imaging with phase enhancement (SWIp).

#### LESION 176_01

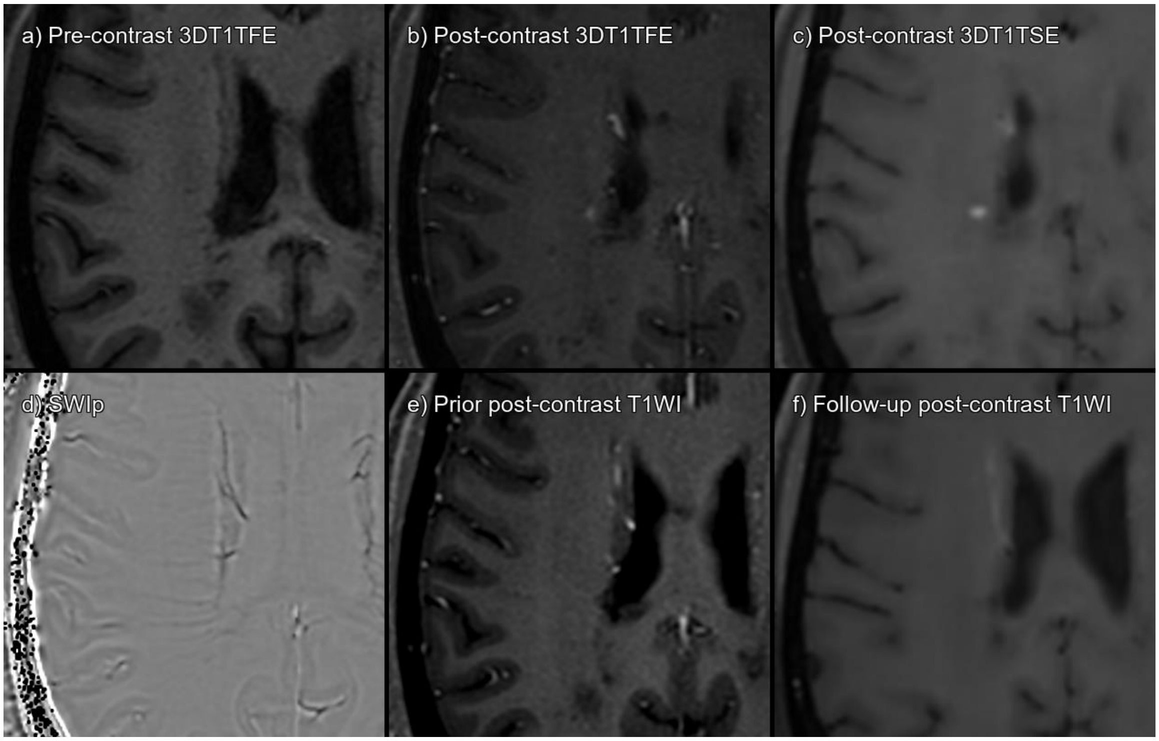


Supplementary figure 37. Lesion 176_01 a) 3DT1TFE without contrast, b) 3DT1TFE post-contrast, c) 3DT1TSE post-contrast, d) SWIp phase, e) Prior post-contrast T1-weighted imaging 2 years pre-baseline, f) Follow-up post-contrast T1-weighted imaging 1 year post-baseline.

True enhancing lesion that has disappeared on follow-up MRI.

| **CONSENSUS** | **True Enhancing MS Lesion** | | | |
|---|---|---|---|---|
| | **TFE sequence** | | **TSE sequence** | |
| Visible in sequence | Yes | | Yes | |
| | **Reader 1** | **Reader 2** | **Reader 1** | **Reader 2** |
| **Detected** | Yes | Yes | Yes | Yes |

Abbreviations: Multiple sclerosis (MS), Turbo Field Echo (TFE), Turbo Spin Echo (TSE), Susceptibility Weighted Imaging with phase enhancement (SWIp).

#### LESION 176_02

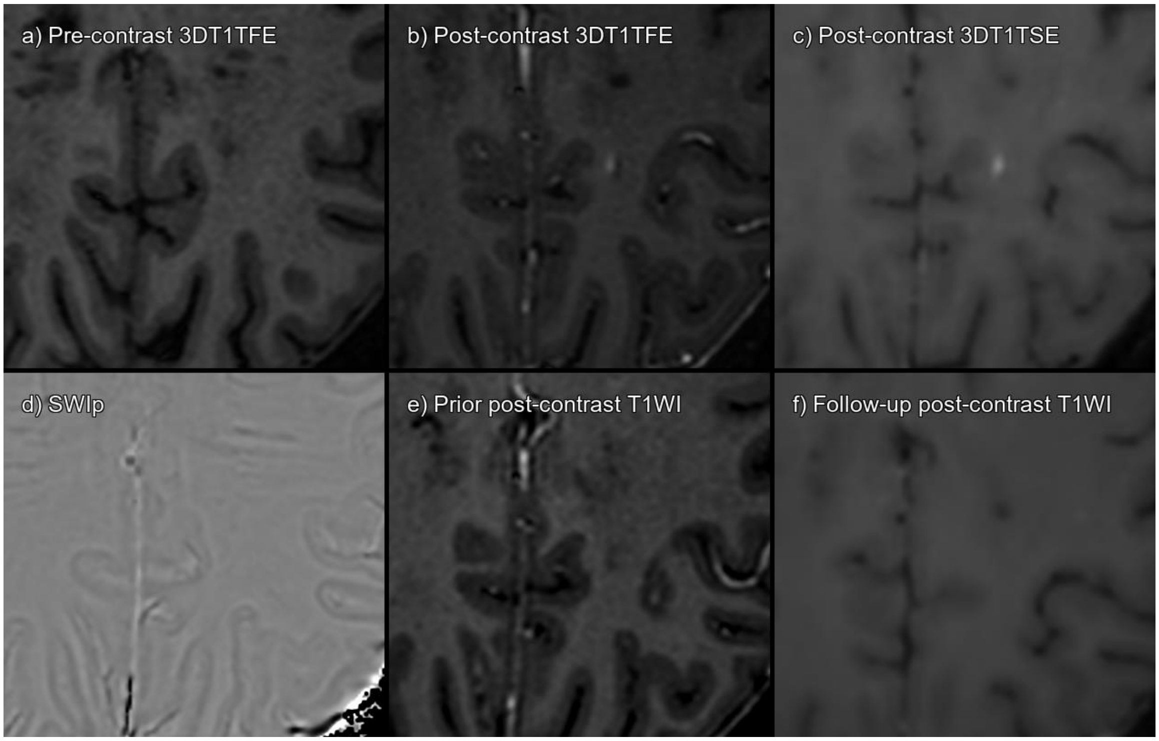


Supplementary figure 38. Lesion 176_02 a) 3DT1TFE without contrast, b) 3DT1TFE post-contrast, c) 3DT1TSE post-contrast, d) SWIp phase, e) Prior post-contrast T1-weighted imaging 2 years pre-baseline, f) Follow-up post-contrast T1-weighted imaging 1 year post-baseline.

True enhancing lesion that has disappeared on follow-up MRI.

| **CONSENSUS** | **True Enhancing MS Lesion** | | | |
|---|---|---|---|---|
| | **TFE sequence** | | **TSE sequence** | |
| Visible in sequence | Yes | | Yes | |
| | **Reader 1** | **Reader 2** | **Reader 1** | **Reader 2** |
| **Detected** | No | Yes | Yes | Yes |

Abbreviations: Multiple sclerosis (MS), Turbo Field Echo (TFE), Turbo Spin Echo (TSE), Susceptibility Weighted Imaging with phase enhancement (SWIp).

#### LESION 178_01

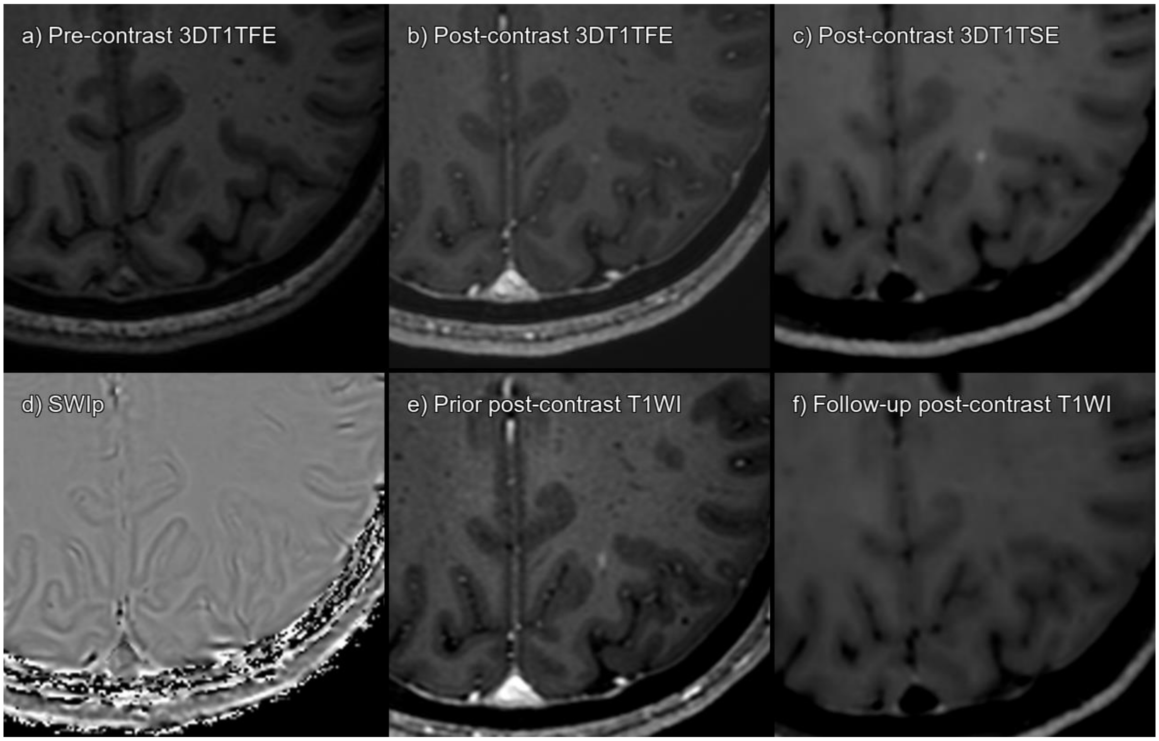


Supplementary figure 39. Lesion 178_01 a) 3DT1TFE without contrast, b) 3DT1TFE post-contrast, c) 3DT1TSE post-contrast, d) SWIp phase, e) Prior post-contrast T1-weighted imaging 1 year pre-baseline, f) Follow-up post-contrast T1-weighted imaging 1 year post-baseline.

Enhancing subcortical lesion visible on 3DT1TFE and on 3DT1TSE post-contrast. Was already present in prior scan. Note that on SWIp it seems to correspond to a small vessel. Consensus established false positive enhancement, probably corresponding to a small intraparenchymal blood vessel. On follow-up imaging it is not visible.

| **CONSENSUS** | **Not acute MS lesion enhancement** | | | |
|---|---|---|---|---|
| | **TFE sequence** | | **TSE sequence** | |
| Visible in sequence | Yes | | Yes | |
| | **Reader 1** | **Reader 2** | **Reader 1** | **Reader 2** |
| **Detected** | No | No | No | Yes |

Abbreviations: Multiple sclerosis (MS), Turbo Field Echo (TFE), Turbo Spin Echo (TSE), Susceptibility Weighted Imaging with phase enhancement (SWIp).

#### LESION 181_01

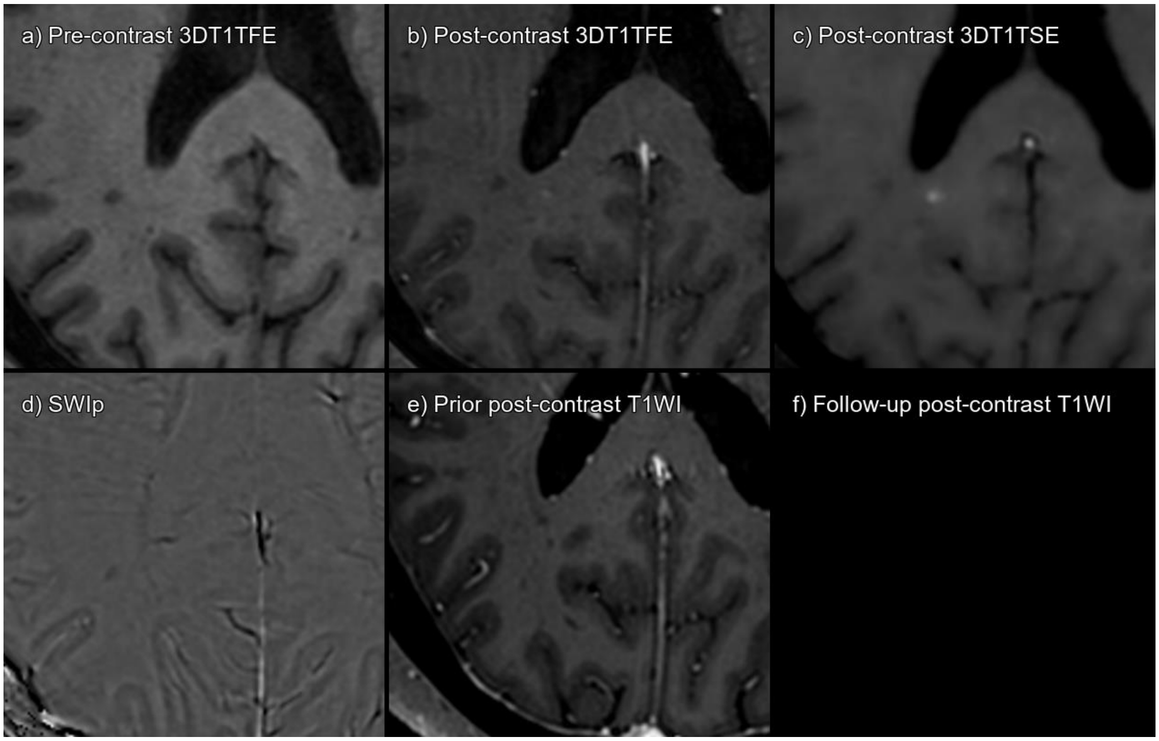


Supplementary figure 40. Lesion 181_01 a) 3DT1TFE without contrast, b) 3DT1TFE post-contrast, c) 3DT1TSE post-contrast, d) SWIp phase, e) Prior post-contrast T1-weighted imaging 3 years pre-baseline, f) Follow-up post-contrast T1-weighted imaging not available.

True enhancing lesion that has disappeared on follow-up MRI.

| **CONSENSUS** | **True Enhancing MS Lesion** | | | |
|---|---|---|---|---|
| | **TFE sequence** | | **TSE sequence** | |
| Visible in sequence | Yes | | Yes | |
| | **Reader 1** | **Reader 2** | **Reader 1** | **Reader 2** |
| **Detected** | No | No | Yes | Yes |

Abbreviations: Multiple sclerosis (MS), Turbo Field Echo (TFE), Turbo Spin Echo (TSE), Susceptibility Weighted Imaging with phase enhancement (SWIp).

#### LESION 193_01

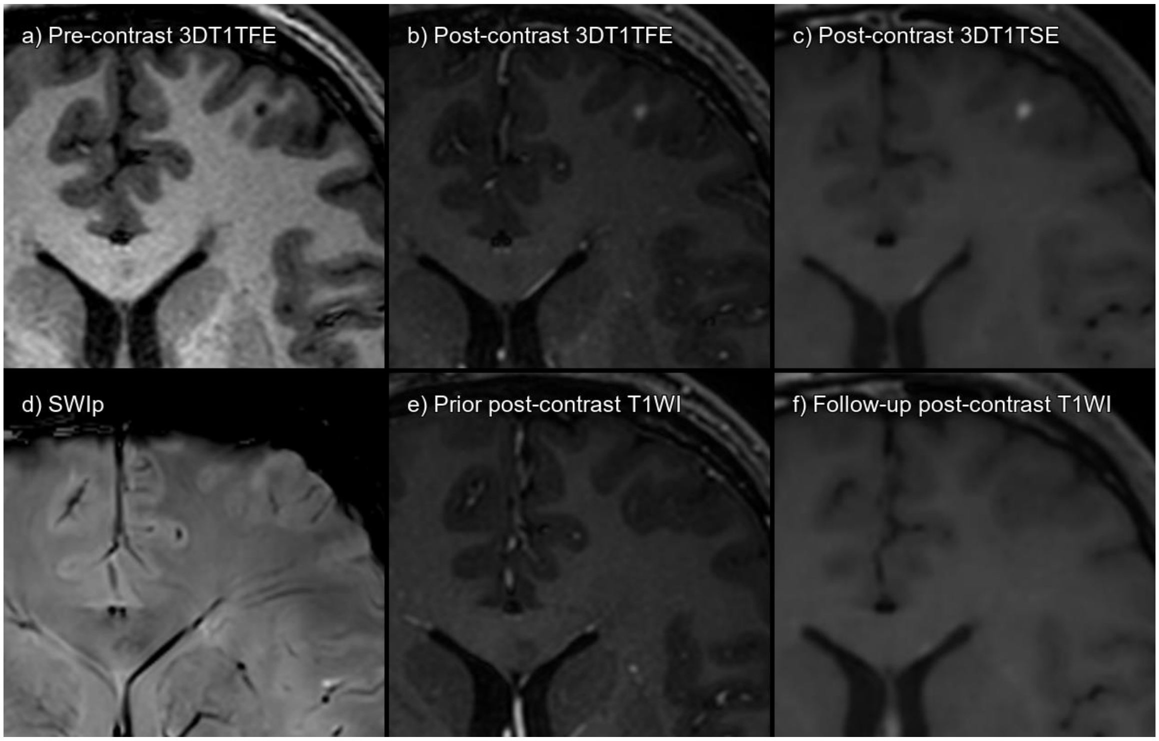


Supplementary figure 41. Lesion 193_01 a) 3DT1TFE without contrast, b) 3DT1TFE post-contrast, c) 3DT1TSE post-contrast, d) SWIp, e) Prior post-contrast T1-weighted imaging 9 months pre-baseline, f) Follow-up post-contrast T1-weighted imaging 1 year post-baseline.

True enhancing lesion that has disappeared on follow-up MRI.

| **CONSENSUS** | **True Enhancing MS Lesion** | | | |
|---|---|---|---|---|
| | **TFE sequence** | | **TSE sequence** | |
| Visible in sequence | Yes | | Yes | |
| | **Reader 1** | **Reader 2** | **Reader 1** | **Reader 2** |
| **Detected** | No | No | Yes | Yes |

Abbreviations: Multiple sclerosis (MS), Turbo Field Echo (TFE), Turbo Spin Echo (TSE), Susceptibility Weighted Imaging with phase enhancement (SWIp).

#### LESION 195_01

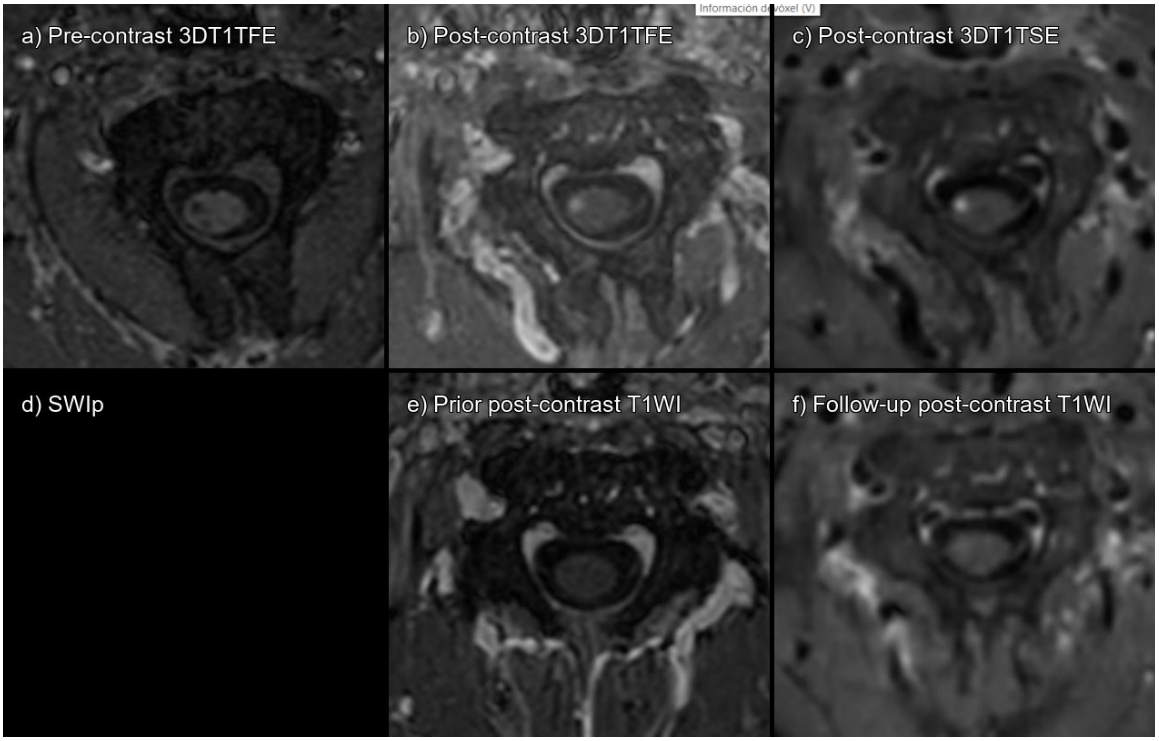


Supplementary figure 42. Lesion 195_01 a) 3DT1TFE without contrast, b) 3DT1TFE post-contrast, c) 3DT1TSE post-contrast, d) SWIp acquisition did not include this cervical level, e) Prior post-contrast T1-weighted imaging 1 year pre-baseline, f) Follow-up post-contrast T1-weighted imaging 9 months post-baseline.

True enhancing lesion that has disappeared on follow-up MRI.

| **CONSENSUS** | **True Enhancing MS Lesion** | | | |
|---|---|---|---|---|
| | **TFE sequence** | | **TSE sequence** | |
| Visible in sequence | Yes | | Yes | |
| | **Reader 1** | **Reader 2** | **Reader 1** | **Reader 2** |
| **Detected** | Yes | Yes | Yes | Yes |

Abbreviations: Multiple sclerosis (MS), Turbo Field Echo (TFE), Turbo Spin Echo (TSE), Susceptibility Weighted Imaging with phase enhancement (SWIp).

#### LESION 195_02

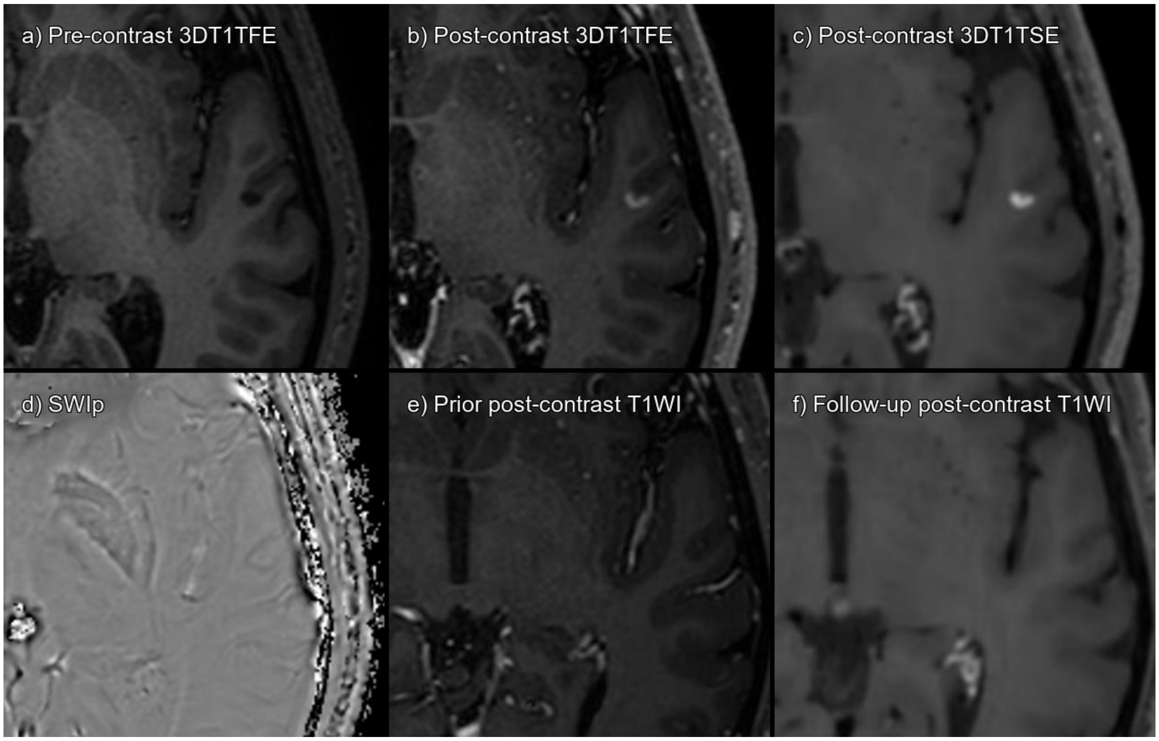


Supplementary figure 43. Lesion 195_02 a) 3DT1TFE without contrast, b) 3DT1TFE post-contrast, c) 3DT1TSE post-contrast, d) SWIp phase, e) Prior post-contrast T1-weighted imaging 1 year pre-baseline, f) Follow-up post-contrast T1-weighted imaging 9 months post-baseline.

True enhancing lesion that has disappeared on follow-up MRI.

| **CONSENSUS** | **True Enhancing MS Lesion** | | | |
|---|---|---|---|---|
| | **TFE sequence** | | **TSE sequence** | |
| Visible in sequence | Yes | | Yes | |
| | **Reader 1** | **Reader 2** | **Reader 1** | **Reader 2** |
| **Detected** | No | No | Yes | Yes |

Abbreviations: Multiple sclerosis (MS), Turbo Field Echo (TFE), Turbo Spin Echo (TSE), Susceptibility Weighted Imaging with phase enhancement (SWIp).

#### LESION 195_03

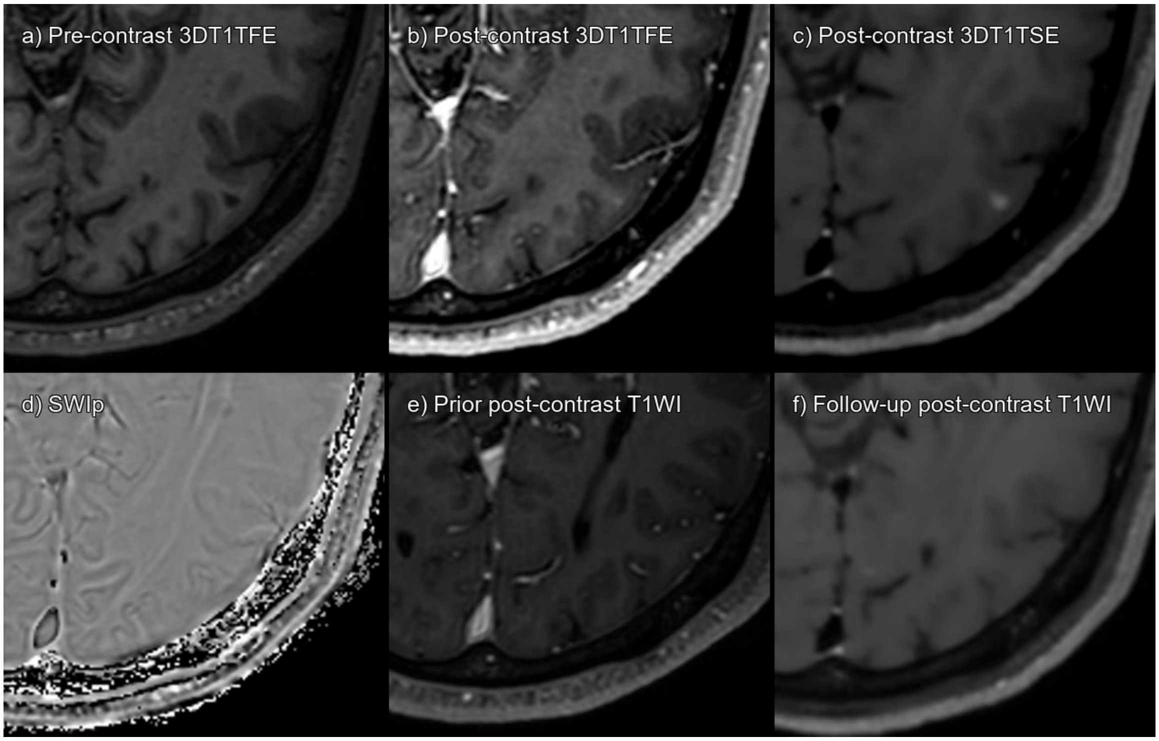


Supplementary figure 44. Lesion 195_03 a) 3DT1TFE without contrast, b) 3DT1TFE post-contrast, c) 3DT1TSE post-contrast, d) SWIp phase, e) Prior post-contrast T1-weighted imaging 1 year pre-baseline, f) Follow-up post-contrast T1-weighted imaging 9 months post-baseline.

True enhancing lesion that has disappeared on follow-up MRI.

| **CONSENSUS** | **True Enhancing MS Lesion** | | | |
|---|---|---|---|---|
| | **TFE sequence** | | **TSE sequence** | |
| Visible in sequence | Yes | | Yes | |
| | **Reader 1** | **Reader 2** | **Reader 1** | **Reader 2** |
| **Detected** | No | No | Yes | Yes |

Abbreviations: Multiple sclerosis (MS), Turbo Field Echo (TFE), Turbo Spin Echo (TSE), Susceptibility Weighted Imaging with phase enhancement (SWIp).

#### LESION 195_04

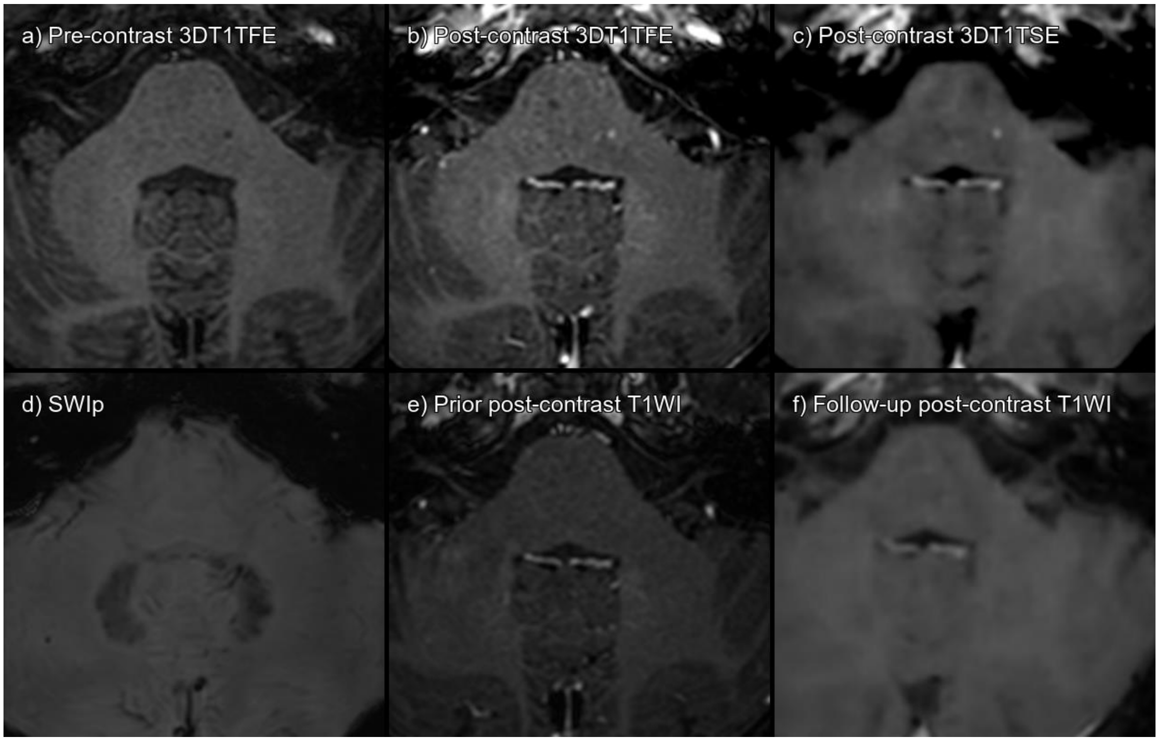


Supplementary figure 45. Lesion 195_04 a) 3DT1TFE without contrast, b) 3DT1TFE post-contrast, c) 3DT1TSE post-contrast, d) SWIp phase, e) Prior post-contrast T1-weighted imaging 1 year pre-baseline, f) Follow-up post-contrast T1-weighted imaging 9 months post-baseline.

True enhancing lesion that has disappeared on follow-up MRI.

| **CONSENSUS** | **True Enhancing MS Lesion** | | | |
|---|---|---|---|---|
| | **TFE sequence** | | **TSE sequence** | |
| Visible in sequence | Yes | | Yes | |
| | **Reader 1** | **Reader 2** | **Reader 1** | **Reader 2** |
| **Detected** | No | No | Yes | Yes |

Abbreviations: Multiple sclerosis (MS), Turbo Field Echo (TFE), Turbo Spin Echo (TSE), Susceptibility Weighted Imaging with phase enhancement (SWIp).

#### LESION 195_05

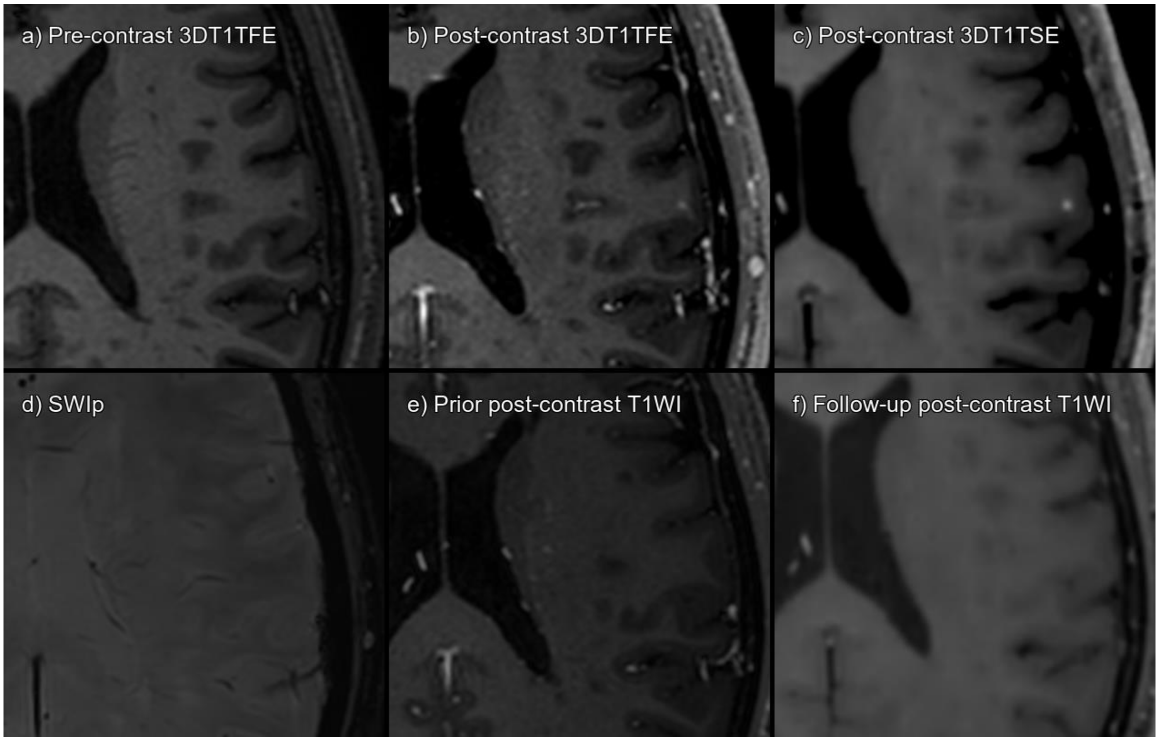


Supplementary figure 46. Lesion 195_05 a) 3DT1TFE without contrast, b) 3DT1TFE post-contrast, c) 3DT1TSE post-contrast, d) SWIp phase, e) Prior post-contrast T1-weighted imaging 1 year pre-baseline, f) Follow-up post-contrast T1-weighted imaging 9 months post-baseline.

True enhancing lesion that has disappeared on follow-up MRI.

| **CONSENSUS** | **True Enhancing MS Lesion** | | | |
|---|---|---|---|---|
| | **TFE sequence** | | **TSE sequence** | |
| Visible in sequence | Yes | | Yes | |
| | **Reader 1** | **Reader 2** | **Reader 1** | **Reader 2** |
| **Detected** | No | No | Yes | Yes |

Abbreviations: Multiple sclerosis (MS), Turbo Field Echo (TFE), Turbo Spin Echo (TSE), Susceptibility Weighted Imaging with phase enhancement (SWIp).

#### LESION 197_01

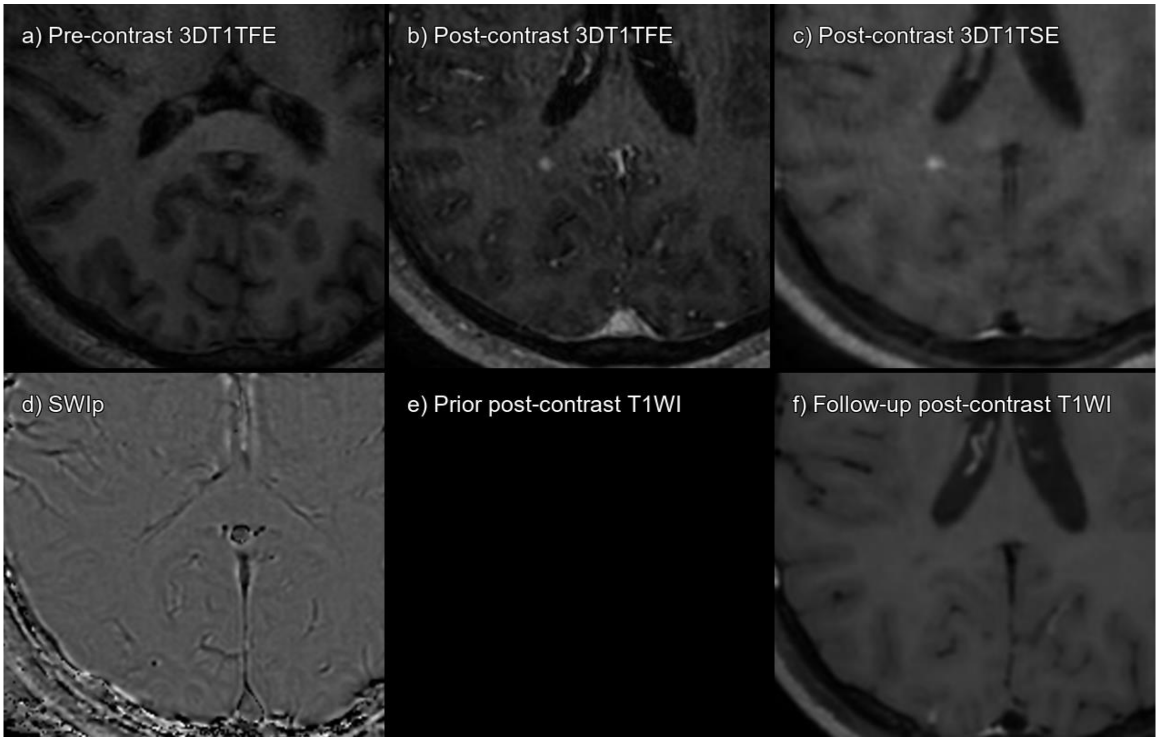


Supplementary figure 47. Lesion 197_01 a) 3DT1TFE without contrast, b) 3DT1TFE post-contrast, c) 3DT1TSE post-contrast, d) SWIp phase, e) Prior post-contrast T1-weighted imaging not available, f) Follow-up post-contrast T1-weighted imaging 1 year post-baseline.

True enhancing lesion that has disappeared on follow-up MRI.

| **CONSENSUS** | **True Enhancing MS Lesion** | | | |
|---|---|---|---|---|
| | **TFE sequence** | | **TSE sequence** | |
| Visible in sequence | Yes | | Yes | |
| | **Reader 1** | **Reader 2** | **Reader 1** | **Reader 2** |
| **Detected** | Yes | Yes | Yes | Yes |

Abbreviations: Multiple sclerosis (MS), Turbo Field Echo (TFE), Turbo Spin Echo (TSE), Susceptibility Weighted Imaging with phase enhancement (SWIp).

#### LESION 197_02

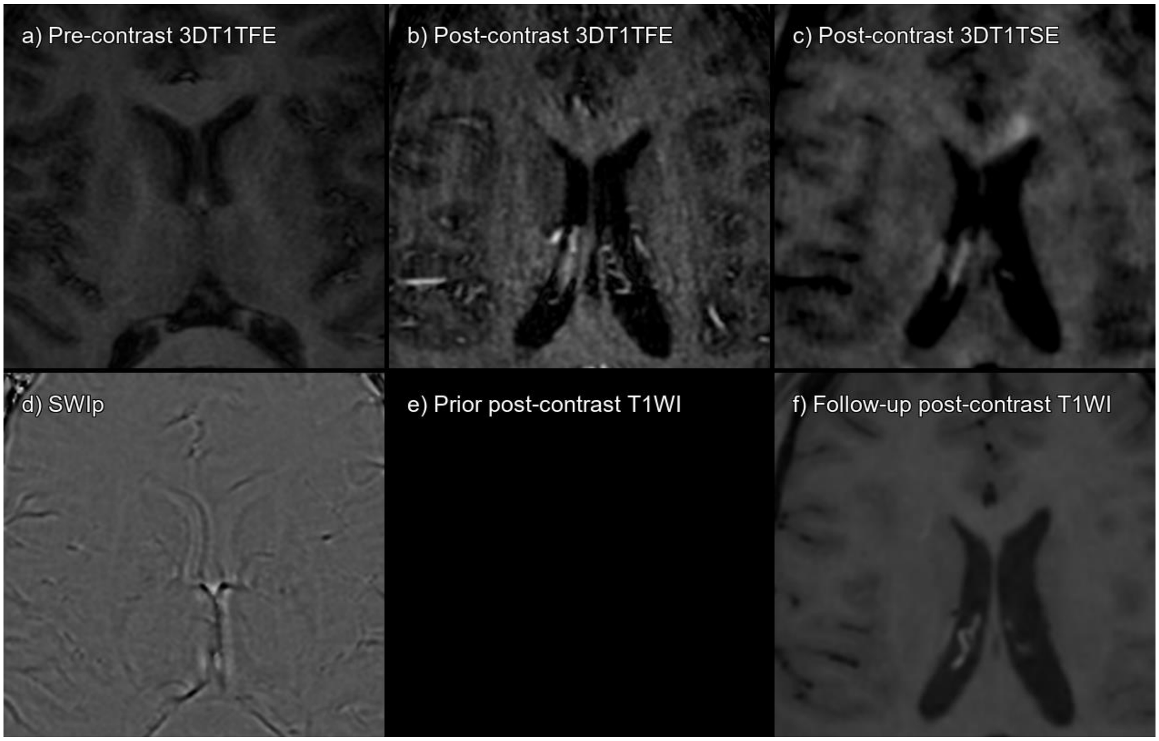


Supplementary figure 48. Lesion 197_02 a) 3DT1TFE without contrast, b) 3DT1TFE post-contrast, c) 3DT1TSE post-contrast, d) SWIp phase, e) Prior post-contrast T1-weighted imaging not available, f) Follow-up post-contrast T1-weighted imaging 1 year post-baseline.

True enhancing lesion that has disappeared on follow-up MRI. Not clearly visible on 3DT1TFE, but in retrospect and together with 3DT1TSE there is a subtle enhancement.

| **CONSENSUS** | **True Enhancing MS Lesion** | | | |
|---|---|---|---|---|
| | **TFE sequence** | | **TSE sequence** | |
| Visible in sequence | Yes | | Yes | |
| | **Reader 1** | **Reader 2** | **Reader 1** | **Reader 2** |
| **Detected** | No | No | Yes | Yes |

Abbreviations: Multiple sclerosis (MS), Turbo Field Echo (TFE), Turbo Spin Echo (TSE), Susceptibility Weighted Imaging with phase enhancement (SWIp).

#### LESION 197_03

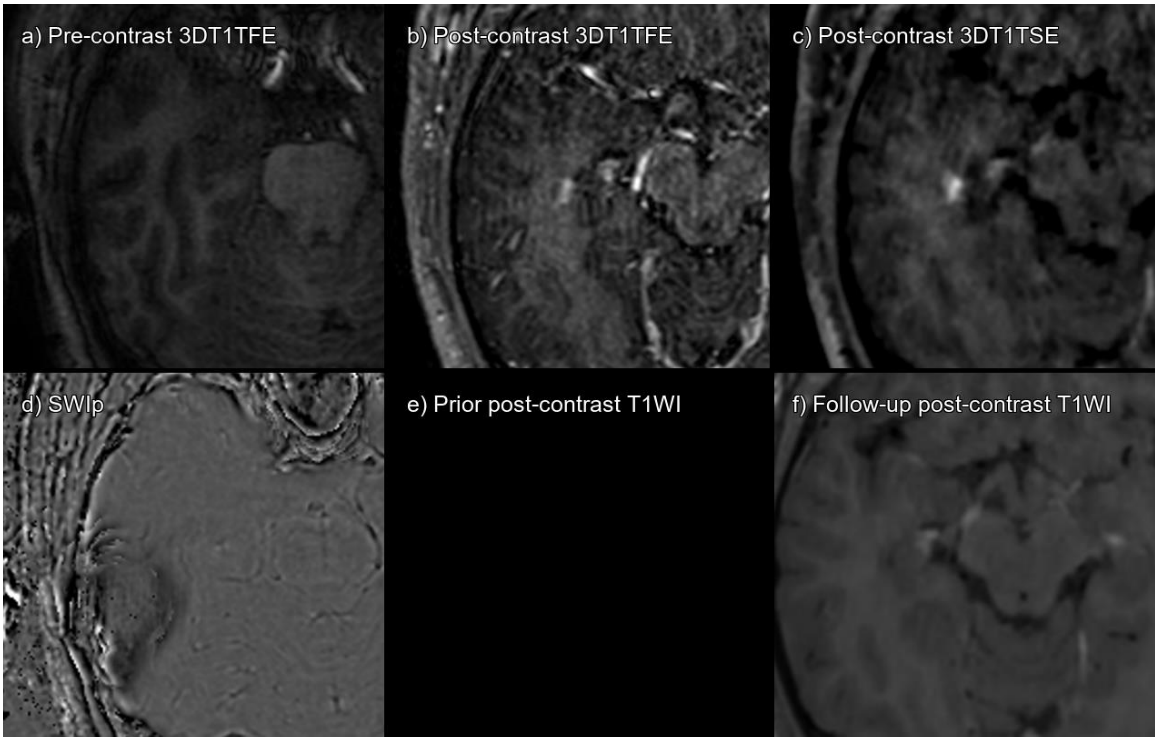


Supplementary figure 49. Lesion 197_03 a) 3DT1TFE without contrast, b) 3DT1TFE post-contrast, c) 3DT1TSE post-contrast, d) SWIp phase, e) Prior post-contrast T1-weighted imaging not available, f) Follow-up post-contrast T1-weighted imaging 1 year post-baseline.

True enhancing lesion that has disappeared on follow-up MRI.

| **CONSENSUS** | **True Enhancing MS Lesion** | | | |
|---|---|---|---|---|
| | **TFE sequence** | | **TSE sequence** | |
| Visible in sequence | Yes | | Yes | |
| | **Reader 1** | **Reader 2** | **Reader 1** | **Reader 2** |
| **Detected** | Yes | Yes | Yes | Yes |

Abbreviations: Multiple sclerosis (MS), Turbo Field Echo (TFE), Turbo Spin Echo (TSE), Susceptibility Weighted Imaging with phase enhancement (SWIp).

#### LESION 197_04

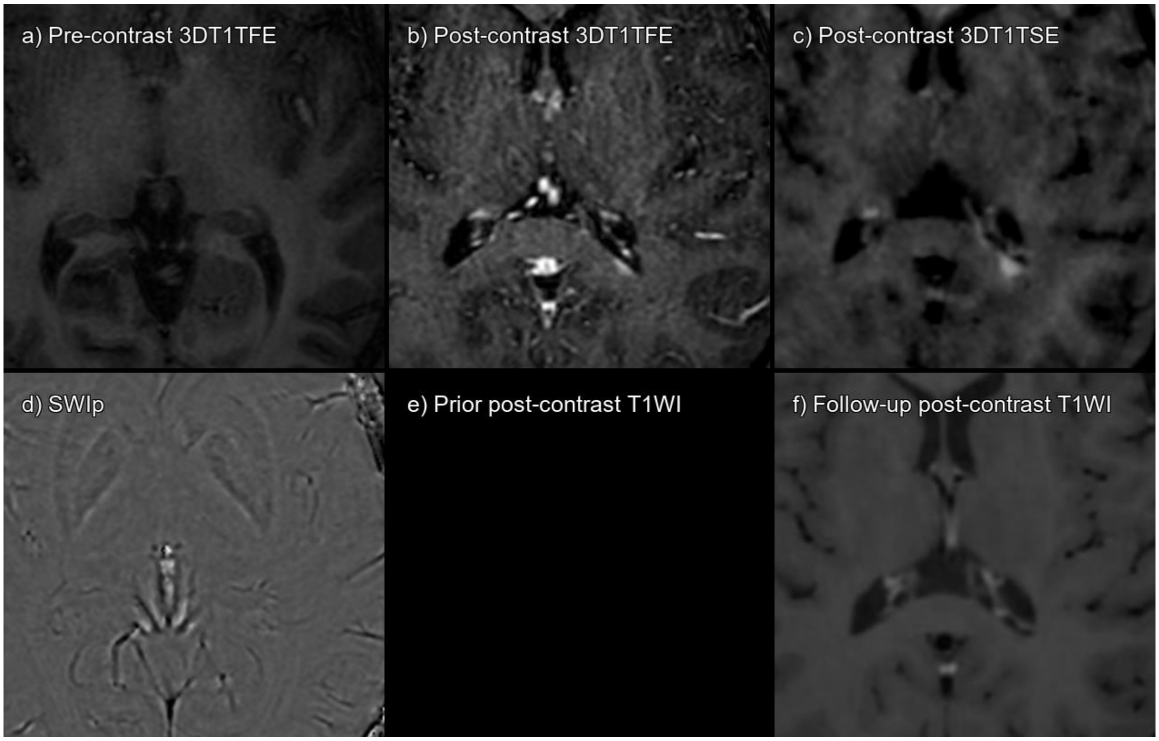


Supplementary figure 50. Lesion 197_04 a) 3DT1TFE without contrast, b) 3DT1TFE post-contrast, c) 3DT1TSE post-contrast, d) SWIp phase, e) Prior post-contrast T1-weighted imaging not available, f) Follow-up post-contrast T1-weighted imaging 1 year post-baseline.

True enhancing lesion that has disappeared on follow-up MRI.

| **CONSENSUS** | **True Enhancing MS Lesion** | | | |
|---|---|---|---|---|
| | **TFE sequence** | | **TSE sequence** | |
| Visible in sequence | Yes | | Yes | |
| | **Reader 1** | **Reader 2** | **Reader 1** | **Reader 2** |
| **Detected** | No | No | Yes | Yes |

Abbreviations: Multiple sclerosis (MS), Turbo Field Echo (TFE), Turbo Spin Echo (TSE), Susceptibility Weighted Imaging with phase enhancement (SWIp).

#### LESION 210_01

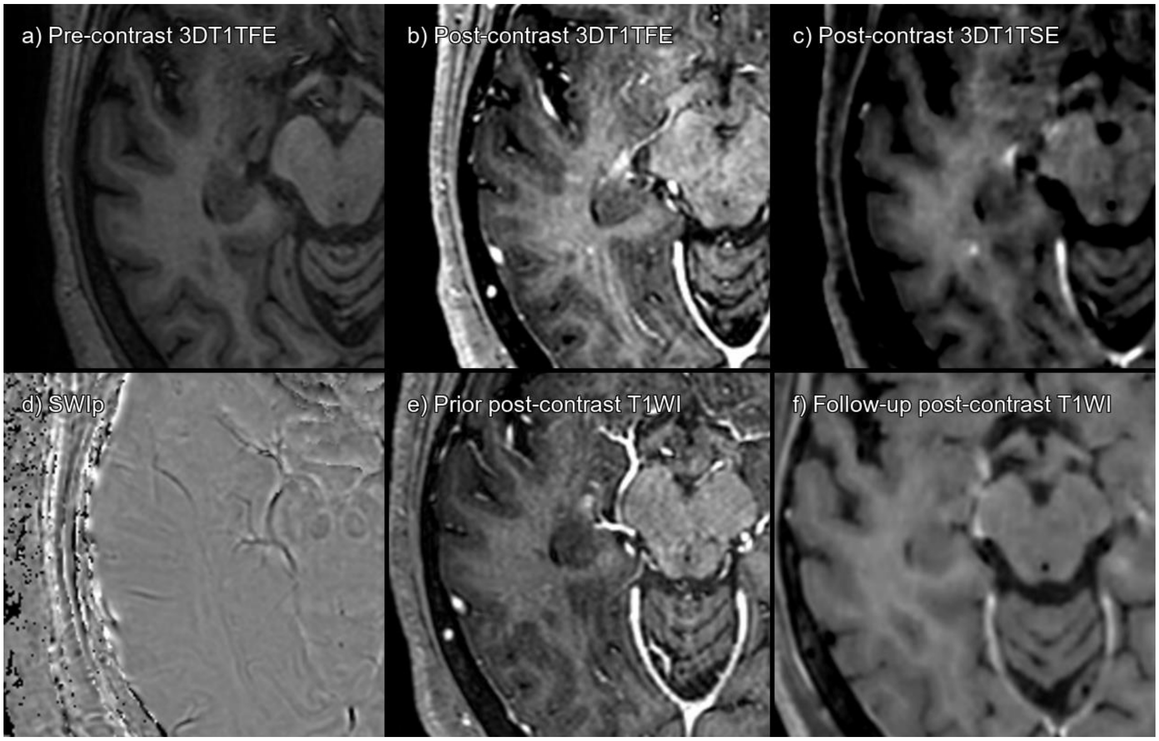


Supplementary figure 51. Lesion 210_01 a) 3DT1TFE without contrast, b) 3DT1TFE post-contrast, c) 3DT1TSE post-contrast, d) SWIp phase, e) Prior post-contrast T1-weighted imaging 1 year pre-baseline, f) Follow-up post-contrast T1-weighted imaging 1 year post-baseline.

True enhancing lesion that has disappeared on follow-up MRI.

| **CONSENSUS** | **True Enhancing MS Lesion** | | | |
|---|---|---|---|---|
| | **TFE sequence** | | **TSE sequence** | |
| Visible in sequence | Yes | | Yes | |
| | **Reader 1** | **Reader 2** | **Reader 1** | **Reader 2** |
| **Detected** | No | No | No | Yes |

Abbreviations: Multiple sclerosis (MS), Turbo Field Echo (TFE), Turbo Spin Echo (TSE), Susceptibility Weighted Imaging with phase enhancement (SWIp).

#### LESION 210_02

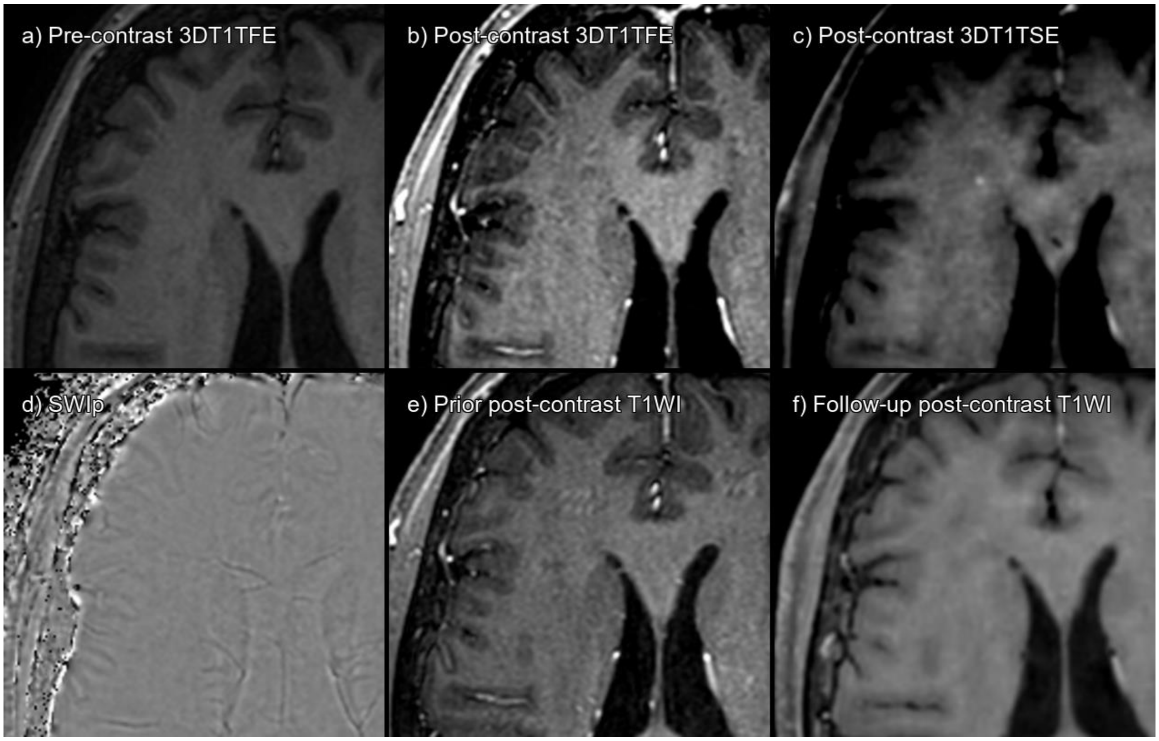


Supplementary figure 52. Lesion 210_02 a) 3DT1TFE without contrast, b) 3DT1TFE post-contrast, c) 3DT1TSE post-contrast, d) SWIp phase, e) Prior post-contrast T1-weighted imaging 1 year pre-baseline, f) Follow-up post-contrast T1-weighted imaging 1 year post-baseline.

True enhancing lesion, visible on 3DT1TSE but not on 3DT1TFE, that has disappeared on follow-up MRI.

| **CONSENSUS** | **True Enhancing MS Lesion** | | | |
|---|---|---|---|---|
| | **TFE sequence** | | **TSE sequence** | |
| Visible in sequence | No | | Yes | |
| | **Reader 1** | **Reader 2** | **Reader 1** | **Reader 2** |
| **Detected** | No | No | No | Yes |

Abbreviations: Multiple sclerosis (MS), Turbo Field Echo (TFE), Turbo Spin Echo (TSE), Susceptibility Weighted Imaging with phase enhancement (SWIp).

### LESION 211_01

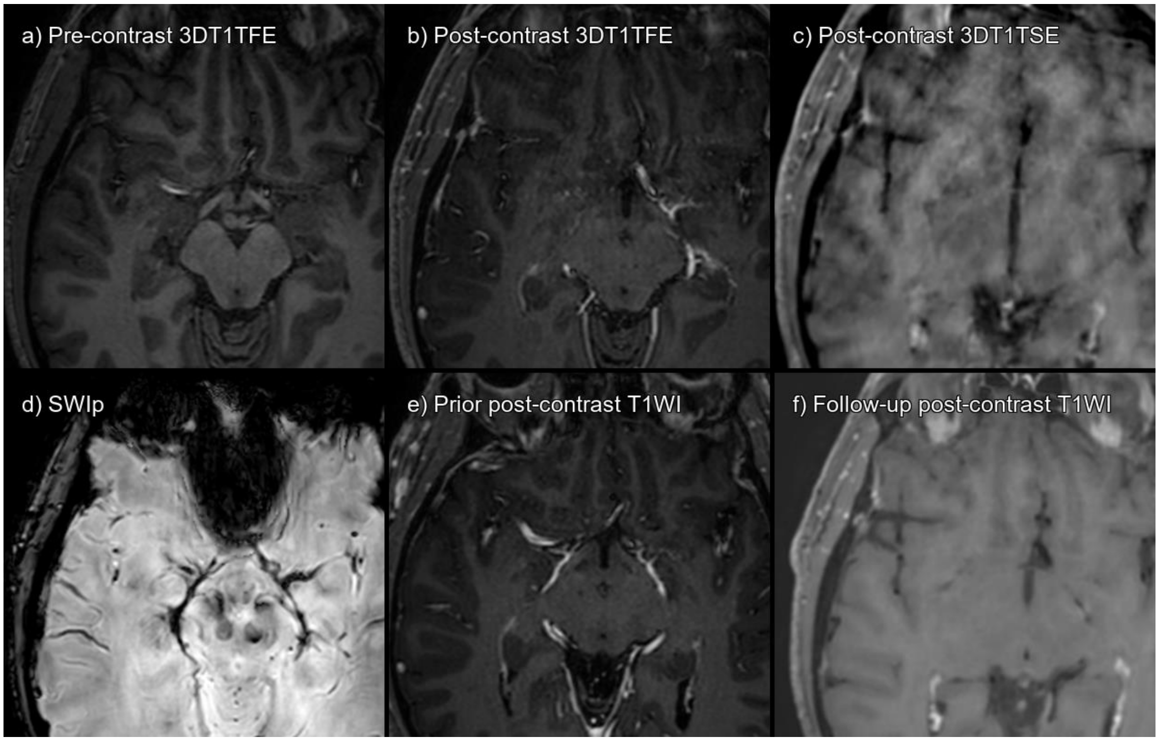


Supplementary figure 53. Lesion 211_01 a) 3DT1TFE without contrast, b) 3DT1TFE post-contrast, c) 3DT1TSE post-contrast, d) SWIp phase, e) Prior post-contrast T1-weighted imaging 1 year pre-baseline, f) Follow-up post-contrast T1-weighted imaging 1 year post-baseline.

There is no enhancement but a subtle artefactual subinsular hyperintensity on baseline 3DT1TSE mistaken for enhancement by one reader.

| **CONSENSUS** | **Not acute MS lesion enhancement** | | | |
|---|---|---|---|---|
| | **TFE sequence** | | **TSE sequence** | |
| Visible in sequence | No | | No | |
| | **Reader 1** | **Reader 2** | **Reader 1** | **Reader 2** |
| **Detected** | No | No | No | Yes |

Abbreviations: Multiple sclerosis (MS), Turbo Field Echo (TFE), Turbo Spin Echo (TSE), Susceptibility Weighted Imaging with phase enhancement (SWIp).

#### LESION 215_01

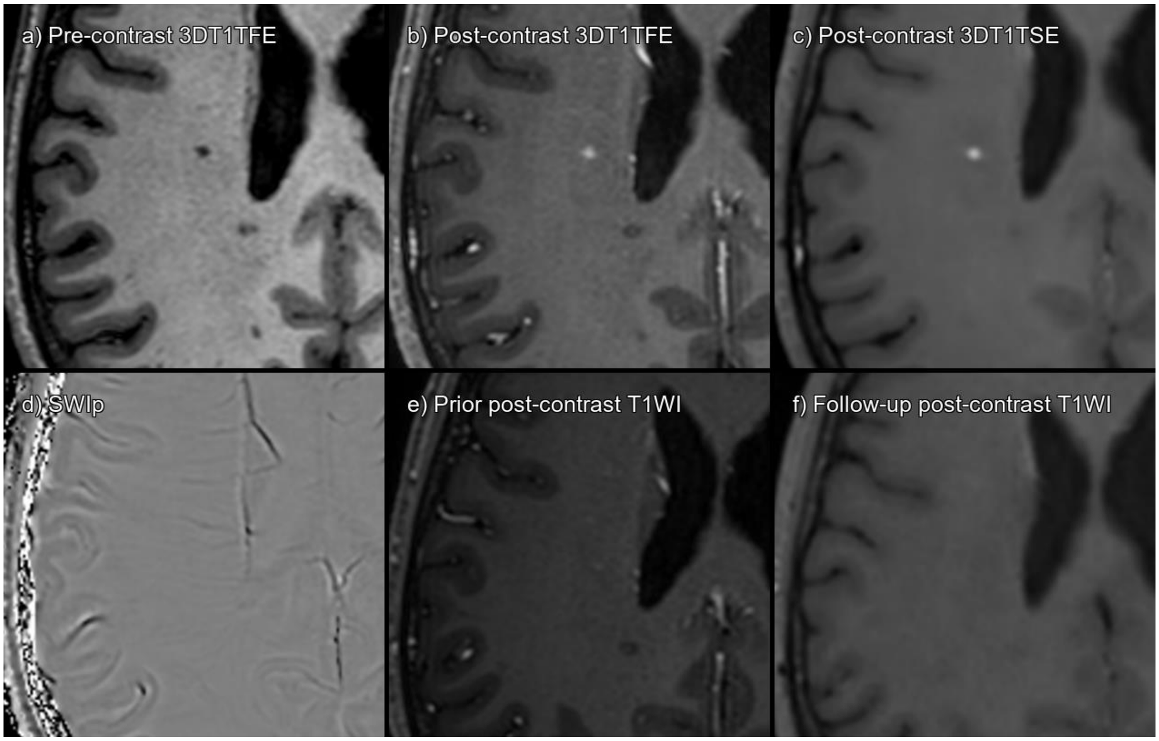


Supplementary figure 54. Lesion 215_01 a) 3DT1TFE without contrast, b) 3DT1TFE post-contrast, c) 3DT1TSE post-contrast, d) SWIp phase, e) Prior post-contrast T1-weighted imaging 1 year pre-baseline, f) Follow-up post-contrast T1-weighted imaging 1 year post-baseline.

True enhancing lesion that has disappeared on follow-up MRI.

| **CONSENSUS** | **True Enhancing MS Lesion** | | | |
|---|---|---|---|---|
| | **TFE sequence** | | **TSE sequence** | |
| Visible in sequence | Yes | | Yes | |
| | **Reader 1** | **Reader 2** | **Reader 1** | **Reader 2** |
| **Detected** | Yes | Yes | Yes | Yes |

Abbreviations: Multiple sclerosis (MS), Turbo Field Echo (TFE), Turbo Spin Echo (TSE), Susceptibility Weighted Imaging with phase enhancement (SWIp).

#### LESION 215_02

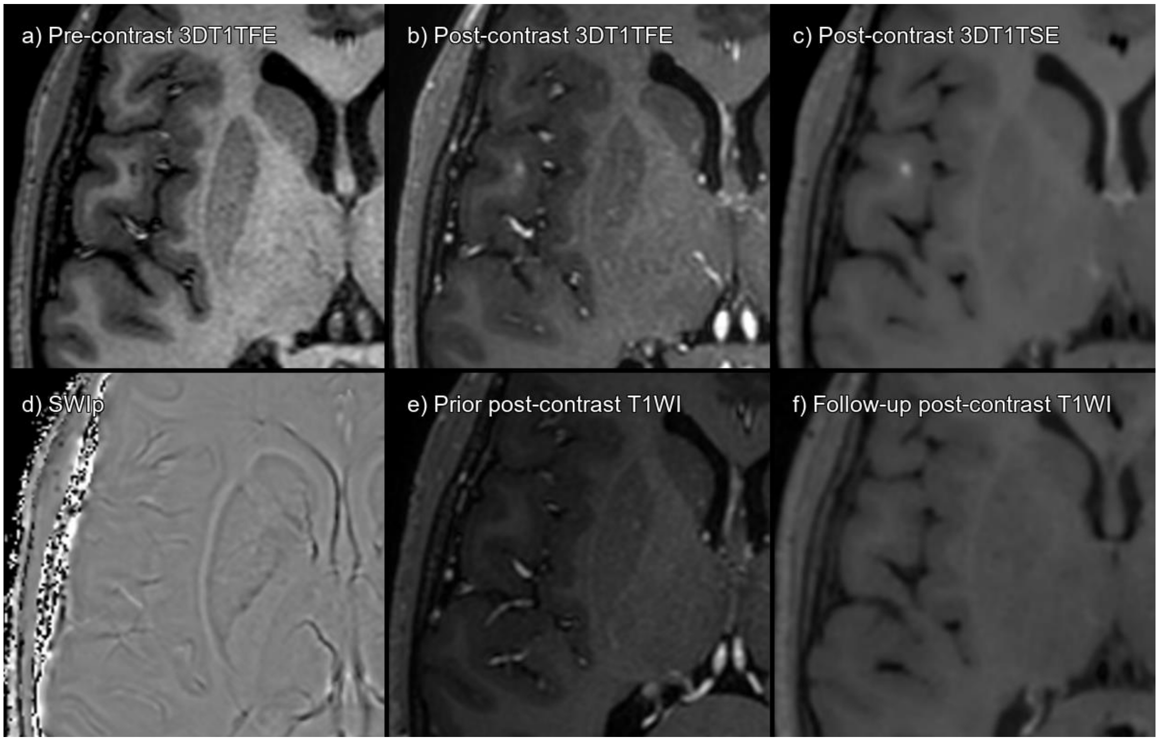


Supplementary figure 55. Lesion 215_02 a) 3DT1TFE without contrast, b) 3DT1TFE post-contrast, c) 3DT1TSE post-contrast, d) SWIp phase, e) Prior post-contrast T1-weighted imaging 1 year pre-baseline, f) Follow-up post-contrast T1-weighted imaging 1 year post-baseline.

True enhancing lesion that has disappeared on follow-up MRI.

| **CONSENSUS** | **True Enhancing MS Lesion** | | | |
|---|---|---|---|---|
| | **TFE sequence** | | **TSE sequence** | |
| Visible in sequence | Yes | | Yes | |
| | **Reader 1** | **Reader 2** | **Reader 1** | **Reader 2** |
| **Detected** | No | No | Yes | Yes |

Abbreviations: Multiple sclerosis (MS), Turbo Field Echo (TFE), Turbo Spin Echo (TSE), Susceptibility Weighted Imaging with phase enhancement (SWIp).

#### LESION 215_03

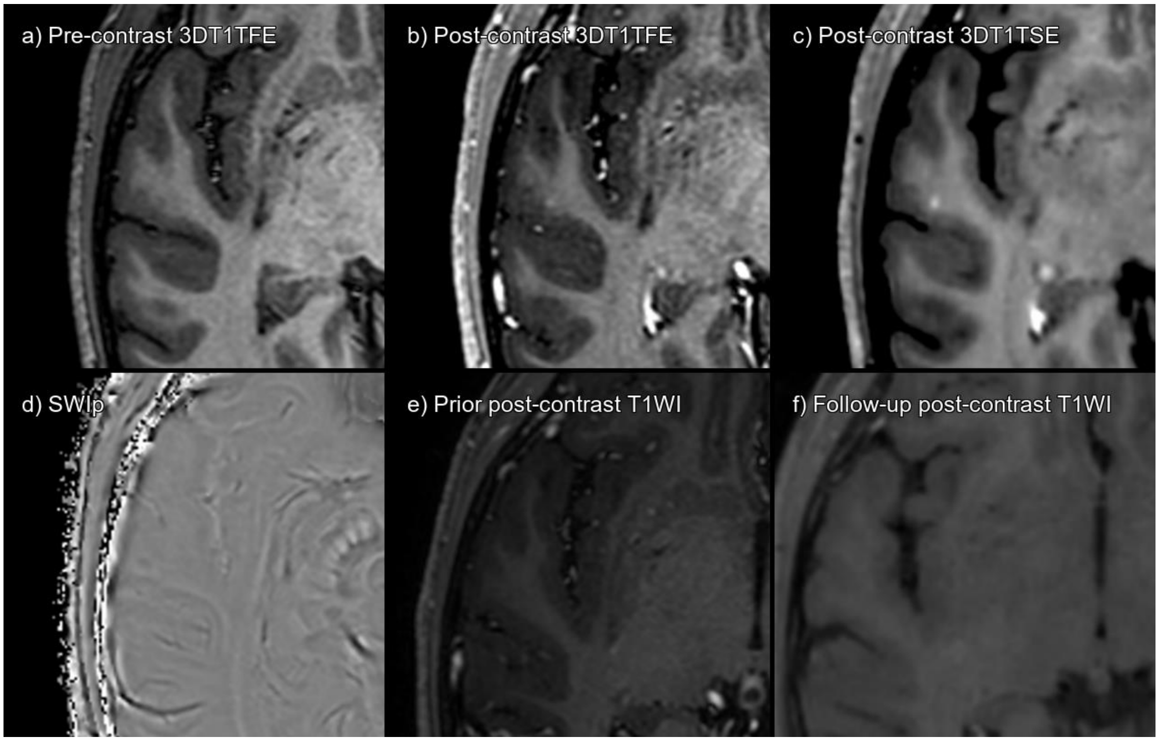


Supplementary figure 56. Lesion 215_03 a) 3DT1TFE without contrast, b) 3DT1TFE post-contrast, c) 3DT1TSE post-contrast, d) SWIp phase, e) Prior post-contrast T1-weighted imaging 1 year pre-baseline, f) Follow-up post-contrast T1-weighted imaging 1 year post-baseline.

True enhancing lesion that has disappeared on follow-up MRI.

| **CONSENSUS** | **True Enhancing MS Lesion** | | | |
|---|---|---|---|---|
| | **TFE sequence** | | **TSE sequence** | |
| Visible in sequence | Yes | | Yes | |
| | **Reader 1** | **Reader 2** | **Reader 1** | **Reader 2** |
| **Detected** | No | No | No | Yes |

Abbreviations: Multiple sclerosis (MS), Turbo Field Echo (TFE), Turbo Spin Echo (TSE), Susceptibility Weighted Imaging with phase enhancement (SWIp).

#### LESION 242_01

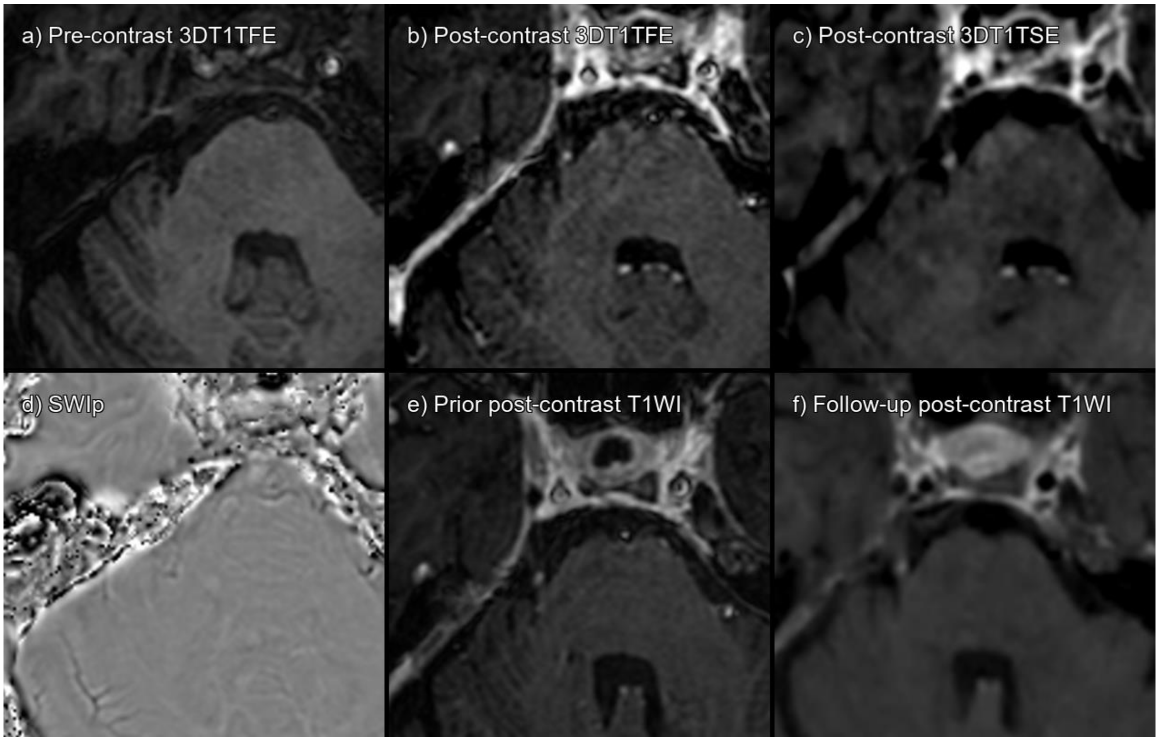


Supplementary figure 57. Lesion 242_01 a) 3DT1TFE without contrast, b) 3DT1TFE post-contrast, c) 3DT1TSE post-contrast, d) SWIp phase, e) Prior post-contrast T1-weighted imaging 6 months pre-baseline, f) Follow-up post-contrast T1-weighted imaging 6 months post-baseline.

Focal enhancement of the right trigeminal nerve, already present on pre- and post-baseline examinations. False positive, probably vascular enhancement.

| **CONSENSUS** | **Not acute MS lesion enhancement** | | | |
|---|---|---|---|---|
| | **TFE sequence** | | **TSE sequence** | |
| Visible in sequence | Yes | | Yes | |
| | **Reader 1** | **Reader 2** | **Reader 1** | **Reader 2** |
| **Detected** | No | No | Yes | No |

Abbreviations: Multiple sclerosis (MS), Turbo Field Echo (TFE), Turbo Spin Echo (TSE), Susceptibility Weighted Imaging with phase enhancement (SWIp).

#### LESION 247_01

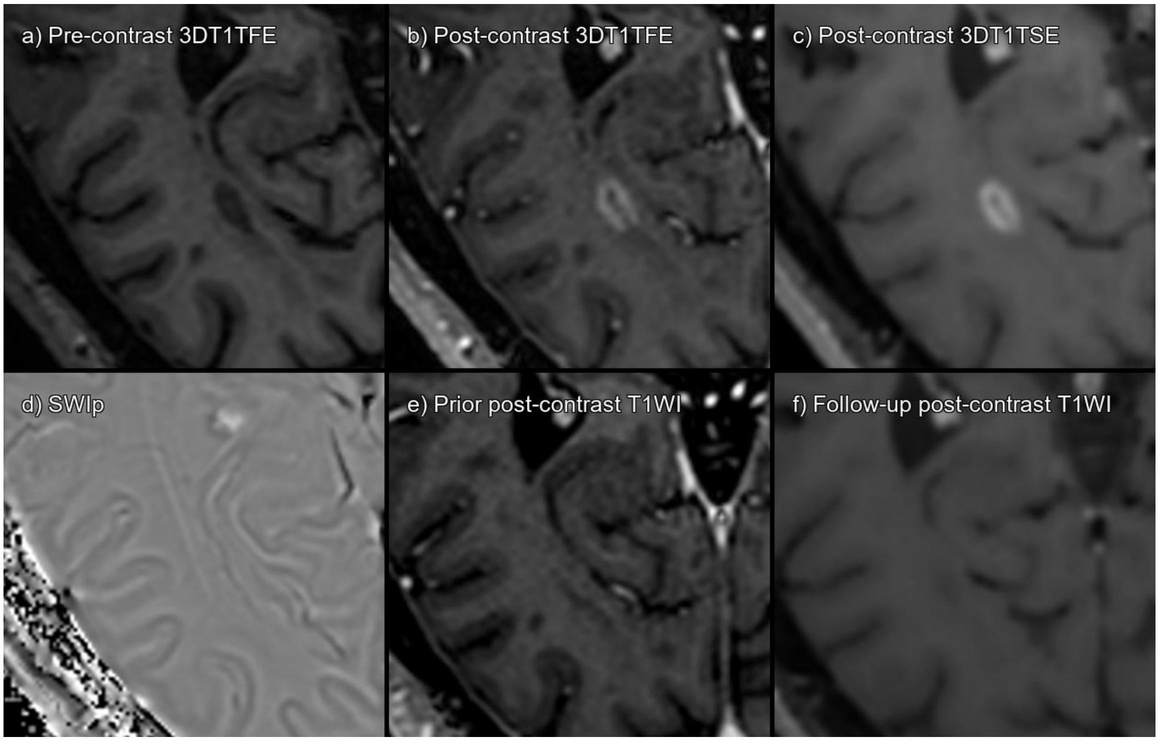


Supplementary figure 58. Lesion 247_01 a) 3DT1TFE without contrast, b) 3DT1TFE post-contrast, c) 3DT1TSE post-contrast, d) SWIp phase, e) Prior post-contrast T1-weighted imaging 2 years pre-baseline, f) Follow-up post-contrast T1-weighted imaging 6 months post-baseline.

True enhancing lesion that has disappeared on follow-up MRI.

| **CONSENSUS** | **True Enhancing MS Lesion** | | | |
|---|---|---|---|---|
| | **TFE sequence** | | **TSE sequence** | |
| Visible in sequence | Yes | | Yes | |
| | **Reader 1** | **Reader 2** | **Reader 1** | **Reader 2** |
| **Detected** | Yes | Yes | Yes | Yes |

Abbreviations: Multiple sclerosis (MS), Turbo Field Echo (TFE), Turbo Spin Echo (TSE), Susceptibility Weighted Imaging with phase enhancement (SWIp).

#### LESION 247_02

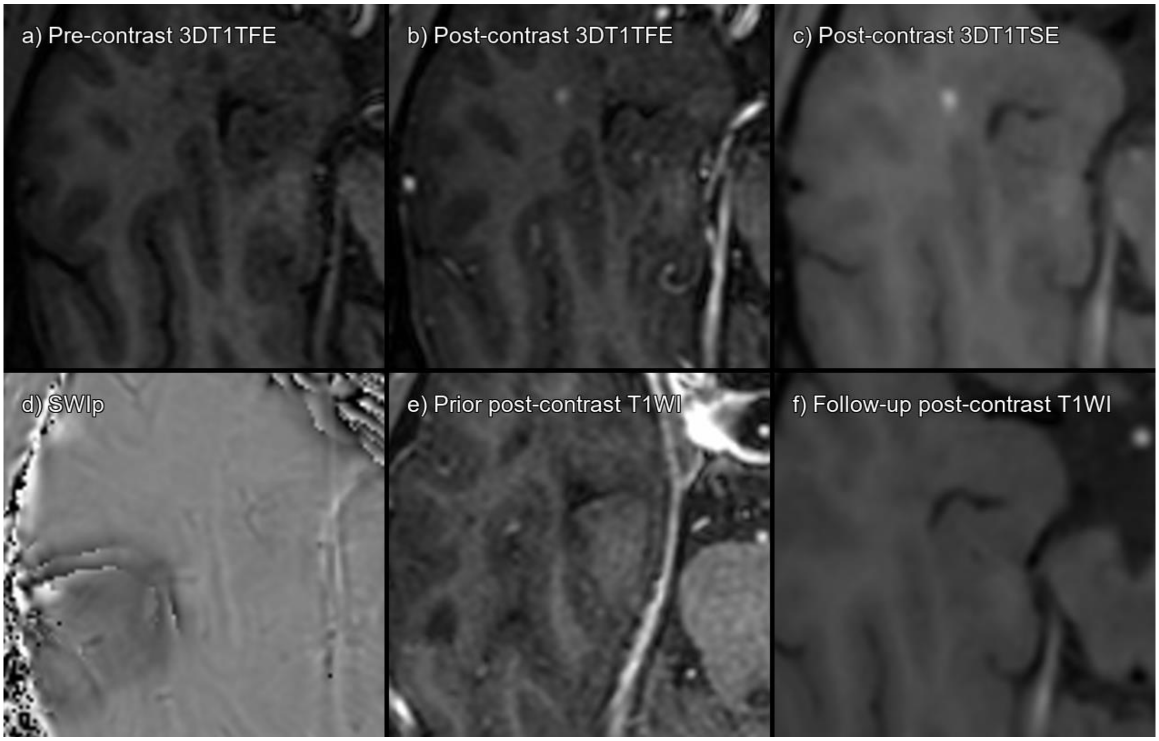


Supplementary figure 59. Lesion 247_02 a) 3DT1TFE without contrast, b) 3DT1TFE post-contrast, c) 3DT1TSE post-contrast, d) SWIp phase, e) Prior post-contrast T1-weighted imaging 2 years pre-baseline, f) Follow-up post-contrast T1-weighted imaging 6 months post-baseline.

True enhancing lesion that has disappeared on follow-up MRI.

| **CONSENSUS** | **True Enhancing MS Lesion** | | | |
|---|---|---|---|---|
| | **TFE sequence** | | **TSE sequence** | |
| Visible in sequence | Yes | | Yes | |
| | **Reader 1** | **Reader 2** | **Reader 1** | **Reader 2** |
| **Detected** | No | Yes | Yes | No |

Abbreviations: Multiple sclerosis (MS), Turbo Field Echo (TFE), Turbo Spin Echo (TSE), Susceptibility Weighted Imaging with phase enhancement (SWIp).

#### LESION 247_03

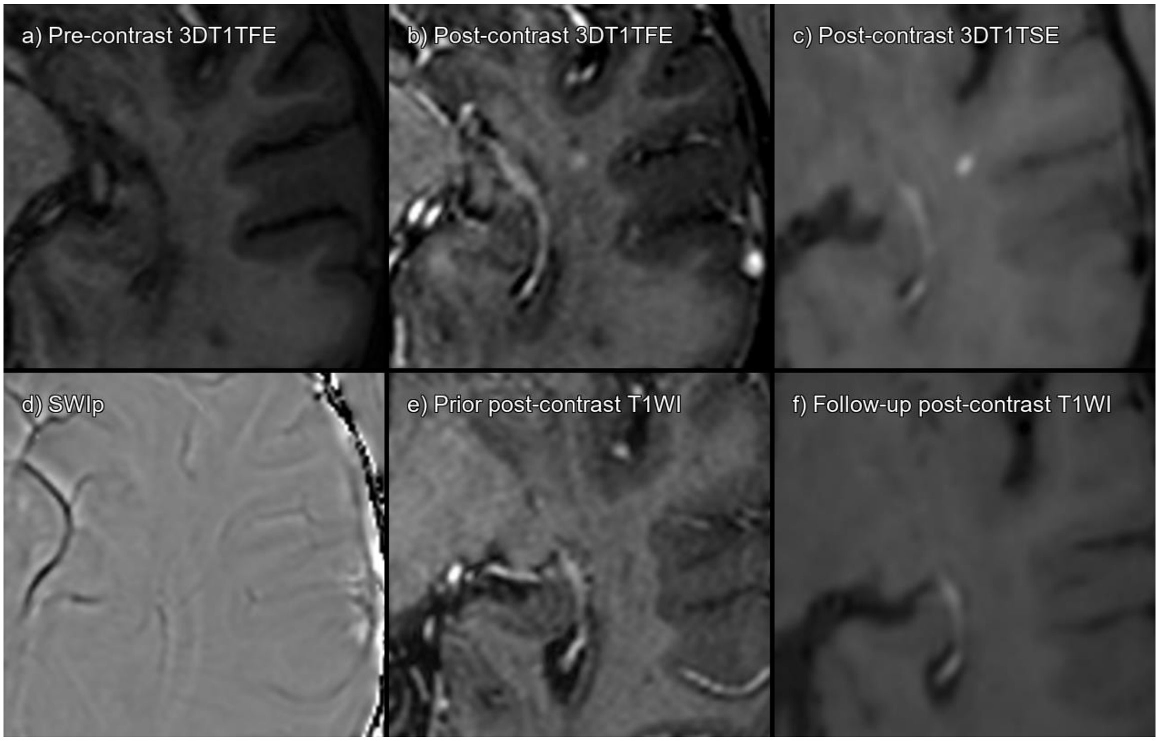


Supplementary figure 60. Lesion 247_03 a) 3DT1TFE without contrast, b) 3DT1TFE post-contrast, c) 3DT1TSE post-contrast, d) SWIp phase, e) Prior post-contrast T1-weighted imaging 2 years pre-baseline, f) Follow-up post-contrast T1-weighted imaging 6 months post-baseline.

True enhancing lesion that has disappeared on follow-up MRI.

| **CONSENSUS** | **True Enhancing MS Lesion** | | | |
|---|---|---|---|---|
| | **TFE sequence** | | **TSE sequence** | |
| Visible in sequence | Yes | | Yes | |
| | **Reader 1** | **Reader 2** | **Reader 1** | **Reader 2** |
| **Detected** | No | Yes | Yes | Yes |

Abbreviations: Multiple sclerosis (MS), Turbo Field Echo (TFE), Turbo Spin Echo (TSE), Susceptibility Weighted Imaging with phase enhancement (SWIp).

#### LESION 247_04

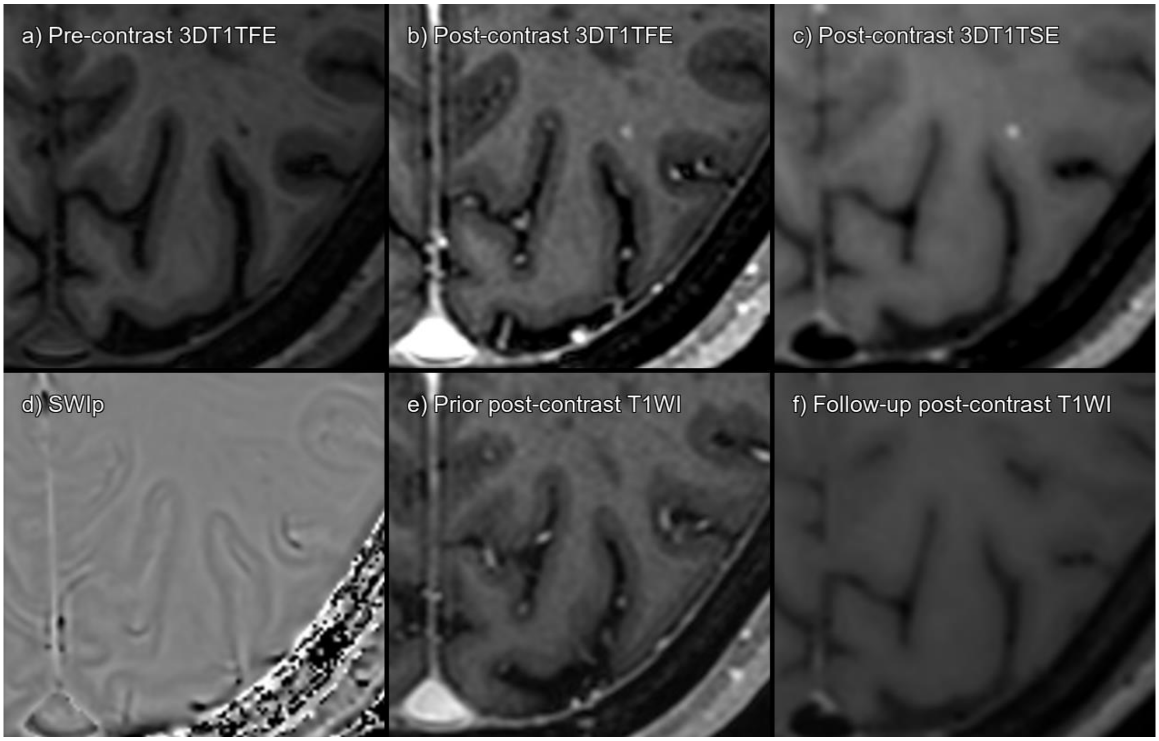


Supplementary figure 61. Lesion 247_04 a) 3DT1TFE without contrast, b) 3DT1TFE post-contrast, c) 3DT1TSE post-contrast, d) SWIp phase, e) Prior post-contrast T1-weighted imaging 2 years pre-baseline, f) Follow-up post-contrast T1-weighted imaging 6 months post-baseline.

True enhancing lesion that has disappeared on follow-up MRI.

| **CONSENSUS** | **True Enhancing MS Lesion** | | | |
|---|---|---|---|---|
| | **TFE sequence** | | **TSE sequence** | |
| Visible in sequence | Yes | | Yes | |
| | **Reader 1** | **Reader 2** | **Reader 1** | **Reader 2** |
| **Detected** | No | No | Yes | Yes |

Abbreviations: Multiple sclerosis (MS), Turbo Field Echo (TFE), Turbo Spin Echo (TSE), Susceptibility Weighted Imaging with phase enhancement (SWIp).

#### LESION 247_05

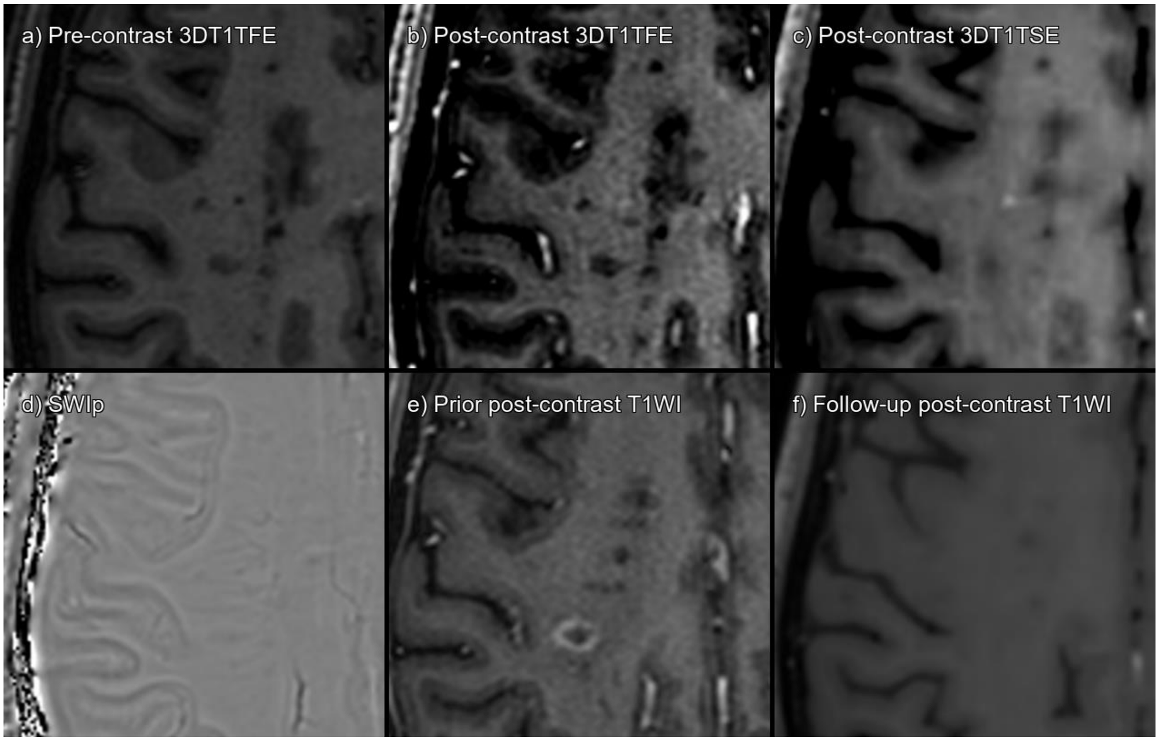


Supplementary figure 62. Lesion 247_05 a) 3DT1TFE without contrast, b) 3DT1TFE post-contrast, c) 3DT1TSE post-contrast, d) SWIp phase, e) Prior post-contrast T1-weighted imaging 2 years pre-baseline, f) Follow-up post-contrast T1-weighted imaging 6 months post-baseline.

True enhancing lesion that has disappeared on follow-up MRI. Not visible on TFE. Note that the prior TFE had a different ring enhancing lesion, now resolved.

| **CONSENSUS** | **True Enhancing MS Lesion** | | | |
|---|---|---|---|---|
| | **TFE sequence** | | **TSE sequence** | |
| Visible in sequence | No | | Yes | |
| | **Reader 1** | **Reader 2** | **Reader 1** | **Reader 2** |
| **Detected** | No | No | Yes | No |

Abbreviations: Multiple sclerosis (MS), Turbo Field Echo (TFE), Turbo Spin Echo (TSE), Susceptibility Weighted Imaging with phase enhancement (SWIp).

#### LESION 247_06

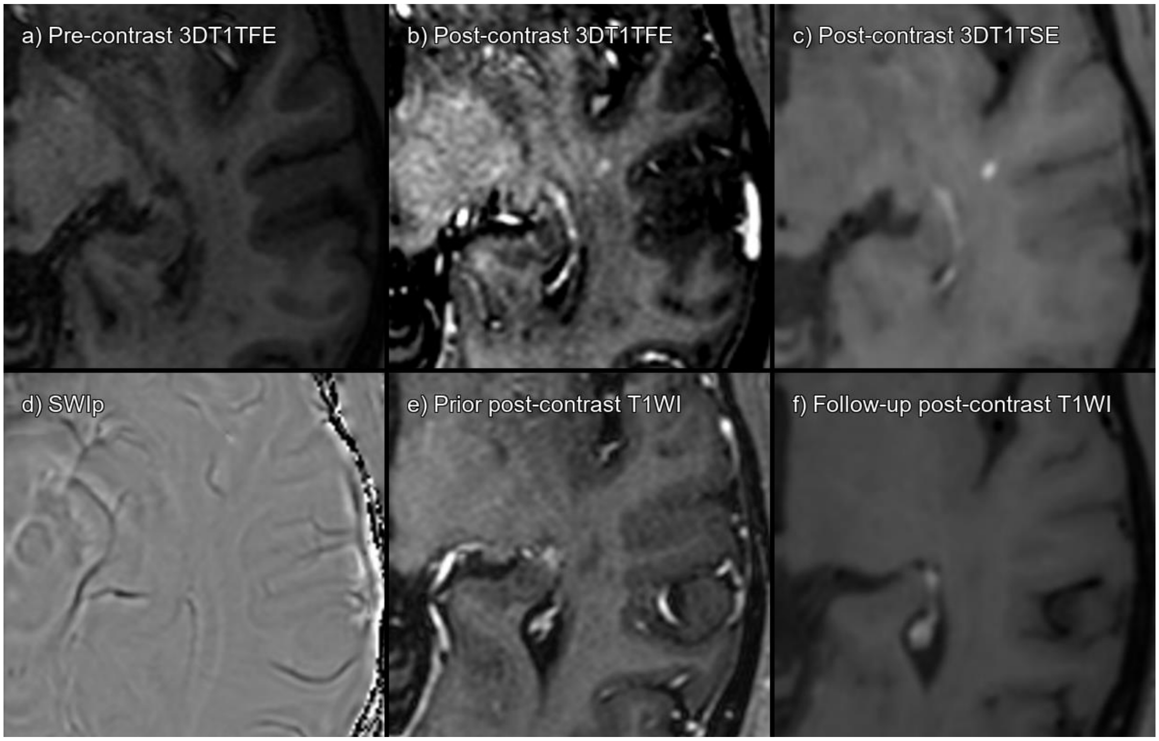


Supplementary figure 63. Lesion 247_06 a) 3DT1TFE without contrast, b) 3DT1TFE post-contrast, c) 3DT1TSE post-contrast, d) SWIp phase, e) Prior post-contrast T1-weighted imaging 2 years pre-baseline, f) Follow-up post-contrast T1-weighted imaging 6 months post-baseline.

True enhancing lesion that has disappeared on follow-up MRI. Very subtle enhancement on 3DT1TFE, but visible in retrospect.

| **CONSENSUS** | **True Enhancing MS Lesion** | | | |
|---|---|---|---|---|
| | **TFE sequence** | | **TSE sequence** | |
| Visible in sequence | Yes | | Yes | |
| | **Reader 1** | **Reader 2** | **Reader 1** | **Reader 2** |
| **Detected** | No | No | Yes | Yes |

Abbreviations: Multiple sclerosis (MS), Turbo Field Echo (TFE), Turbo Spin Echo (TSE), Susceptibility Weighted Imaging with phase enhancement (SWIp).

#### LESION 247_07

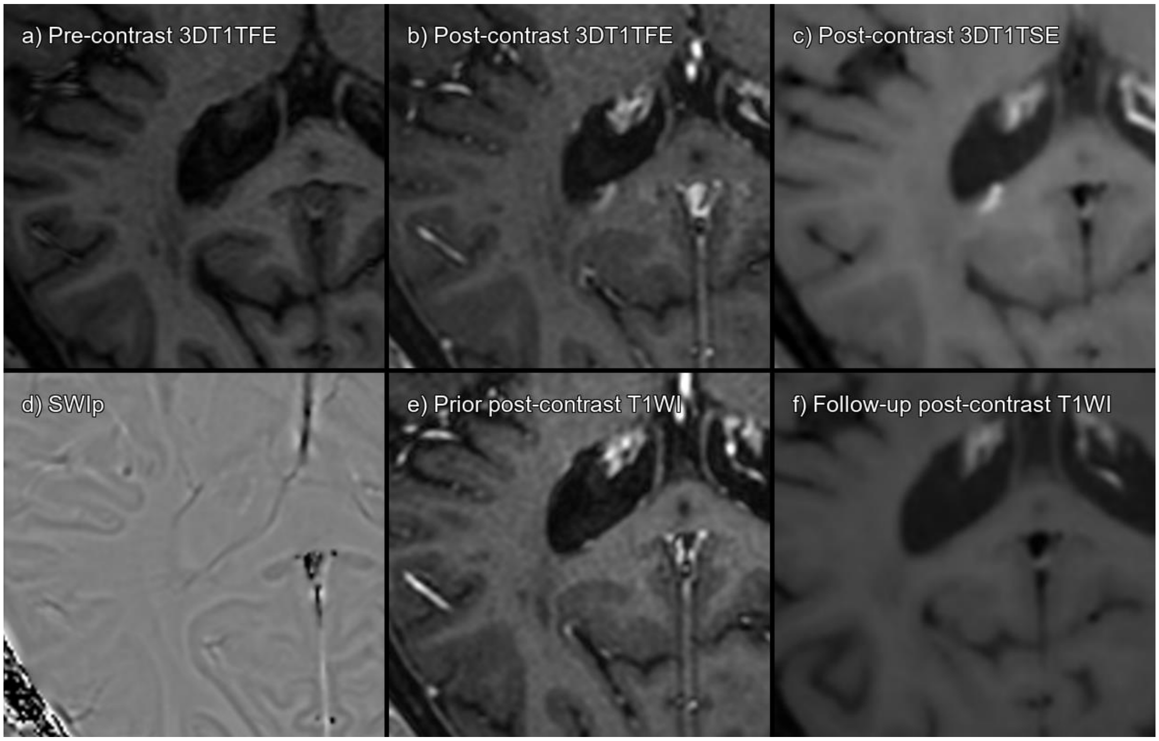


Supplementary figure 64. Lesion 247_07 a) 3DT1TFE without contrast, b) 3DT1TFE post-contrast, c) 3DT1TSE post-contrast, d) SWIp phase, e) Prior post-contrast T1-weighted imaging 2 years pre-baseline, f) Follow-up post-contrast T1-weighted imaging 6 months post-baseline.

True enhancing lesion that has disappeared on follow-up MRI.

| **CONSENSUS** | **True Enhancing MS Lesion** | | | |
|---|---|---|---|---|
| | **TFE sequence** | | **TSE sequence** | |
| Visible in sequence | Yes | | Yes | |
| | **Reader 1** | **Reader 2** | **Reader 1** | **Reader 2** |
| **Detected** | No | No | Yes | Yes |

Abbreviations: Multiple sclerosis (MS), Turbo Field Echo (TFE), Turbo Spin Echo (TSE), Susceptibility Weighted Imaging with phase enhancement (SWIp).

#### LESION 248_01

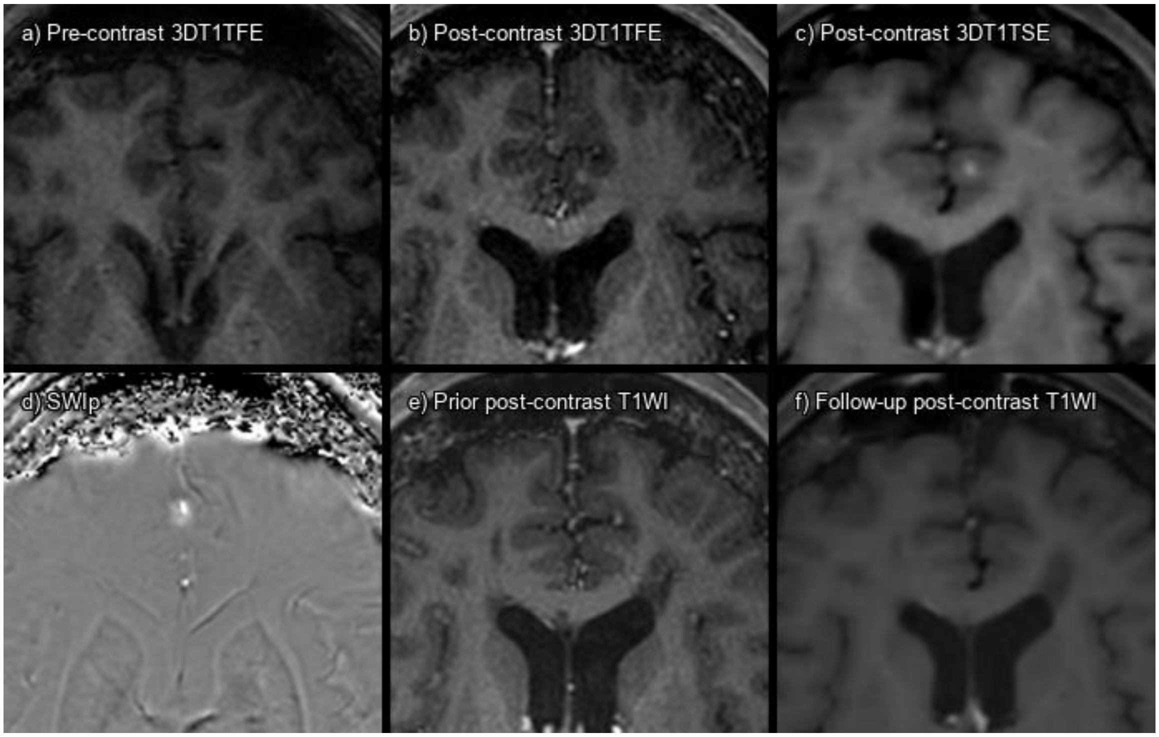


Supplementary figure 65. Lesion 248_01 a) 3DT1TFE without contrast, b) 3DT1TFE post-contrast, c) 3DT1TSE post-contrast, d) SWIp phase, e) Prior post-contrast T1-weighted imaging 3 months pre-baseline, f) Follow-up post-contrast T1-weighted imaging 4 months post-baseline.

True enhancing lesion that has disappeared on follow-up MRI. Very subtle enhancement on 3DT1TFE, but confirmed on comparative analysis.

| **CONSENSUS** | **True Enhancing MS Lesion** | | | |
|---|---|---|---|---|
| | **TFE sequence** | | **TSE sequence** | |
| Visible in sequence | Yes | | Yes | |
| | **Reader 1** | **Reader 2** | **Reader 1** | **Reader 2** |
| **Detected** | No | No | No | Yes |

Abbreviations: Multiple sclerosis (MS), Turbo Field Echo (TFE), Turbo Spin Echo (TSE), Susceptibility Weighted Imaging with phase enhancement (SWIp).

#### LESION 248_02

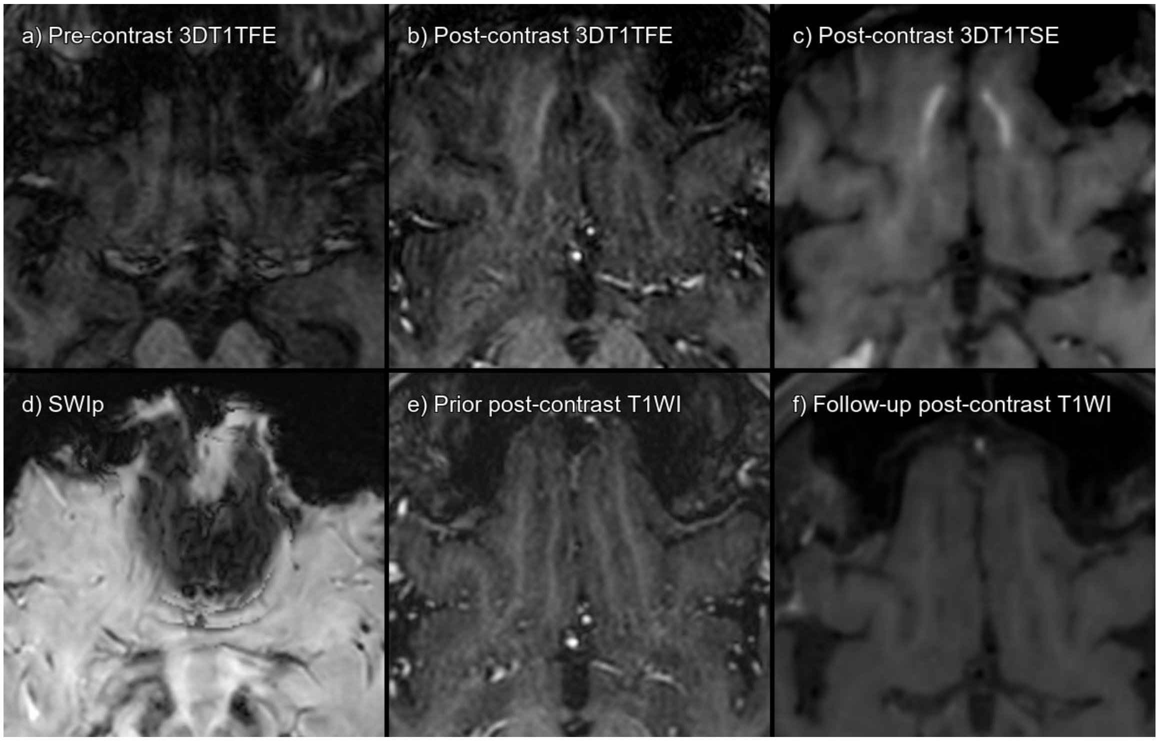


Supplementary figure 66. Lesion 248_02 a) 3DT1TFE without contrast, b) 3DT1TFE post-contrast, c) 3DT1TSE post-contrast, d) SWIp, e) Prior post-contrast T1-weighted imaging 3 months pre-baseline, f) Follow-up post-contrast T1-weighted imaging 4 months post-baseline.

True enhancing lesion that has disappeared on follow-up MRI. Lesion 248_02 refers to the right rectal gyrus lesion. This region is not discernible on SWIp due to strong susceptibility artifacts.

| **CONSENSUS** | **True Enhancing MS Lesion** | | | |
|---|---|---|---|---|
| | **TFE sequence** | | **TSE sequence** | |
| Visible in sequence | Yes | | Yes | |
| | **Reader 1** | **Reader 2** | **Reader 1** | **Reader 2** |
| **Detected** | No | No | No | Yes |

Abbreviations: Multiple sclerosis (MS), Turbo Field Echo (TFE), Turbo Spin Echo (TSE), Susceptibility Weighted Imaging with phase enhancement (SWIp).

#### LESION 248_03

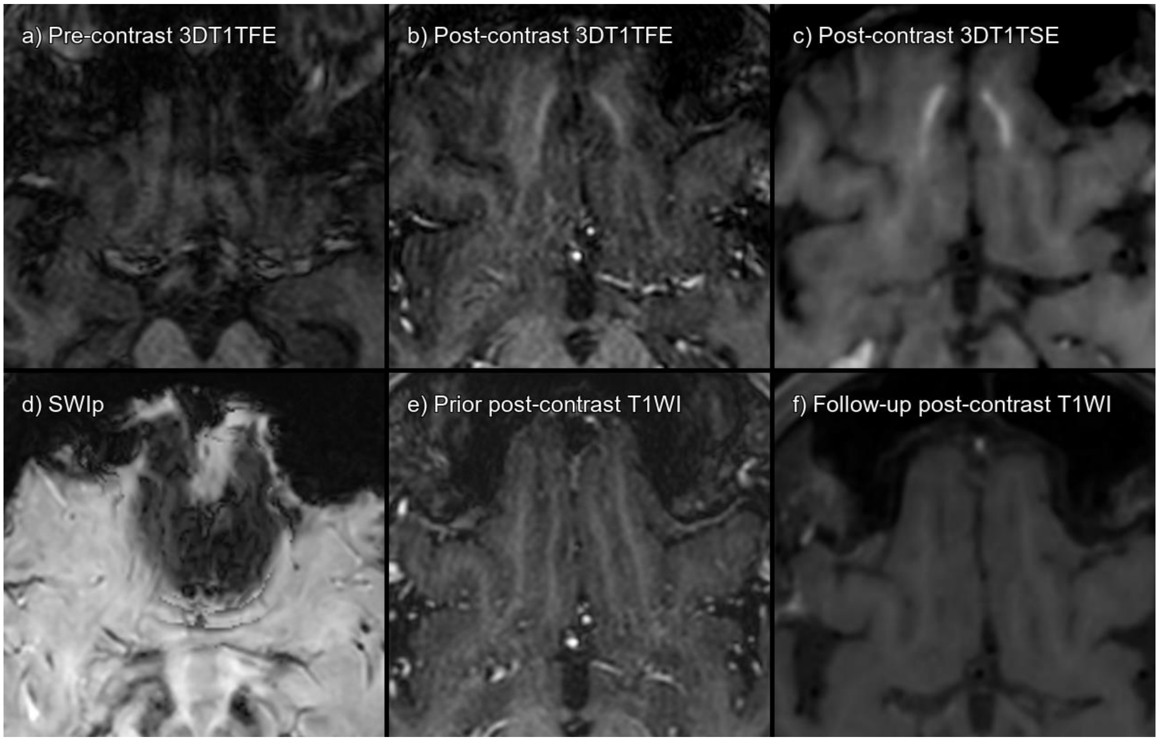


Supplementary figure 67. Lesion 248_03 a) 3DT1TFE without contrast, b) 3DT1TFE post-contrast, c) 3DT1TSE post-contrast, d) SWIp, e) Prior post-contrast T1-weighted imaging 3 months pre-baseline, f) Follow-up post-contrast T1-weighted imaging 4 months post-baseline.

True enhancing lesion that has disappeared on follow-up MRI. Lesion 248_03 refers to the left rectal gyrus lesion. This region is not discernible on SWIp due to strong susceptibility artifacts.

| **CONSENSUS** | **True Enhancing MS Lesion** | | | |
|---|---|---|---|---|
| | **TFE sequence** | | **TSE sequence** | |
| Visible in sequence | Yes | | Yes | |
| | **Reader 1** | **Reader 2** | **Reader 1** | **Reader 2** |
| **Detected** | No | No | No | Yes |

Abbreviations: Multiple sclerosis (MS), Turbo Field Echo (TFE), Turbo Spin Echo (TSE), Susceptibility Weighted Imaging with phase enhancement (SWIp).

#### LESION 249_01

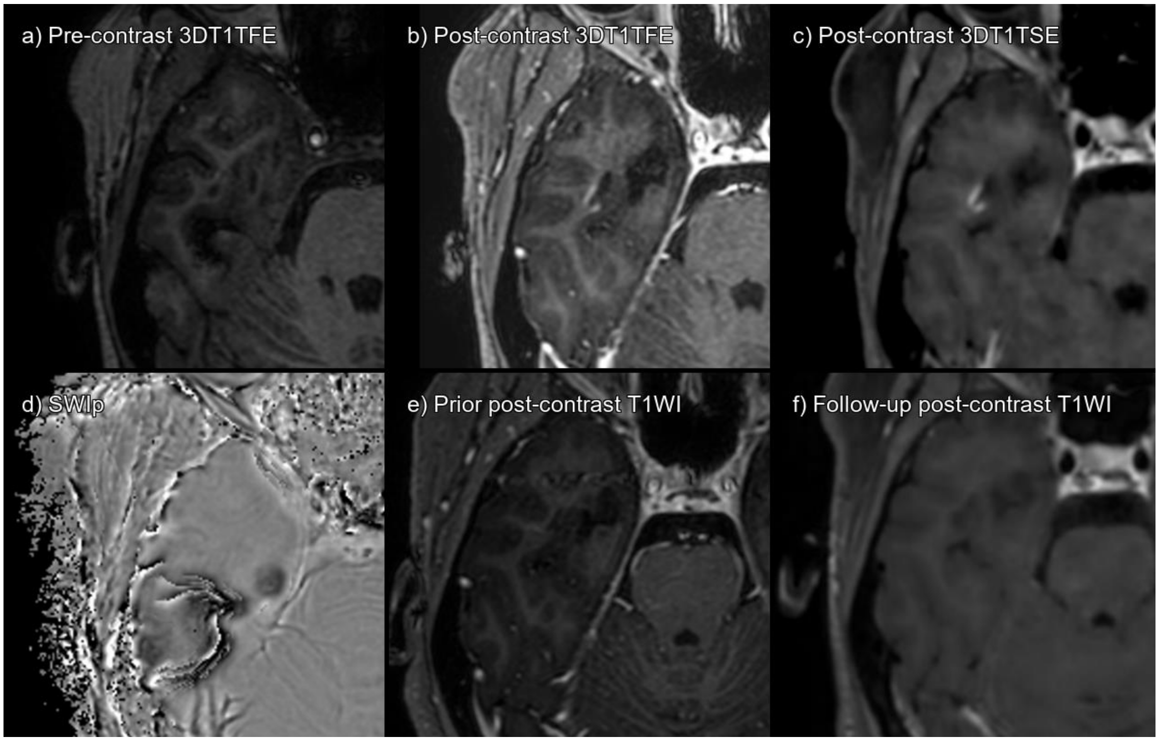


Supplementary figure 68. Lesion 249_01 a) 3DT1TFE without contrast, b) 3DT1TFE post-contrast, c) 3DT1TSE post-contrast, d) SWIp phase, e) Prior post-contrast T1-weighted imaging 6 months pre-baseline, f) Follow-up post-contrast T1-weighted imaging 9 months post-baseline.

True enhancing lesion that has disappeared on follow-up MRI.

| **CONSENSUS** | **True Enhancing MS Lesion** | | | |
|---|---|---|---|---|
| | **TFE sequence** | | **TSE sequence** | |
| Visible in sequence | Yes | | Yes | |
| | **Reader 1** | **Reader 2** | **Reader 1** | **Reader 2** |
| **Detected** | Yes | Yes | Yes | Yes |

Abbreviations: Multiple sclerosis (MS), Turbo Field Echo (TFE), Turbo Spin Echo (TSE), Susceptibility Weighted Imaging with phase enhancement (SWIp).

#### LESION 249_02

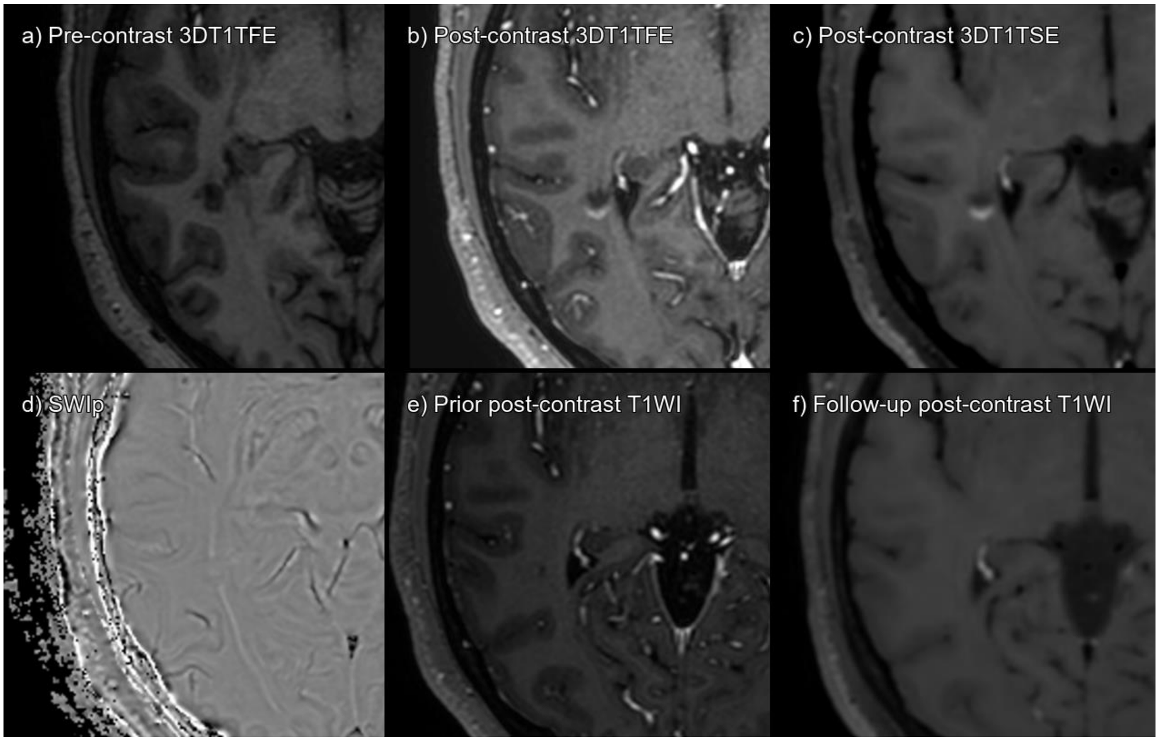


Supplementary figure 69. Lesion 249_02 a) 3DT1TFE without contrast, b) 3DT1TFE post-contrast, c) 3DT1TSE post-contrast, d) SWIp phase, e) Prior post-contrast T1-weighted imaging 6 months pre-baseline, f) Follow-up post-contrast T1-weighted imaging 9 months post-baseline.

True enhancing lesion that has disappeared on follow-up MRI.

| **CONSENSUS** | **True Enhancing MS Lesion** | | | |
|---|---|---|---|---|
| | **TFE sequence** | | **TSE sequence** | |
| Visible in sequence | Yes | | Yes | |
| | **Reader 1** | **Reader 2** | **Reader 1** | **Reader 2** |
| **Detected** | Yes | Yes | Yes | Yes |

Abbreviations: Multiple sclerosis (MS), Turbo Field Echo (TFE), Turbo Spin Echo (TSE), Susceptibility Weighted Imaging with phase enhancement (SWIp).

#### LESION 250_01

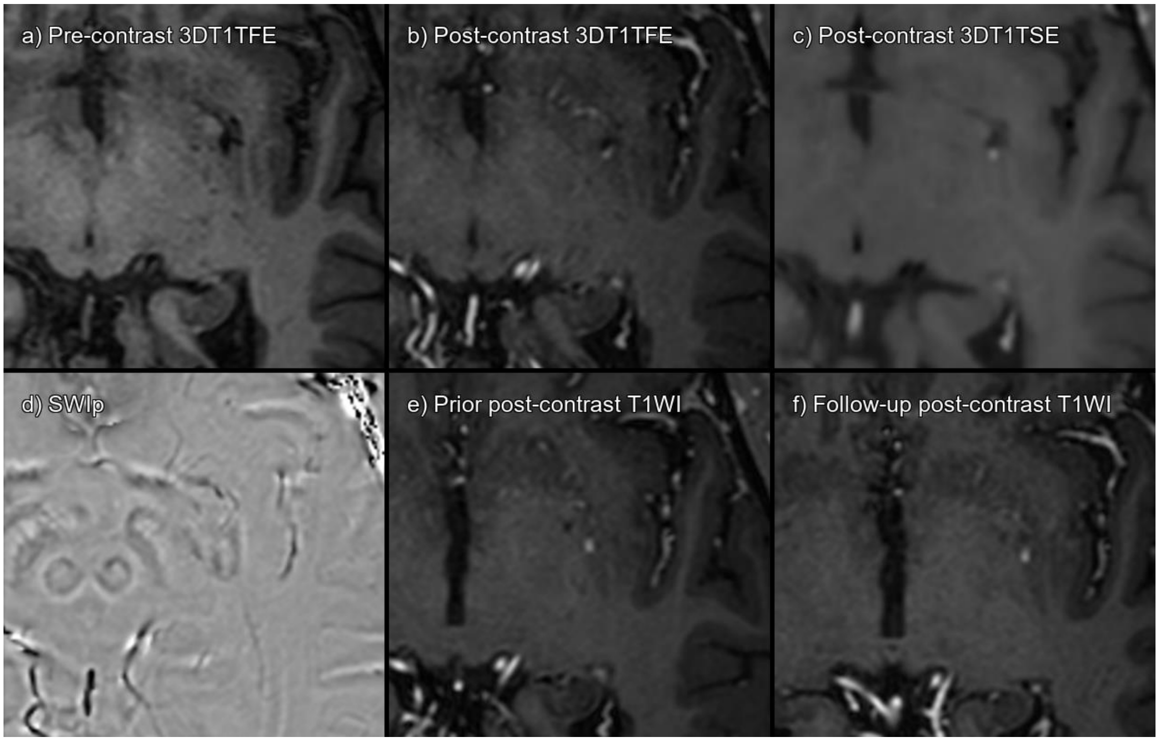


Supplementary figure 70. Lesion 250_01 a) 3DT1TFE without contrast, b) 3DT1TFE post-contrast, c) 3DT1TSE post-contrast, d) SWIp phase, e) Prior post-contrast T1-weighted imaging 9 months pre-baseline, f) Follow-up post-contrast T1-weighted imaging 1 year post-baseline.

Enhancing lesion, that is already present on pre- and post-baseline examination. On SWIp correspond to a vessel. Consensus established false positive enhancement on both 3DT1TFE and 3DT1TSE, vascular enhancement.

| **CONSENSUS** | **Not acute MS lesion enhancement** | | | |
|---|---|---|---|---|
| | **TFE sequence** | | **TSE sequence** | |
| Visible in sequence | Yes | | Yes | |
| | **Reader 1** | **Reader 2** | **Reader 1** | **Reader 2** |
| **Detected** | Yes | No | Yes | Yes |

Abbreviations: Multiple sclerosis (MS), Turbo Field Echo (TFE), Turbo Spin Echo (TSE), Susceptibility Weighted Imaging with phase enhancement (SWIp).

### LESION 258_01

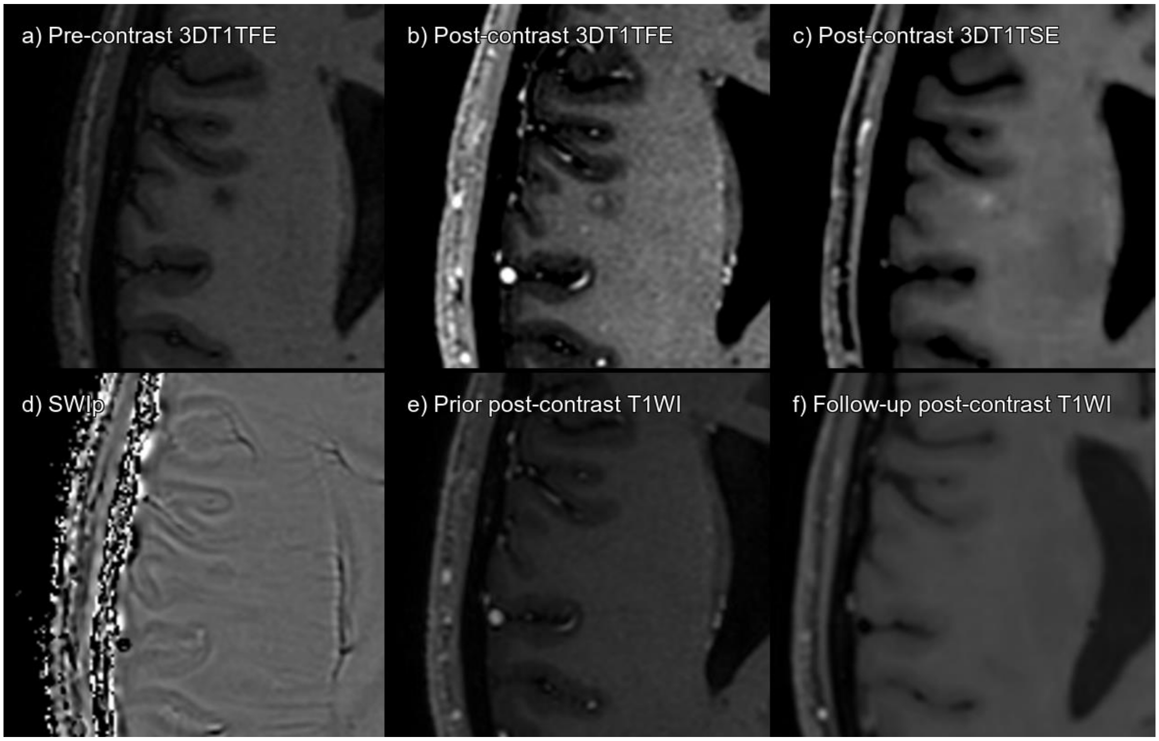


Supplementary figure 71. Lesion 258_01 a) 3DT1TFE without contrast, b) 3DT1TFE post-contrast, c) 3DT1TSE post-contrast, d) SWIp phase, e) Prior post-contrast T1-weighted imaging 1 year pre-baseline, f) Follow-up post-contrast T1-weighted imaging 1 year post-baseline.

True enhancing lesion that has disappeared on follow-up MRI.

| **CONSENSUS** | **True Enhancing MS Lesion** | | | |
|---|---|---|---|---|
| | **TFE sequence** | | **TSE sequence** | |
| Visible in sequence | Yes | | Yes | |
| | **Reader 1** | **Reader 2** | **Reader 1** | **Reader 2** |
| **Detected** | Yes | No | No | No |

Abbreviations: Multiple sclerosis (MS), Turbo Field Echo (TFE), Turbo Spin Echo (TSE), Susceptibility Weighted Imaging with phase enhancement (SWIp).

#### LESION 268_01

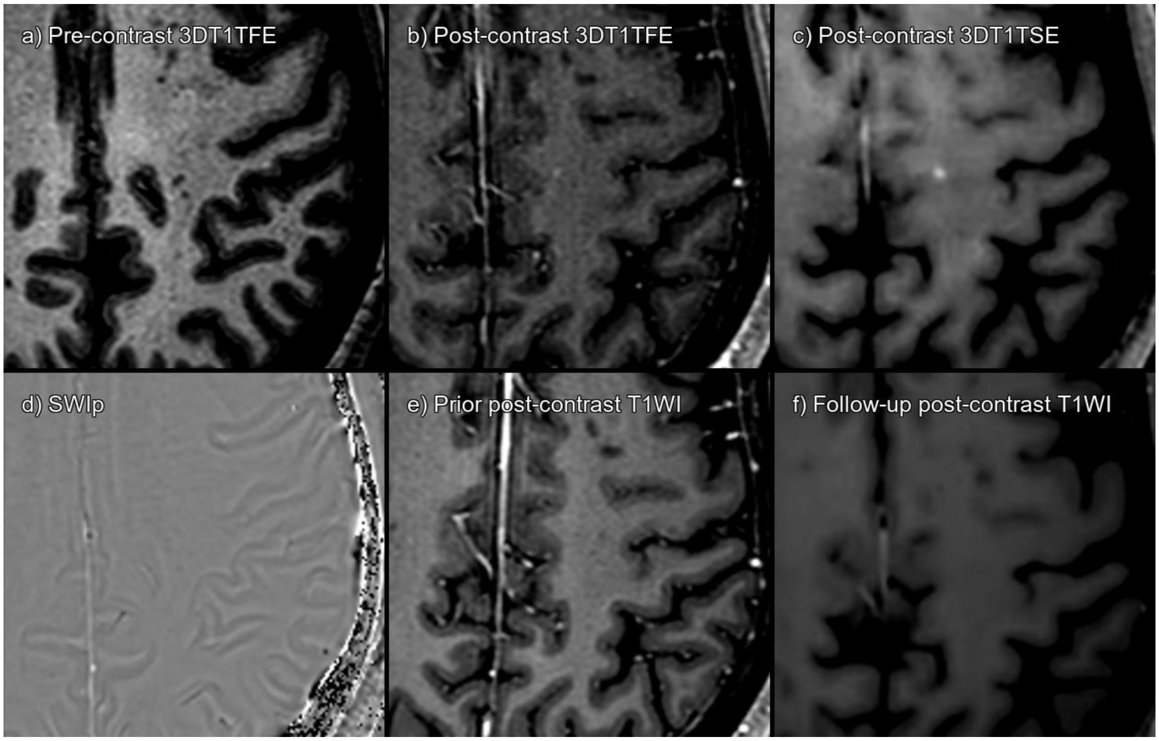


Supplementary figure 72. Lesion 268_01 a) 3DT1TFE without contrast, b) 3DT1TFE post-contrast, c) 3DT1TSE post-contrast, d) SWIp phase, e) Prior post-contrast T1-weighted imaging 5 months pre-baseline, f) Follow-up post-contrast T1-weighted imaging 7 months post-baseline.

True enhancing lesion that has disappeared on follow-up MRI.

| **CONSENSUS** | **True Enhancing MS Lesion** | | | |
|---|---|---|---|---|
| | **TFE sequence** | | **TSE sequence** | |
| Visible in sequence | Yes | | Yes | |
| | **Reader 1** | **Reader 2** | **Reader 1** | **Reader 2** |
| **Detected** | No | No | Yes | Yes |

Abbreviations: Multiple sclerosis (MS), Turbo Field Echo (TFE), Turbo Spin Echo (TSE), Susceptibility Weighted Imaging with phase enhancement (SWIp).

#### LESION 268_02

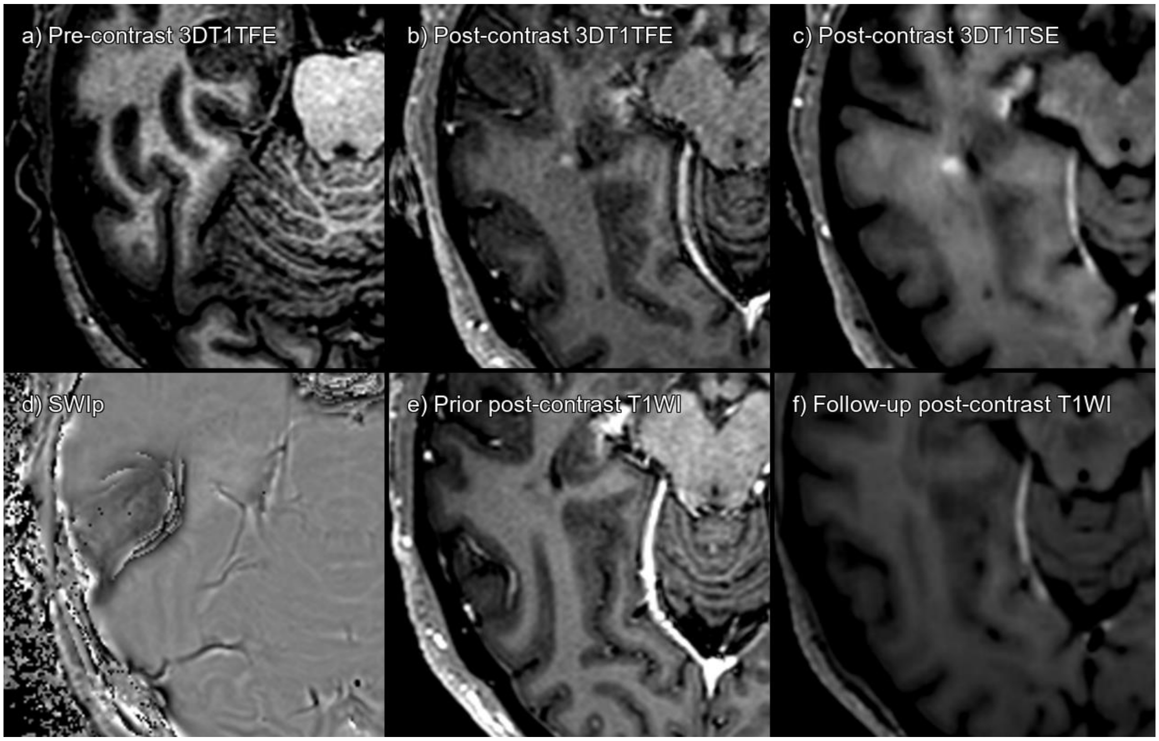


Supplementary figure 73. Lesion 268_02 a) 3DT1TFE without contrast, b) 3DT1TFE post-contrast, c) 3DT1TSE post-contrast, d) SWIp phase, e) Prior post-contrast T1-weighted imaging 5 months pre-baseline, f) Follow-up post-contrast T1-weighted imaging 7 months post-baseline.

True enhancing lesion that has disappeared on follow-up MRI.

| **CONSENSUS** | **True Enhancing MS Lesion** | | | |
|---|---|---|---|---|
| | **TFE sequence** | | **TSE sequence** | |
| Visible in sequence | Yes | | Yes | |
| | **Reader 1** | **Reader 2** | **Reader 1** | **Reader 2** |
| **Detected** | No | No | Yes | Yes |

Abbreviations: Multiple sclerosis (MS), Turbo Field Echo (TFE), Turbo Spin Echo (TSE), Susceptibility Weighted Imaging with phase enhancement (SWIp).

#### LESION 272_01

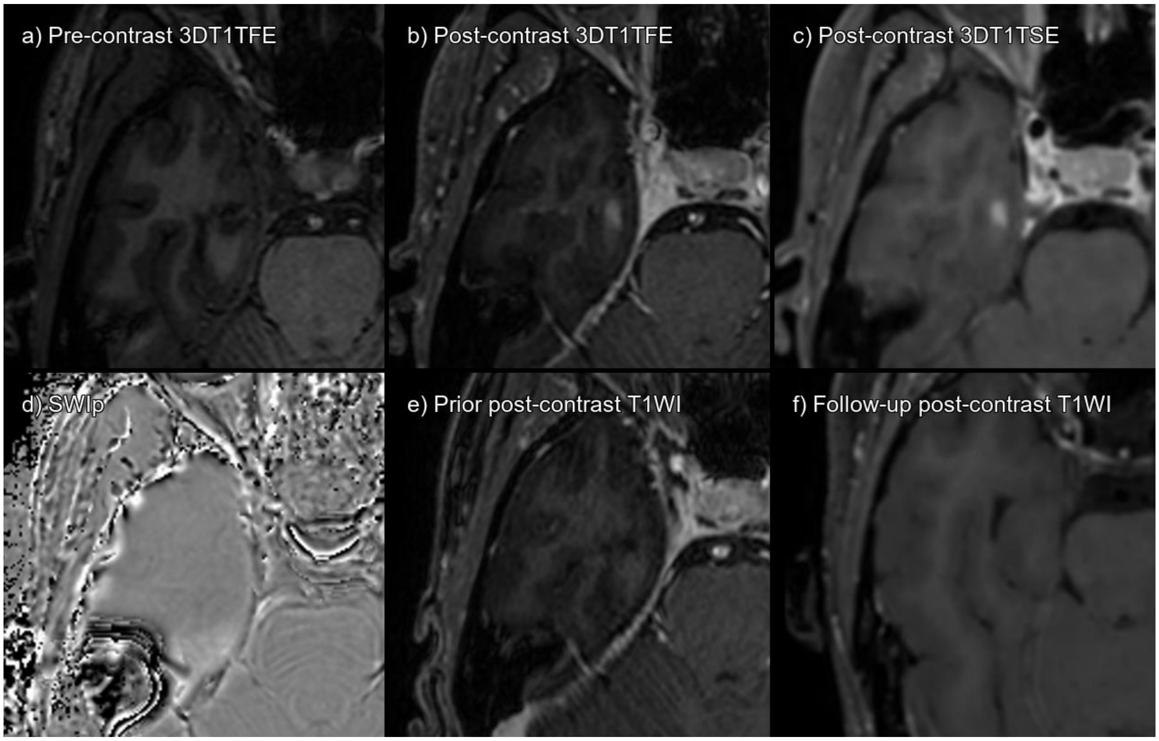


Supplementary figure 74. Lesion 272_01 a) 3DT1TFE without contrast, b) 3DT1TFE post-contrast, c) 3DT1TSE post-contrast, d) SWIp phase, e) Prior post-contrast T1-weighted imaging 5 months pre-baseline, f) Follow-up post-contrast T1-weighted imaging 9 months post-baseline.

True enhancing lesion that has disappeared on follow-up MRI.

| **CONSENSUS** | **True Enhancing MS Lesion** | | | |
|---|---|---|---|---|
| | **TFE sequence** | | **TSE sequence** | |
| Visible in sequence | Yes | | Yes | |
| | **Reader 1** | **Reader 2** | **Reader 1** | **Reader 2** |
| **Detected** | Yes | Yes | Yes | Yes |

Abbreviations: Multiple sclerosis (MS), Turbo Field Echo (TFE), Turbo Spin Echo (TSE), Susceptibility Weighted Imaging with phase enhancement (SWIp).

#### LESION 272_02

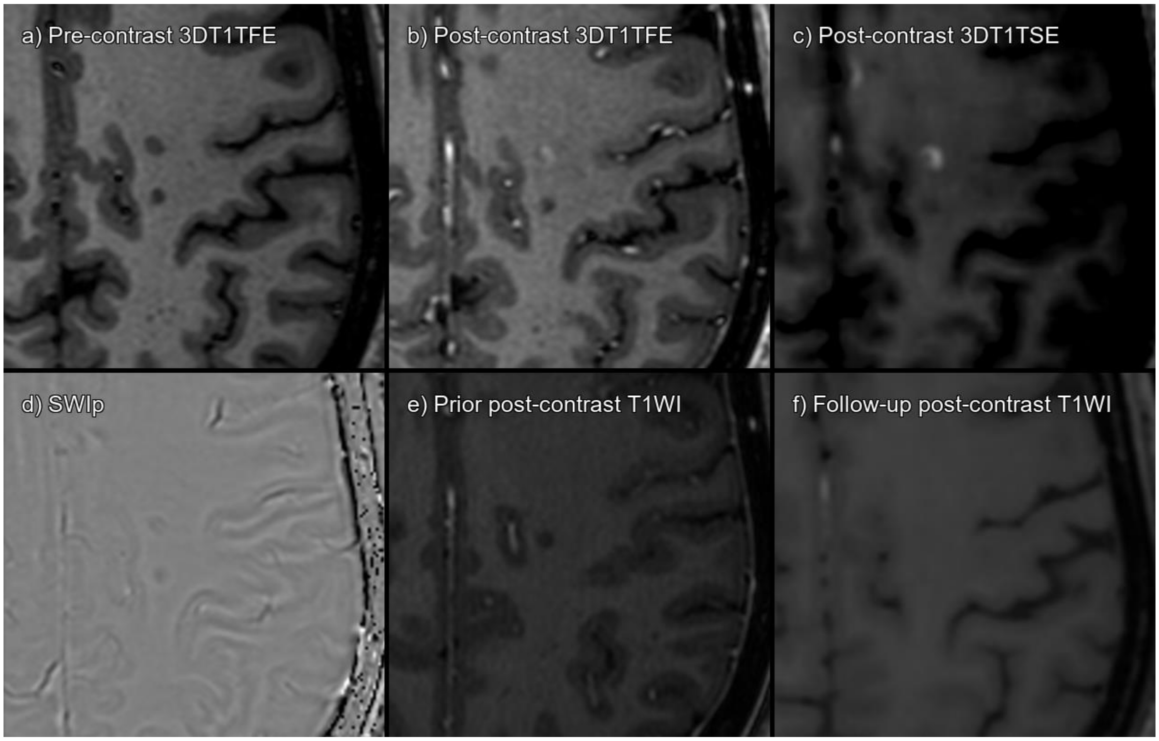


Supplementary figure 75. Lesion 272_02 a) 3DT1TFE without contrast, b) 3DT1TFE post-contrast, c) 3DT1TSE post-contrast, d) SWIp phase, e) Prior post-contrast T1-weighted imaging 5 months pre-baseline, f) Follow-up post-contrast T1-weighted imaging 9 months post-baseline.

True enhancing lesion that has disappeared on follow-up MRI.

| **CONSENSUS** | **True Enhancing MS Lesion** | | | |
|---|---|---|---|---|
| | **TFE sequence** | | **TSE sequence** | |
| Visible in sequence | Yes | | Yes | |
| | **Reader 1** | **Reader 2** | **Reader 1** | **Reader 2** |
| **Detected** | Yes | Yes | Yes | Yes |

Abbreviations: Multiple sclerosis (MS), Turbo Field Echo (TFE), Turbo Spin Echo (TSE), Susceptibility Weighted Imaging with phase enhancement (SWIp).

#### LESION 272_03

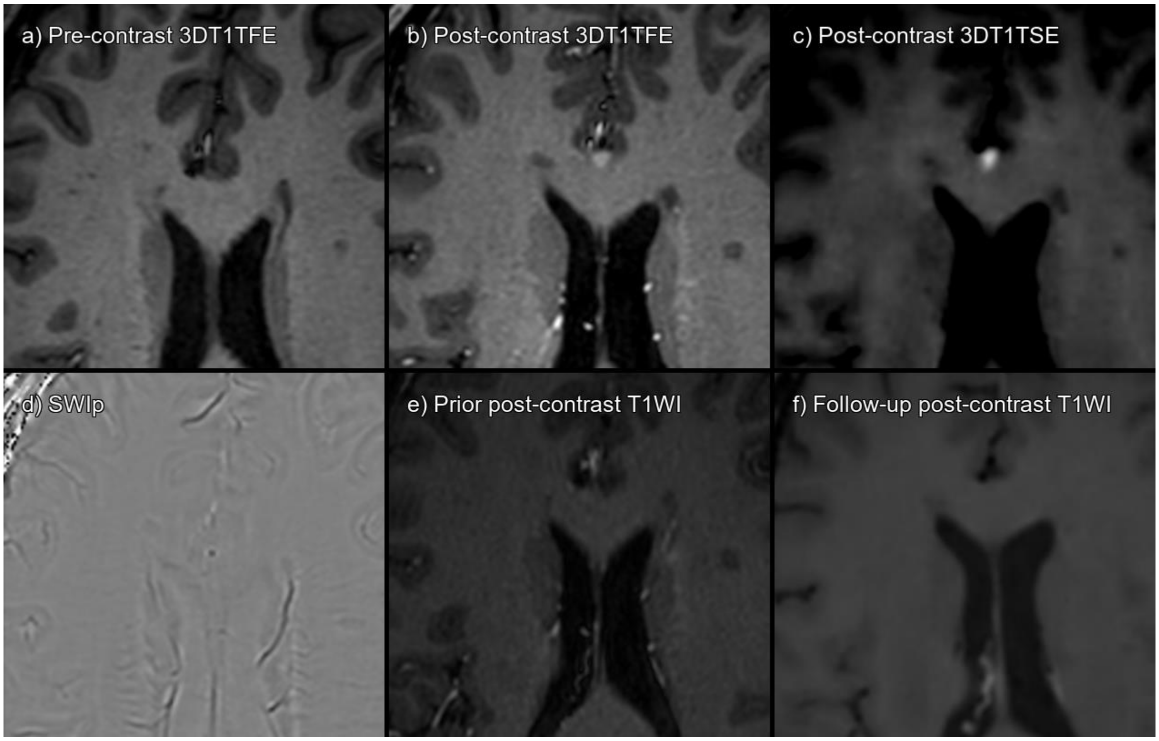


Supplementary figure 76. Lesion 272_03 a) 3DT1TFE without contrast, b) 3DT1TFE post-contrast, c) 3DT1TSE post-contrast, d) SWIp phase, e) Prior post-contrast T1-weighted imaging 5 months pre-baseline, f) Follow-up post-contrast T1-weighted imaging 9 months post-baseline.

True enhancing lesion that has disappeared on follow-up MRI.

| **CONSENSUS** | **True Enhancing MS Lesion** | | | |
|---|---|---|---|---|
| | **TFE sequence** | | **TSE sequence** | |
| Visible in sequence | Yes | | Yes | |
| | **Reader 1** | **Reader 2** | **Reader 1** | **Reader 2** |
| **Detected** | No | No | Yes | Yes |

Abbreviations: Multiple sclerosis (MS), Turbo Field Echo (TFE), Turbo Spin Echo (TSE), Susceptibility Weighted Imaging with phase enhancement (SWIp).

#### LESION 287_01

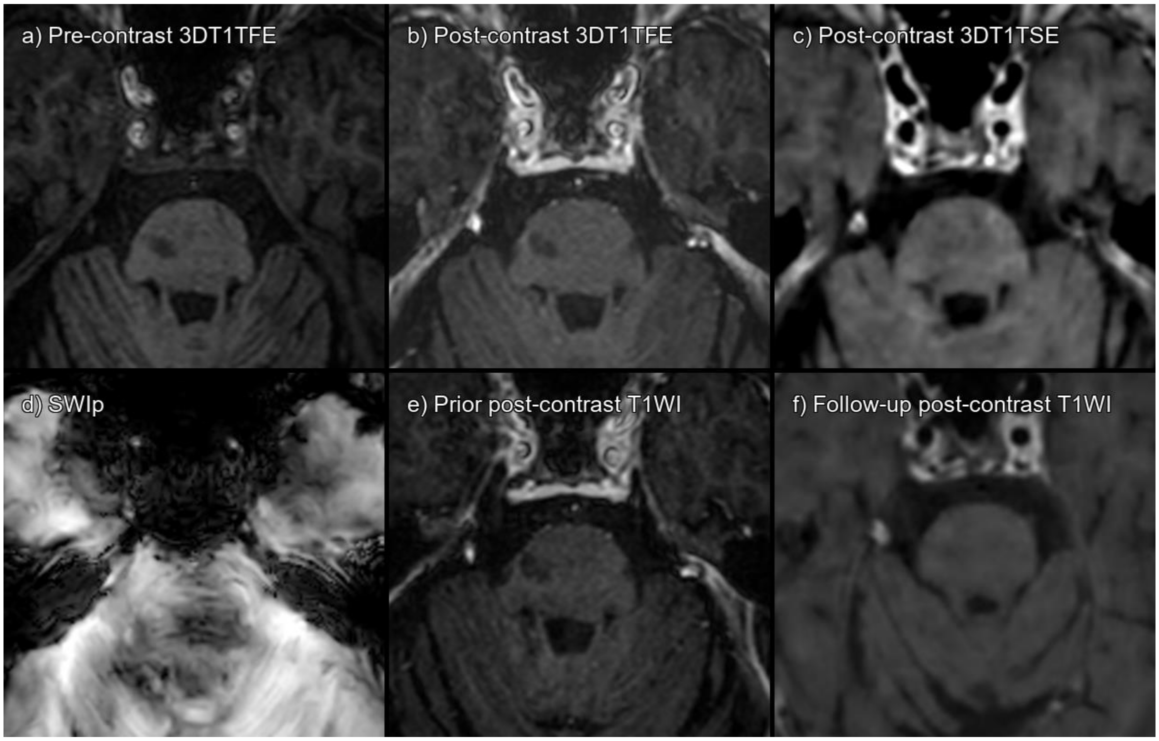


Supplementary figure 77. Lesion 287_01 a) 3DT1TFE without contrast, b) 3DT1TFE post-contrast, c) 3DT1TSE post-contrast, d) SWIp phase, e) Prior post-contrast T1-weighted imaging 1 year pre-baseline, f) Follow-up post-contrast T1-weighted imaging 6 months post-baseline.

Subtle hyperintensity in post-contrast 3DT1TSE in anterior left margin of the pons, in a relatively low-quality image with movement artifacts. False positive, artefactual in nature.

| **CONSENSUS** | **Not acute MS lesion enhancement** | | | |
|---|---|---|---|---|
| | **TFE sequence** | | **TSE sequence** | |
| Visible in sequence | No | | No | |
| | **Reader 1** | **Reader 2** | **Reader 1** | **Reader 2** |
| **Detected** | No | No | No | Yes |

Abbreviations: Multiple sclerosis (MS), Turbo Field Echo (TFE), Turbo Spin Echo (TSE), Susceptibility Weighted Imaging with phase enhancement (SWIp).

#### LESION 338_01

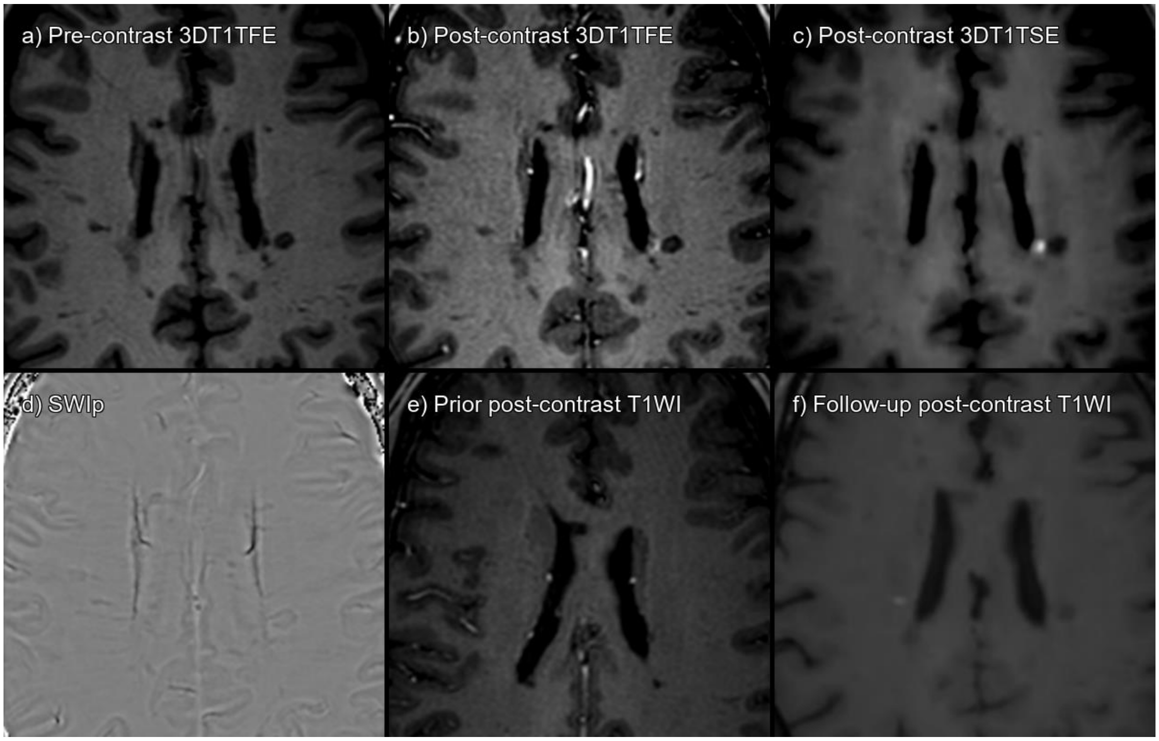


Supplementary figure 78. Lesion 338_01 a) 3DT1TFE without contrast, b) 3DT1TFE post-contrast, c) 3DT1TSE post-contrast, d) SWIp phase, e) Prior post-contrast T1-weighted imaging 1 year pre-baseline, f) Follow-up post-contrast T1-weighted imaging 4 months post-baseline.

True enhancing lesion that has disappeared on follow-up MRI.

| **CONSENSUS** | **True Enhancing MS Lesion** | | | |
|---|---|---|---|---|
| | **TFE sequence** | | **TSE sequence** | |
| Visible in sequence | Yes | | Yes | |
| | **Reader 1** | **Reader 2** | **Reader 1** | **Reader 2** |
| **Detected** | Yes | Yes | Yes | Yes |

Abbreviations: Multiple sclerosis (MS), Turbo Field Echo (TFE), Turbo Spin Echo (TSE), Susceptibility Weighted Imaging with phase enhancement (SWIp).

#### LESION 338_02

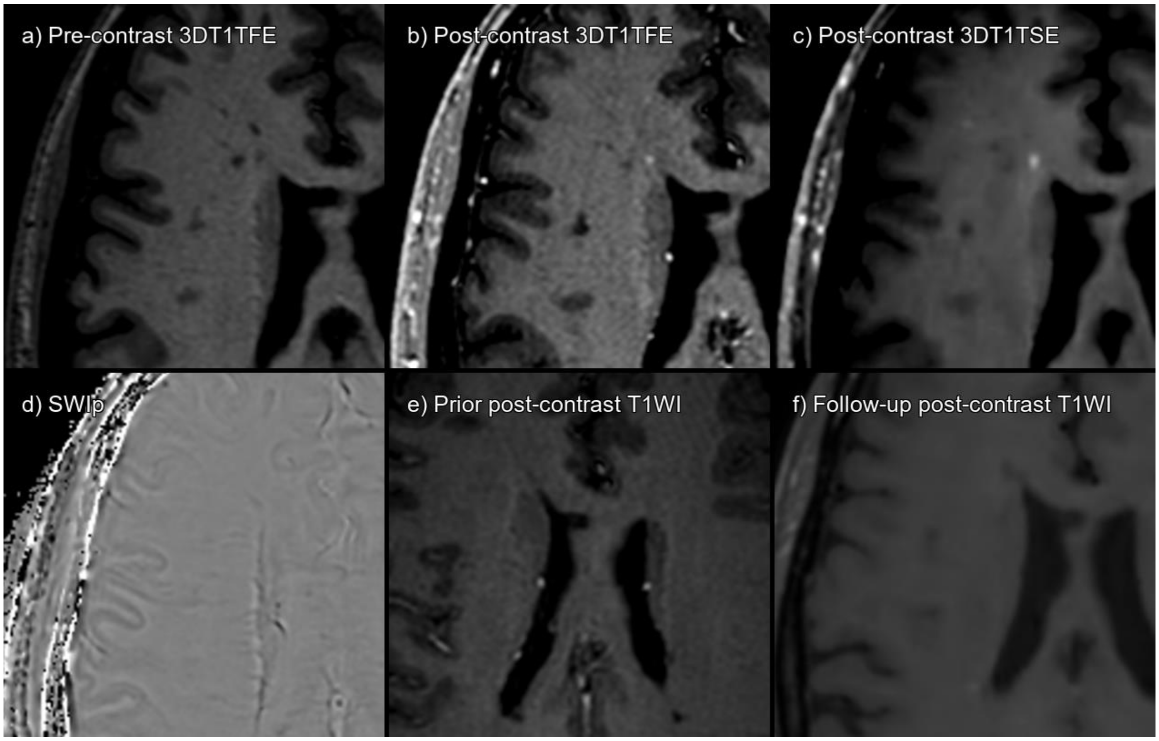


Supplementary figure 79. Lesion 338_02 a) 3DT1TFE without contrast, b) 3DT1TFE post-contrast, c) 3DT1TSE post-contrast, d) SWIp phase, e) Prior post-contrast T1-weighted imaging 1 year pre-baseline, f) Follow-up post-contrast T1-weighted imaging 4 months post-baseline.

True enhancing lesion that has disappeared on follow-up MRI.

| **CONSENSUS** | **True Enhancing MS Lesion** | | | |
|---|---|---|---|---|
| | **TFE sequence** | | **TSE sequence** | |
| Visible in sequence | Yes | | Yes | |
| | **Reader 1** | **Reader 2** | **Reader 1** | **Reader 2** |
| **Detected** | Yes | Yes | Yes | Yes |

Abbreviations: Multiple sclerosis (MS), Turbo Field Echo (TFE), Turbo Spin Echo (TSE), Susceptibility Weighted Imaging with phase enhancement (SWIp).

#### LESION 338_03

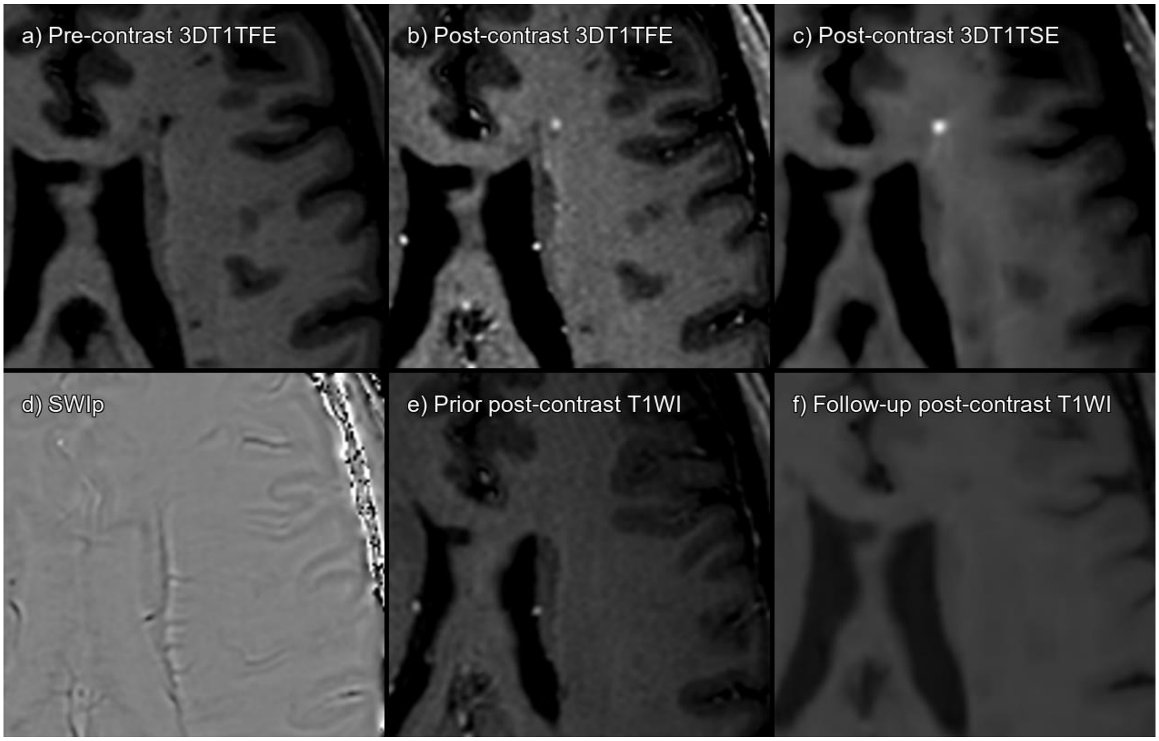


Supplementary figure 80. Lesion 338_03 a) 3DT1TFE without contrast, b) 3DT1TFE post-contrast, c) 3DT1TSE post-contrast, d) SWIp phase, e) Prior post-contrast T1-weighted imaging 1 year pre-baseline, f) Follow-up post-contrast T1-weighted imaging 4 months post-baseline.

True enhancing lesion that has disappeared on follow-up MRI.

| **CONSENSUS** | **True Enhancing MS Lesion** | | | |
|---|---|---|---|---|
| | **TFE sequence** | | **TSE sequence** | |
| Visible in sequence | Yes | | Yes | |
| | **Reader 1** | **Reader 2** | **Reader 1** | **Reader 2** |
| **Detected** | Yes | Yes | Yes | Yes |

Abbreviations: Multiple sclerosis (MS), Turbo Field Echo (TFE), Turbo Spin Echo (TSE), Susceptibility Weighted Imaging with phase enhancement (SWIp).

#### LESION 338_04

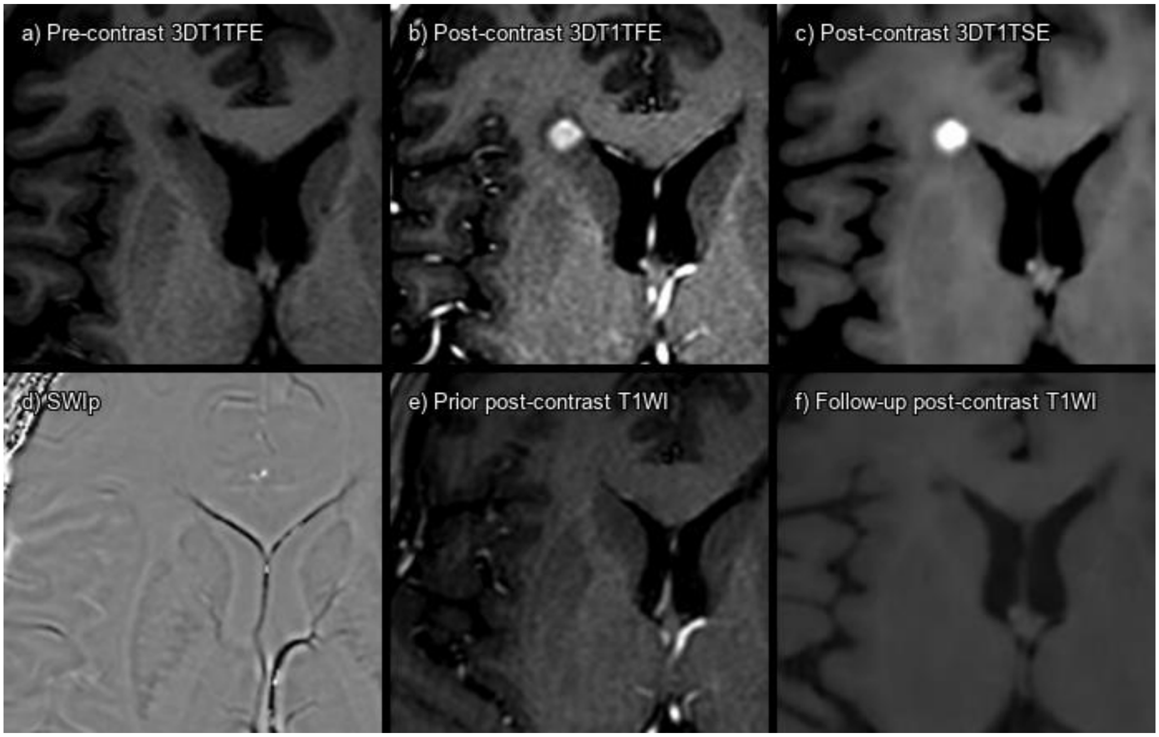


Supplementary figure 81. Lesion 338_04 a) 3DT1TFE without contrast, b) 3DT1TFE post-contrast, c) 3DT1TSE post-contrast, d) SWIp phase, e) Prior post-contrast T1-weighted imaging 1 year pre-baseline, f) Follow-up post-contrast T1-weighted imaging 4 months post-baseline.

True enhancing lesion that has disappeared on follow-up MRI.

| **CONSENSUS** | **True Enhancing MS Lesion** | | | |
|---|---|---|---|---|
| | **TFE sequence** | | **TSE sequence** | |
| Visible in sequence | Yes | | Yes | |
| | **Reader 1** | **Reader 2** | **Reader 1** | **Reader 2** |
| **Detected** | Yes | Yes | Yes | Yes |

Abbreviations: Multiple sclerosis (MS), Turbo Field Echo (TFE), Turbo Spin Echo (TSE), Susceptibility Weighted Imaging with phase enhancement (SWIp).

#### LESION 338_05

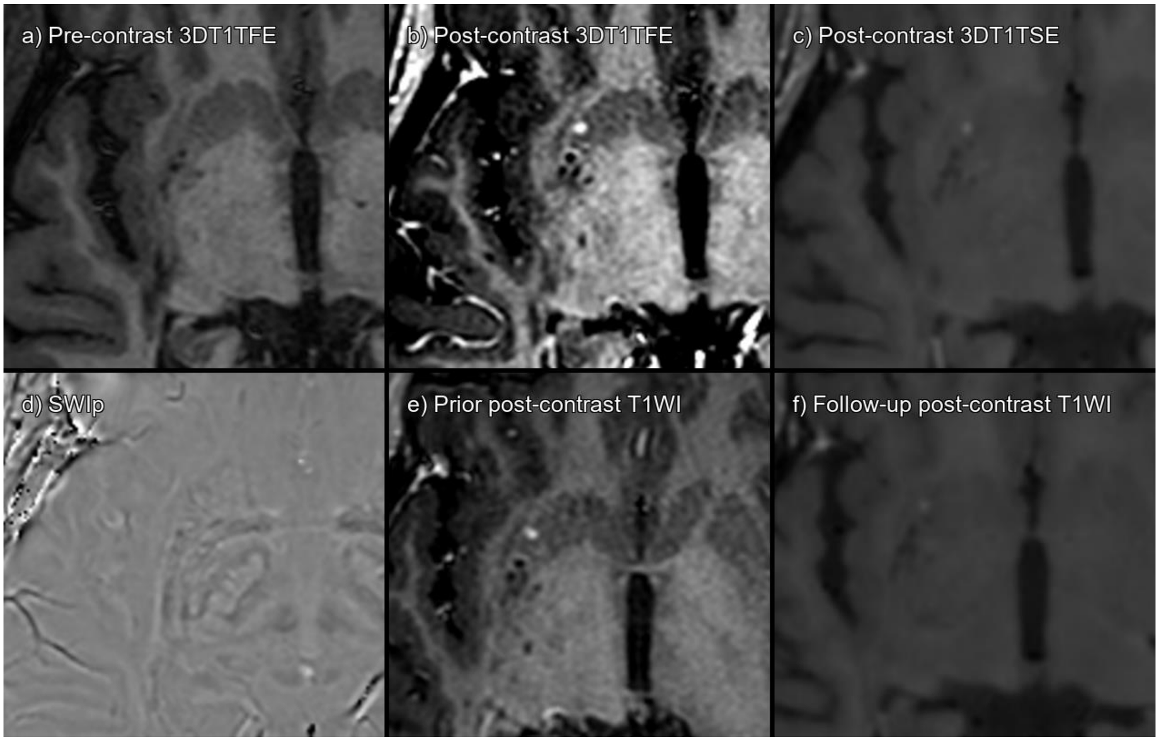


Supplementary figure 82. Lesion 338_05 a) 3DT1TFE without contrast, b) 3DT1TFE post-contrast, c) 3DT1TSE post-contrast, d) SWIp phase, e) Prior post-contrast T1-weighted imaging 1 year pre-baseline, f) Follow-up post-contrast T1-weighted imaging 4 months post-baseline.

Enhancing spot in the right putamen that is already present in previous and persists on follow-up images. There is a vessel in its center on SWIp. False positive, probably vascular enhancement.

| **CONSENSUS** | **Not acute MS lesion enhancement** | | | |
|---|---|---|---|---|
| | **TFE sequence** | | **TSE sequence** | |
| Visible in sequence | Yes | | Yes | |
| | **Reader 1** | **Reader 2** | **Reader 1** | **Reader 2** |
| **Detected** | No | No | Yes | Yes |

Abbreviations: Multiple sclerosis (MS), Turbo Field Echo (TFE), Turbo Spin Echo (TSE), Susceptibility Weighted Imaging with phase enhancement (SWIp).

#### LESION 338_06

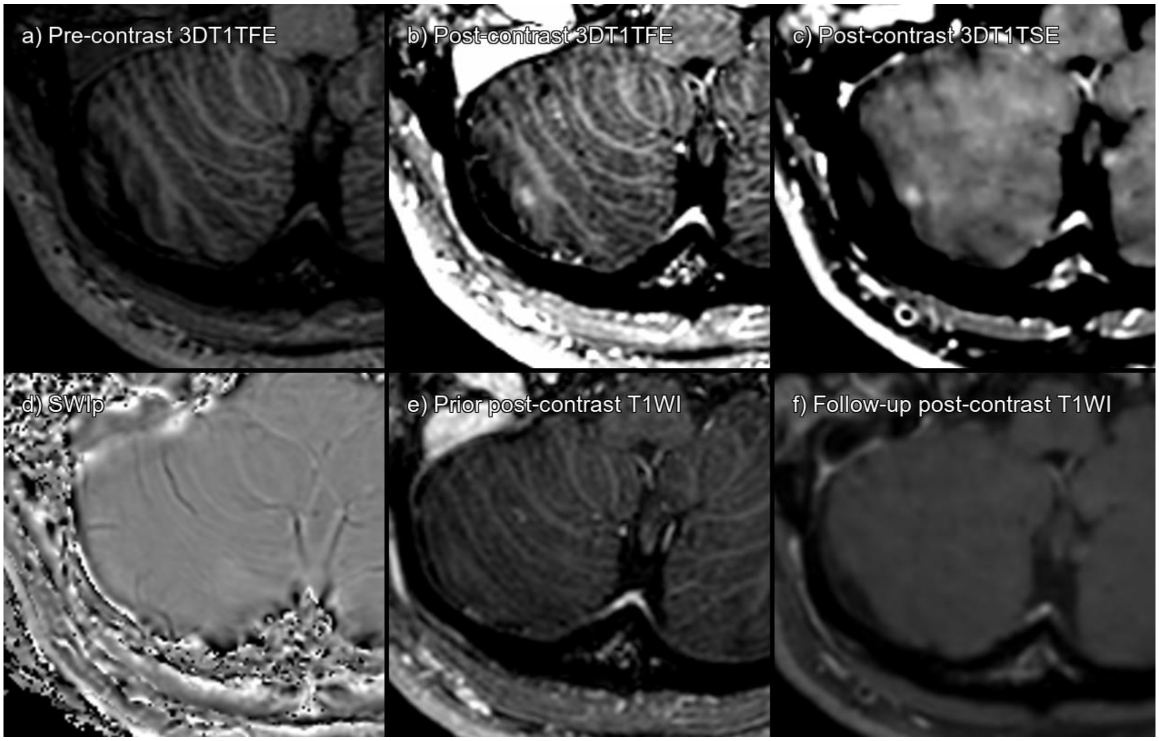


Supplementary figure 83. Lesion 338_06 a) 3DT1TFE without contrast, b) 3DT1TFE post-contrast, c) 3DT1TSE post-contrast, d) SWIp phase, e) Prior post-contrast T1-weighted imaging 1 year pre-baseline, f) Follow-up post-contrast T1-weighted imaging 4 months post-baseline.

True enhancing lesion that has disappeared on follow-up MRI.

| **CONSENSUS** | **True Enhancing MS Lesion** | | | |
|---|---|---|---|---|
| | **TFE sequence** | | **TSE sequence** | |
| Visible in sequence | Yes | | Yes | |
| | **Reader 1** | **Reader 2** | **Reader 1** | **Reader 2** |
| **Detected** | No | No | No | Yes |

Abbreviations: Multiple sclerosis (MS), Turbo Field Echo (TFE), Turbo Spin Echo (TSE), Susceptibility Weighted Imaging with phase enhancement (SWIp).
